# Sex-specific metabolic signatures of adiposity associated with clinical biomarkers in the UK Biobank

**DOI:** 10.1101/2025.01.23.25321010

**Authors:** Christos K. Papagiannopoulos, Georgios Markozannes, Christos V. Chalitsios, Sofia Christakoudi, Marc J Gunter, Laure Dossus, Richard M. Martin, Ioanna Tzoulaki, Christopher Papandreou, Konstantinos K. Tsilidis

**Affiliations:** Department of Hygiene and Epidemiology, School of Medicine, University of Ioannina, Ioannina, Greece; Department of Epidemiology and Biostatistics, School of Public Health, Imperial College London, London, UK; Nutrition and Metabolism Branch, International Agency for Research on Cancer, World Health Organization, Lyon, France; Nutrition and Metabolism Branch, International Agency for Research on Cancer, Lyon, France; Population Health Sciences, Bristol Medical School, University of Bristol, Bristol, United Kingdom; MRC Integrative Epidemiology Unit, University of Bristol, Bristol, United Kingdom; NIHR Bristol Biomedical Research Centre, Hospitals Bristol and Weston NHS Foundation Trust and the University of Bristol, Bristol, United Kingdom; Biomedical Research Foundation of the Academy of Athens, Athens, Greece; Institute of Health Pere Virgili (IISPV), Reus, Spain; Department of Nutrition and Dietetics Sciences, School of Health Sciences, Hellenic Mediterranean University (HMU), Siteia, Greece

**Keywords:** adiposity, metabolomics, machine learning, biomarkers, epidemiology, UK Biobank

## Abstract

Excessive adiposity increases disease risk, yet the exact underlying mechanisms remain unknown. We compared sex-specific metabolic signatures (MSs) of adiposity indices (non-allometric: body fat %, waist circumference, hip circumference, waist-to-hip ratio, body mass index; allometric: a body shape index, hip index, waist-to-hip-index ratio) and examined their associations with 29 clinical biomarkers in 151,526 UK Biobank participants. MSs performance was validated in an independent cohort. In females, MSs mainly consisted of lipoprotein particle concentrations, apolipoproteins, fatty acids and inflammation-linked glycoprotein acetyls, whereas in males lipoproteins rich in cholesteryl esters and aromatic/branched-chain amino acids predominated. The highest percentages of common metabolites were observed between non-allometric adiposity indices (median: 42.4%; range: 9%–56%). MSs were independently associated with over 25 biomarkers with differences observed by sex and adiposity index, and these associations were stronger compared to the respective phenotypic associations. MSABSI was found to be more atherogenic, whereas MSHI was more favourable for health. This study highlights i) that different regions of adipose tissue undergo distinct metabolic processes overall and by sex, each having unique impact on health, and ii) the importance of considering metabolic factors beyond simple adiposity indices in assessing health risk.

## Introduction

Increased adiposity represents a major public health concern (1–3). Evidence from recent observational and Mendelian randomization studies consistently shows that higher levels of total fat mass, general, and abdominal obesity are associated with an increased risk of cardiometabolic diseases, several cancers, and other chronic diseases (4–6). In contrast, higher gluteofemoral fat deposition has been associated with a lower risk of adverse health outcomes (7, 8). The differential associations of adiposity indices on health outcomes (7, 9, 10), could be partially explained by differences in their effects on metabolism. Metabolomics combined with applied machine learning (11) can advance our understanding of the molecular mechanisms linking adiposity to health outcomes (12–14). By comparing the metabolic signatures (MSs) of different adiposity indices we can gain insights into how variations in adiposity impact metabolism (15).

Adiposity tissue deposition and function differ by sex due to hormone metabolism differences (16, 17), which could partially explain its sexual dimorphism on health outcomes (18–20). However, the exact mechanisms through which adiposity exerts sex-specific health effects are not completely understood. Few studies suggest sex-dependent differences in metabolic profiles associated with adiposity in adults (21–23) with the most prominent differences being higher concentrations of lipoprotein particles and fatty acids in females and higher concentrations of amino acids in males. However, these studies have limitations, such as lack of replication of the findings, limiting the robustness and generalizability of the findings.

Clinical biomarkers serve as risk factors for non-communicable diseases and diagnostic measures for clinical conditions. Associating the MSs of adiposity indices with intermediate biomarkers of health could provide insights into the relationship between metabolism and health risk. Previous research suggests that MSs may explain the association between adiposity indices and several metabolic biomarkers (13) and prostate cancer (14). However, whether the shared and distinct metabolic processes across multiple adiposity indices can explain their respective effects on health is unclear.

In this study, we aimed to identify sex-specific MSs associated with different adiposity indices using machine learning techniques and metabolomics data from over 150,000 participants in the UK Biobank (UKBB) and validated our findings in the Epirus Health Study (EHS). Then, we compared the MS and examined their associations with 29 clinical biomarkers and explained these associations through their shared and distinct metabolites.

## Results

### Populations’ characteristics

In both UKBB and EHS, characteristics of participants including non-allometric (body fat [BF] %, body mass index [BMI], waist circumference [WC], hip circumference [HC], waist to hip ratio [WHR]) and allometric (a body shape index [ABSI], hip index [HI], waist to HI index [WHI]) adiposity indices are shown in **Table 1** and **Supplementary Table S1**, respectively. The mean (SD) age was 55.30 (8.02) years for the 151,526 UKBB participants and 44.70 (10.65) years for the 1,127 EHS participants. About 54% of participants were female in both populations. In the UKBB, compared to females, males were more likely to be current smokers and physically active, and had higher alcohol consumption. They were also more likely to have higher levels of adiposity indices, except for BF% and HI. In contrast, men were less likely to experience weight changes within 1 year of recruitment and to use either non-steroidal anti-inflammatory drugs (NSAID) or paracetamol (**Table 1**). UKBB participants compared to those in EHS were older and more likely to report higher alcohol consumption and physical activity, and had higher BF% and HI values, but they were less likely to smoke and had lower ABSI and WHI values.

**Table 1.**
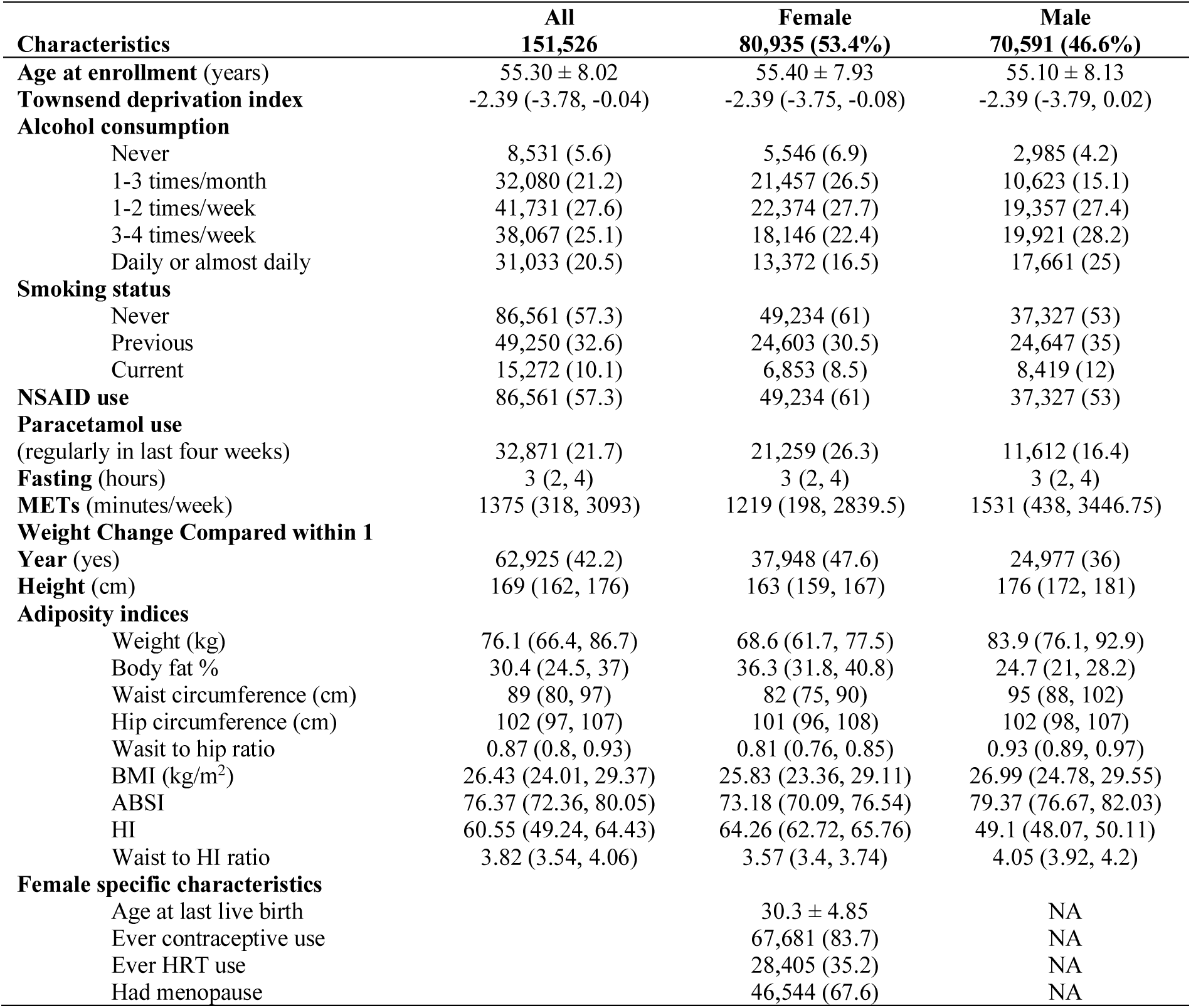
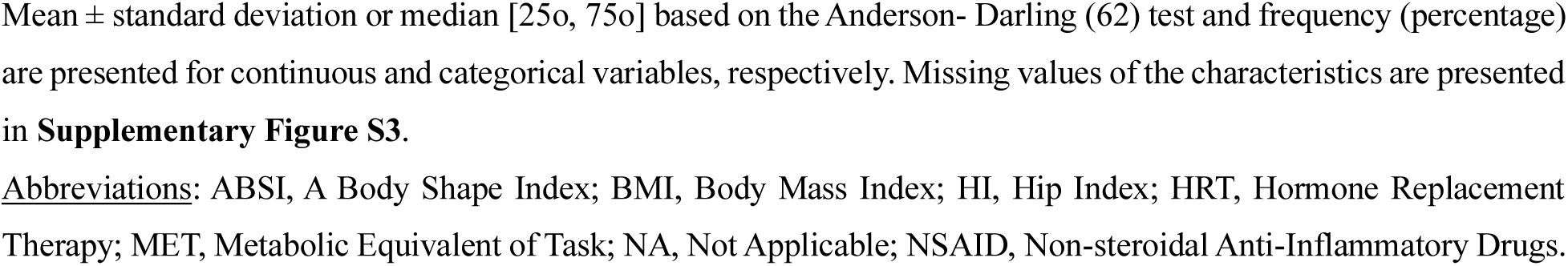
Overall and sex-specific characteristics of participants in the UK Biobank.

### Performance of the metabolic signatures

**Table 2** presents the sex-stratified performances of the final multi-step adaptive elastic net regression models (MSA-Elnet) selected for each adiposity index. For both sexes, the internal Pearson correlation coefficient (PCC) for the five non-allometric adiposity indices and their respective MSs was higher than 0.4 (range: 0.42–0.63). In contrast, for the allometric ABSI and HI, the PCC of the MSs was lower than 0.4 (range: 0.13–0.32). The highest internal PCC (females/males) was observed for MS_BF%_ (0.63/0.58) and MS_BMI_ (0.61/0.57) and the lowest for MS_HI_ (0.17/0.13). In terms of the number of metabolites composing the MSs, the median was 19.5 for females and 20.5 for males. The internal median PCCwas 0.49 for females 0.47 for males. The performance of the MSs in the EHS was consistent to the discovery population, indicating robust associations.

**Table 2.** Internal and External 1000-bootstrapped performance metrics of each metabolomic signature stratified by sex.

| Female | Internal Validation |  |  | External Validation |  |  | m |
| --- | --- | --- | --- | --- | --- | --- | --- |
|  | RMSE | Pearson | R<sup>2</sup> | RMSE | Pearson | R<sup>2</sup> |  |
| BF% | 0.87(0.86,0.88) | 0.62(0.61,0.63) | 39.05(37.81,40.31) | 0.89(0.84,0.93) | 0.61(0.57,0.64) | 36.84(32.39,41.59) | 30 |
| Waist | 0.88(0.88,0.88) | 0.61(0.6,0.62) | 37.58(36.46,38.71) | 0.95(0.91,0.99) | 0.55(0.5,0.59) | 29.78(25.01,34.96) | 25 |
| Hip | 1(1,1.01) | 0.5(0.49,0.51) | 24.88(24.24,25.53) | 1.11(1.06,1.17) | 0.38(0.32,0.44) | 14.44(10.07,19.6) | 29 |
| WHR | 1(0.99,1.01) | 0.5(0.49,0.51) | 25.25(24.39,26.13) | 1.05(1,1.10) | 0.45(0.4,0.5) | 20.23(15.61,25.46) | 8 |
| BMI | 0.88(0.87,0.88) | 0.61(0.6,0.62) | 37.75(36.54,38.97) | 0.88(0.83,0.92) | 0.61(0.57,0.66) | 37.71(32.63,43.16) | 32 |
| ABSI | 1.16(1.15,1.17) | 0.32(0.32,0.33) | 10.27(9.96,10.58) | 1.29(1.25,1.34) | 0.17(0.11,0.22) | 3.03(1.14,4.89) | 14 |
| HI | 1.29(1.28,1.30) | 0.17(0.16,0.18) | 2.93(2.68,3.2) | 1.31(1.25,1.36) | 0.14(0.08,0.21) | 1.99(0.60,4.20) | 13 |
| WHI | 1.22(1.22,1.22) | 0.25(0.24,0.26) | 6.43(5.95,6.92) | 1.23(1.19,1.28) | 0.24(0.19,0.29) | 5.66(3.47,8.39) | 8 |

| Male | Internal Validation |  |  | External Validation |  |  | m |
| --- | --- | --- | --- | --- | --- | --- | --- |
|  | RMSE | Pearson | R<sup>2</sup> | RMSE | Pearson | R<sup>2</sup> |  |
| BF% | 0.92(0.91,0.92) | 0.58(0.58,0.58) | 33.59(33.14,34.05) | 0.90(0.86,0.95) | 0.59(0.55,0.63) | 34.86(30.33,39.7) | 22 |
| Waist | 0.94(0.94,0.94) | 0.56(0.56,0.56) | 31.35(31.13,31.57) | 0.94(0.88,1) | 0.56(0.50,0.61) | 31.05(25.39,37.27) | 20 |
| Hip | 1.07(1.06,1.08) | 0.42(0.41,0.43) | 17.89(17.13,18.67) | 1.07(1.02,1.13) | 0.42(0.37,0.48) | 17.98(13.42,23.20) | 27 |
| WHR | 0.99(0.99,0.99) | 0.51(0.51,0.51) | 26.04(25.69,26.39) | 1.05(1,1.11) | 0.45(0.39,0.50) | 19.90(15.37,25.01) | 19 |
| BMI | 0.93(0.92,0.93) | 0.57(0.57,0.57) | 32.48(32.25,32.71) | 0.93(0.88,0.98) | 0.57(0.52,0.62) | 32.42(27.36,37.91) | 17 |
| ABSI | 1.22(1.22,1.22) | 0.26(0.25,0.27) | 6.54(6.05,7.06) | 1.23(1.17,1.28) | 0.24(0.18,0.30) | 5.80(3.38,8.87) | 22 |
| HI | 1.32(1.31,1.33) | 0.13(0.12,0.14) | 1.73(1.55,1.91) | 1.32(1.27,1.37) | 0.13(0.07,0.19) | 1.66(0.48,3.55) | 15 |
| WHI | 1.18(1.18,1.19) | 0.3(0.29,0.31) | 9.07(8.69,9.47) | 1.24(1.19,1.29) | 0.23(0.17,0.29) | 5.09(2.76,8.14) | 21 |

Data are presented as mean (95%CI).
**Abbreviations:** ABSI, A body shape index; BF%, body fat %; BMI, body mass index; HI, hip index; m, number of metabolites; R<sup>2</sup>, coefficient of determination; RMSE, root of mean square error; WHI, waist to hip index; WHR, waist to hip ratio.

### Metabolic signatures by adiposity indices and sex

Of the 249 metabolites analyzed by nuclear magnetic resonance (NMR), 61 and 68 metabolites were selected at least once from the MSA-Elnet for the eight adiposity MSs in females and males, respectively. These MSs (females/males) were found to mainly consist of relative lipoprotein lipid concentrations (11/17), cholesteryl esters (5/6) and fatty acids (FAs) (5/5) (**Figure 1**). In females, the adiposity indices were predominantly associated with concentrations of lipoprotein particles, apolipoproteins (Apo), fatty acids (FA), and the low-grade inflammation marker glycoprotein acetyls (GlycA), whereas in males with lipoproteins rich in cholesteryl esters, glycolysis related metabolites and aromatic/branched-chain amino acids (BCAA) (**Figure 1**). Most of the associations for both sexes among the five non-allometric adiposity indices included positive associations primarily with lipoprotein subclasses, lipoproteins particles rich in triglycerides (TG), GlycA, and FA. Inverse associations were observed with high density lipoprotein (HDL) subclasses, amino acids (AA), fluid balance metabolites and lipoproteins rich in phospholipids. Among the five non-allometric adiposity indices, differences between sexes were observed in the identified metabolites. In females, strong positive associations were observed with ApoA1, omega-6 FAs, S-HDL-P and PUFA/MUFA, and strong inverse associations with total cholines, and L-VLDL-CE. In males, positive associations were notable with L-HDL-PL, omega-6 FAs, XL-VLDL-CE and XS-VLDL-FC, whereas negative associations with L-VLDL-CE, total cholines, HDL particle sizes (average diameter) and linoleic acid (LA).

**Figure 1:**
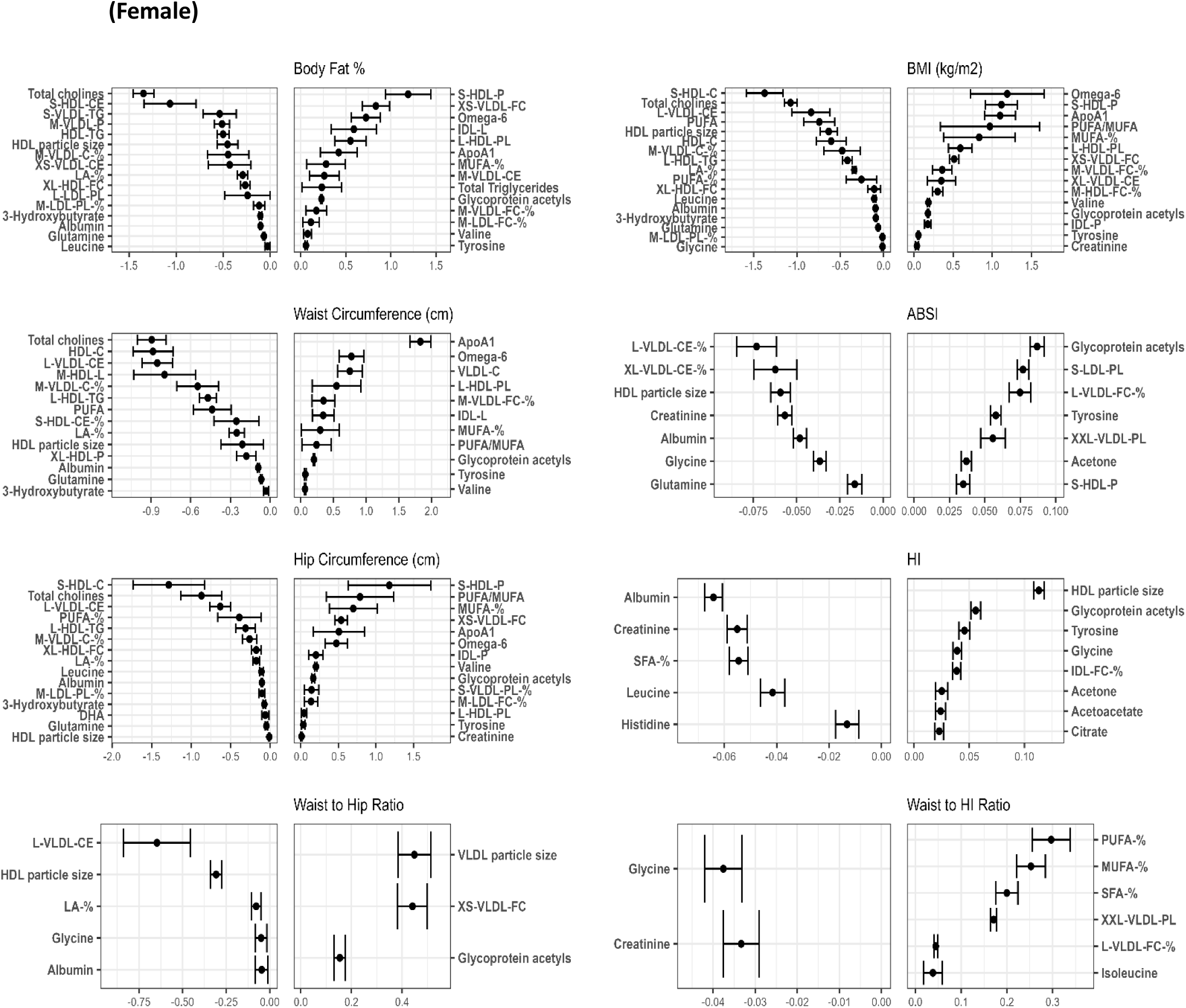
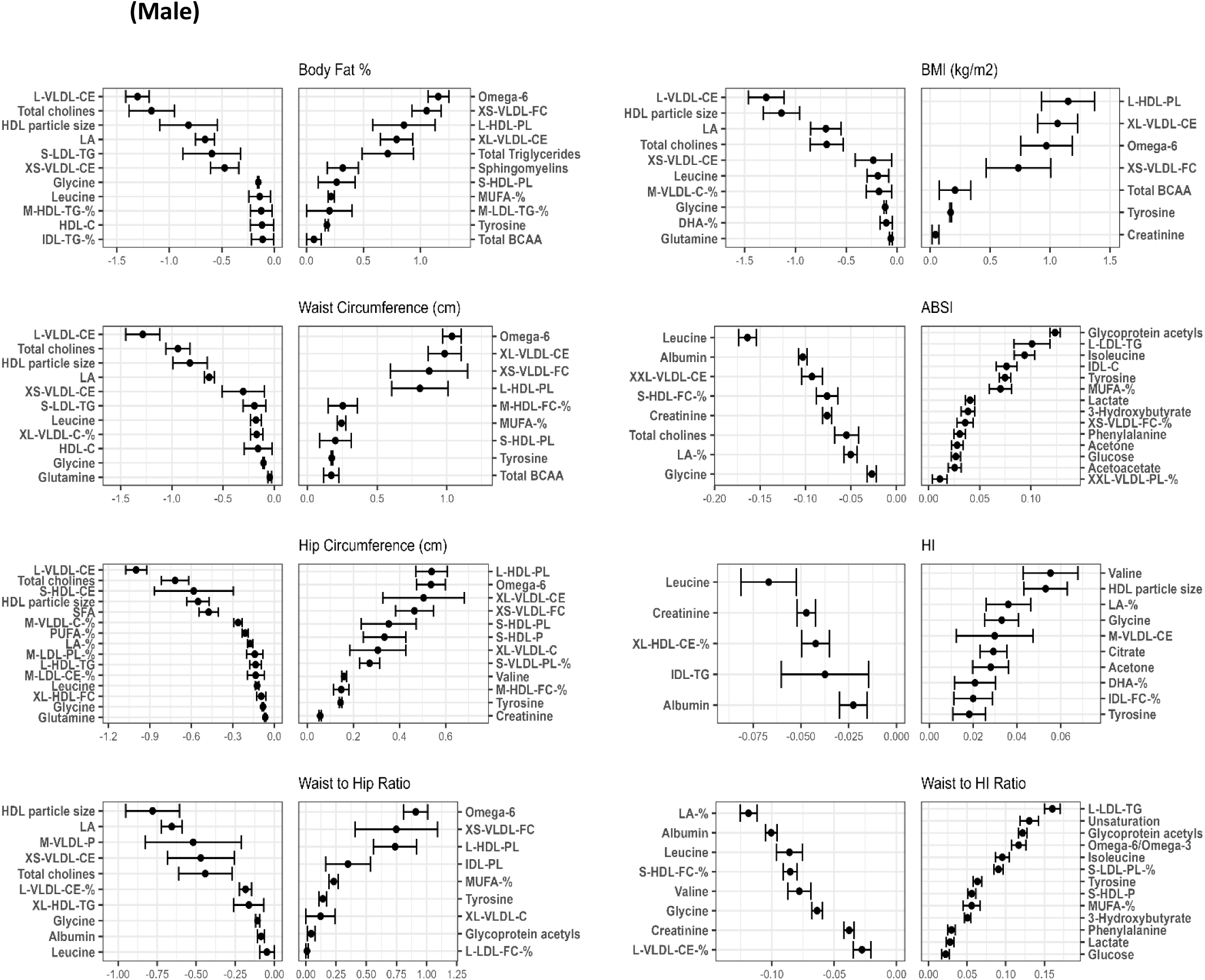
Metabolic Signatures of each adiposity index ranked from lowest to highest MSA-net negative to positive regression coefficient (bootstrap confidence intervals for the coefficients). Exposure contrast is per SD/z-score increase of the metabolite. <u>Abbreviations</u>: ABSI, a body shape index; BMI, body mass index; -C, cholesterol; HI, hip index; -TG, triglycerides; -PL, Phospholipids; -CE, Cholesteryl esters; -FC, Free cholesterol; -L, Total lipids; -P, Lipoprotein particle concentrations; XXL-, Chylomicrons and extremely large; XL-, Very large; L-, Large; M-, Medium; S-, Small; XS-Very small; LA, Linoleic Acid; DHA, docosahexaenoic acid; FA, fatty acids; HDL, high-density lipoprotein; IDL, intermediate-density lipoproteins; LDL, low-density lipoprotein; MUFA, monounsaturated fatty acid; PUFA, polyunsaturated fatty acid; SFA, saturated fatty acid; VLDL, very low-density lipoprotein; VHDL, very high-density lipoprotein.

Differences were also observed in metabolite associations and composition between allometric and non-allometric adiposity indices for each sex. In males, ABSI and WHI was associated with higher GlycA and L-LDL-TG and lower leucine and albumin. In females, ABSI was associated with higher GlycA and lower HDL particle sizes (average diameter) and L-VLDL-CE, and WHI was associated with higher PUFA% and MUFA% and inversely with glycine.

The largest differences were observed with non-allometric adiposity indices, ABSI and WHI compared to the HI. In both sexes, HDL subclasses, fluid balance and ketone bodies were positively associated with HI, and inverse associations were observed with TG, lipoprotein subclasses and fluid balance. Sex-specific variations in HI, included higher associations in females with HDL particle sizes and lower albumin, and in males with higher valine and lower albumin.

### Common and distinct metabolites in the metabolic signatures by sex

**Supplementary Figure S1** presents the overlap of MSs in females and males. On average, each index had 9.5 metabolites common to both women and men, with the lowest count being 4 metabolites for WHI. The median percentage of common metabolites was 25% (range: 16%–51%). There was a larger number of distinct metabolites in women compared to men for BF, BMI, WC, and HC, a similar number for HI, and a considerably lower number than men for WHR, WHI and ABSI.

### Common and distinct metabolites in the metabolic signatures by adiposity index

We identified 25 and 31 distinct metabolites among all MSs in females and males, respectively (**Supplementary Table S2**), with differences in their classes and counts. In females/males, most of distinct metabolites were relative lipoprotein lipid concentrations (6/13), lipoproteins rich in: TG (3/4), cholesteryl esters (4/3) and AA (2/0). Across all signatures, GlycA, albumin and HDL particle size (average diameter) were the common metabolites in females (except WHI), whereas glycine, tyrosine and leucine were common in males. GlycA and albumin had positive and negative coefficients, respectively, while HDL particle size (average diameter) had positive coefficients only for HI. Positive and inverse associations were observed in tyrosine and leucine, and glycine was positively associated only with HI. (**Supplementary Table S2**).

Furthermore, we examined pairwise overlap between the MSs of adiposity indices by sex (**Supplementary table S3**). Overall, the mean percentage of the common metabolites was 24% (range: 2.7%–74.3%) in females, and 27.1% (range: 10.3%–68%) in males. In both sexes, the highest percentages of common metabolites were observed between non-allometric adiposity indices (median: 42.4%; range: 9%–56%). Conversely, comparisons between the allometric and non-allometric MSs revealed the lowest counts of common metabolites.

### Associations of the adiposity indices and corresponding metabolic signatures with clinical Biomarkers by sex

The sex-specific associations of the eight MSs of the adiposity indices with 29 clinical biomarkers are shown in **Figure 2**, before and after adjusting for the respective phenotypic adiposity indices with minor differences on the effect estimates between the two models. **Supplementary Table S4** presents the descriptive statistics for the biomarkers. There were only a few qualitative differences by sex for testosterone (inverse in males, null in females) with MS_ABSI_, MS_HI_ and MS_WHR_; urea (inverse in males, positive in females) with M_ABSI_; alkaline phosphatase (ALP) (inverse in males, positive in females) with M_HI_; lipoprotein-a (inverse in females, null in males) with MS_ABSI_. There were many quantitative differences in associations by sex (**Supplementary Table S5-S6**) with the strongest differences to be observed for testosterone (stronger inverse in males), c-reactive protein (CRP) (stronger positive in females), ALP (stronger positive in females) and alanine transaminase (ALT) (stronger positive in males) concentrations with MS_BF%_, MS_BMI_, MS_WC_ and MS_HC_; TG (stronger positive in females), LDL (stronger positive in females), HDL (stronger inverse in females) concentrations with MS_ABSI_ and MS_WHR_; testosterone (stronger positive in males), TG (stronger inverse in males), ALT (stronger inverse in males) with MS_HI_; TG (stronger positive in females), testosterone (stronger inverse in males) and CRP (stronger positive in males) with MS_WHI_.

**Figure 2.**
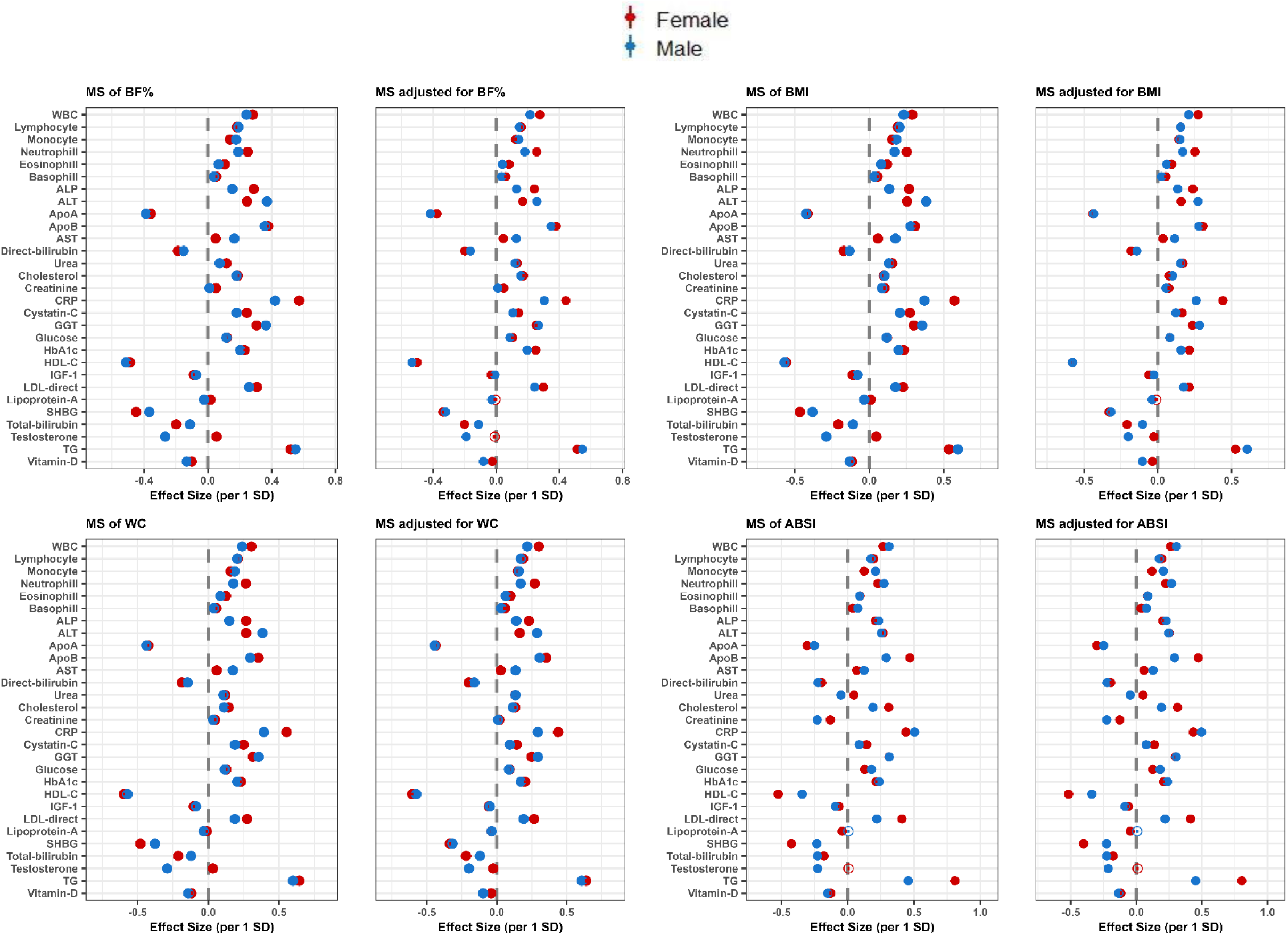
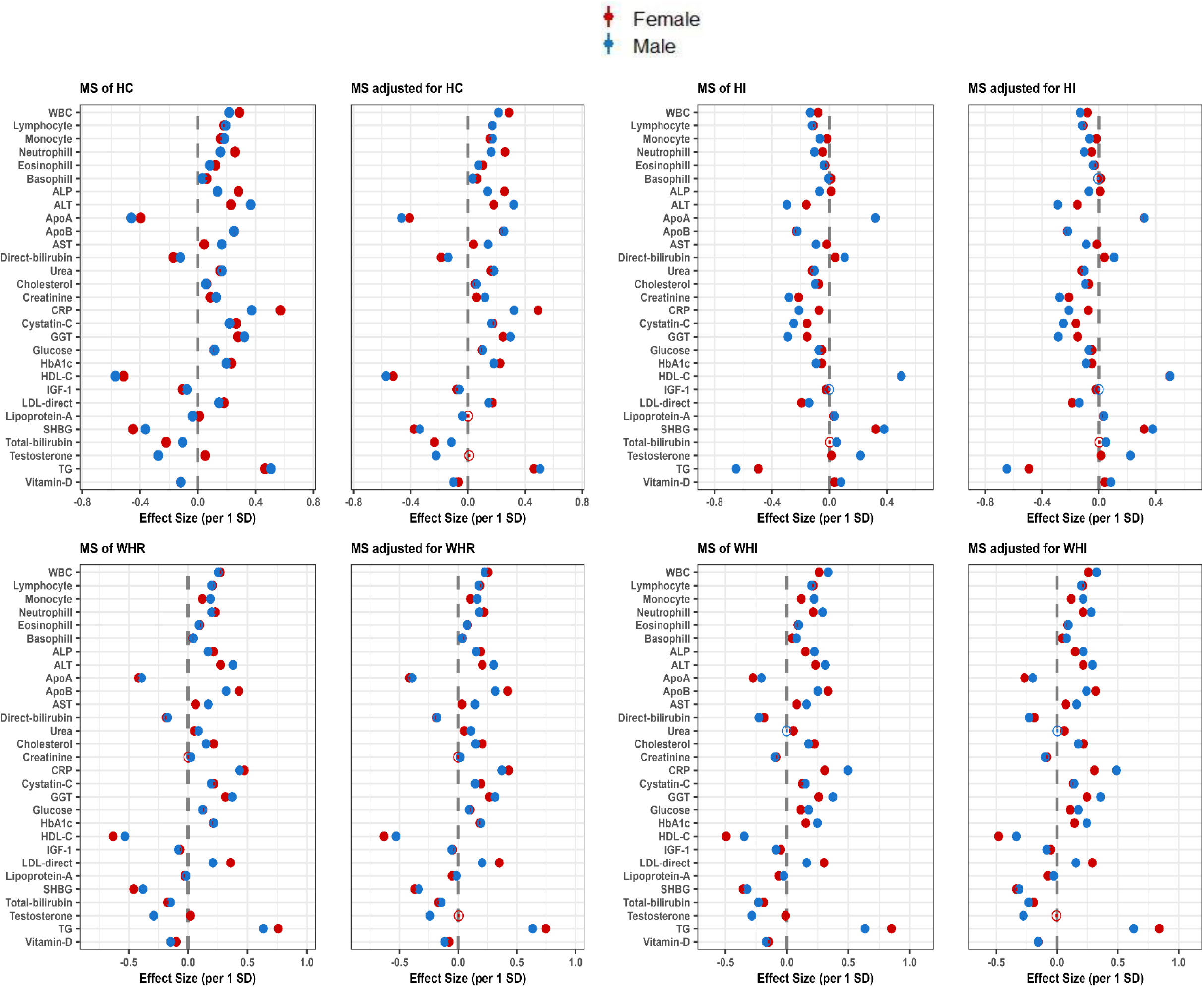
Sex-specific associations of standardized MSs and standardized MSs adjusted for the corresponding adiposity index with 29 clinical biomarkers. Multivariable-adjusted model, based on age, alcohol consumption, smoking status, metabolic equivalent of task, townsend deprivation index, paracetamol use, NSAID, weight change, fasting time, and in females also menopausal status, oral contraceptives use and HRT. Females and males are presented with red and blue colors, respectively. Coefficients are expressed as 1/SD increase. <u>Abbreviations</u>: ABSI, a body shape index; ALP, alkaline phosphatase; ALT, alanine aminotransferase; Apo, apolipoprotein; AST, aspartate aminotransferase; BF%; body fat %; BMI, body mass index; CRP, C-Reactive protein; GGT, gamma glutamyltransferase; HbA1c, glycated hemoglobin; Hip, hip circumference; HI, hip index; MS, metabolic signature; TG, triglycerides; WHR, waist to hip ratio; WBC, white blood cells.

### Associations of the adiposity indices and corresponding metabolic signatures with clinical biomarkers by adiposity index

The adiposity-specific associations of the eight MSs with 29 clinical biomarkers by sex are shown in **Figure 3**. In both sexes, the associations revealed that MS_HI_ qualitatively differed from the other MSs in most biomarker associations. The association estimates in both sexes were more pronounced for TG, HDL-C, CRP, ApoA, ApoB, LDL and sex hormone binding globulin (SHBG) regardless of adiposity index. **Supplementary Table S6** extends findings presented in **Figures 2 and 3** by also providing results for the associations of phenotypic adiposity indices before and after adjustment for the respective MSs with each clinical biomarker. Associations between adiposity indices and biomarkers were attenuated and, in some cases, became insignificant after further adjustment for respective MS.

**Figure 3.**
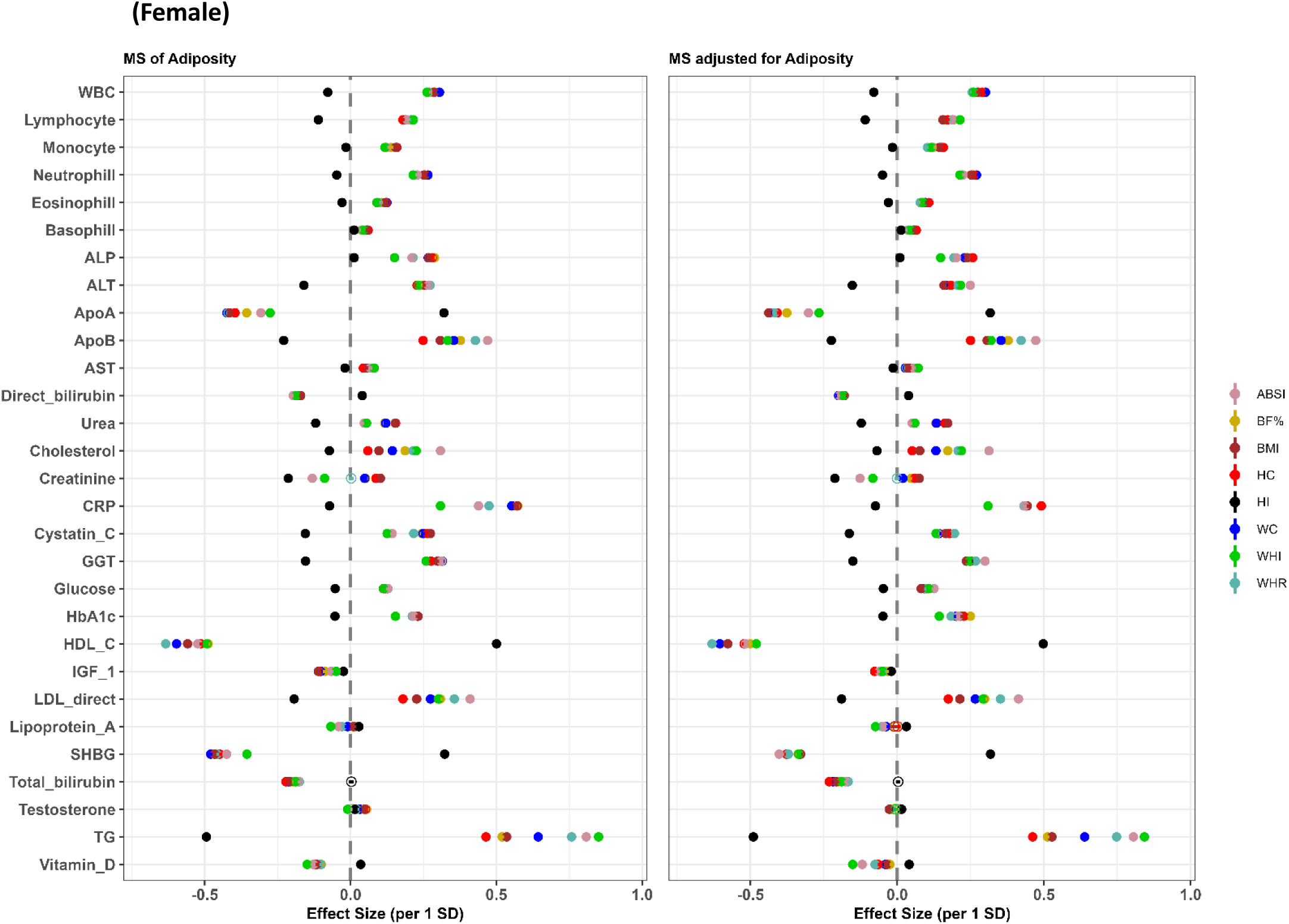
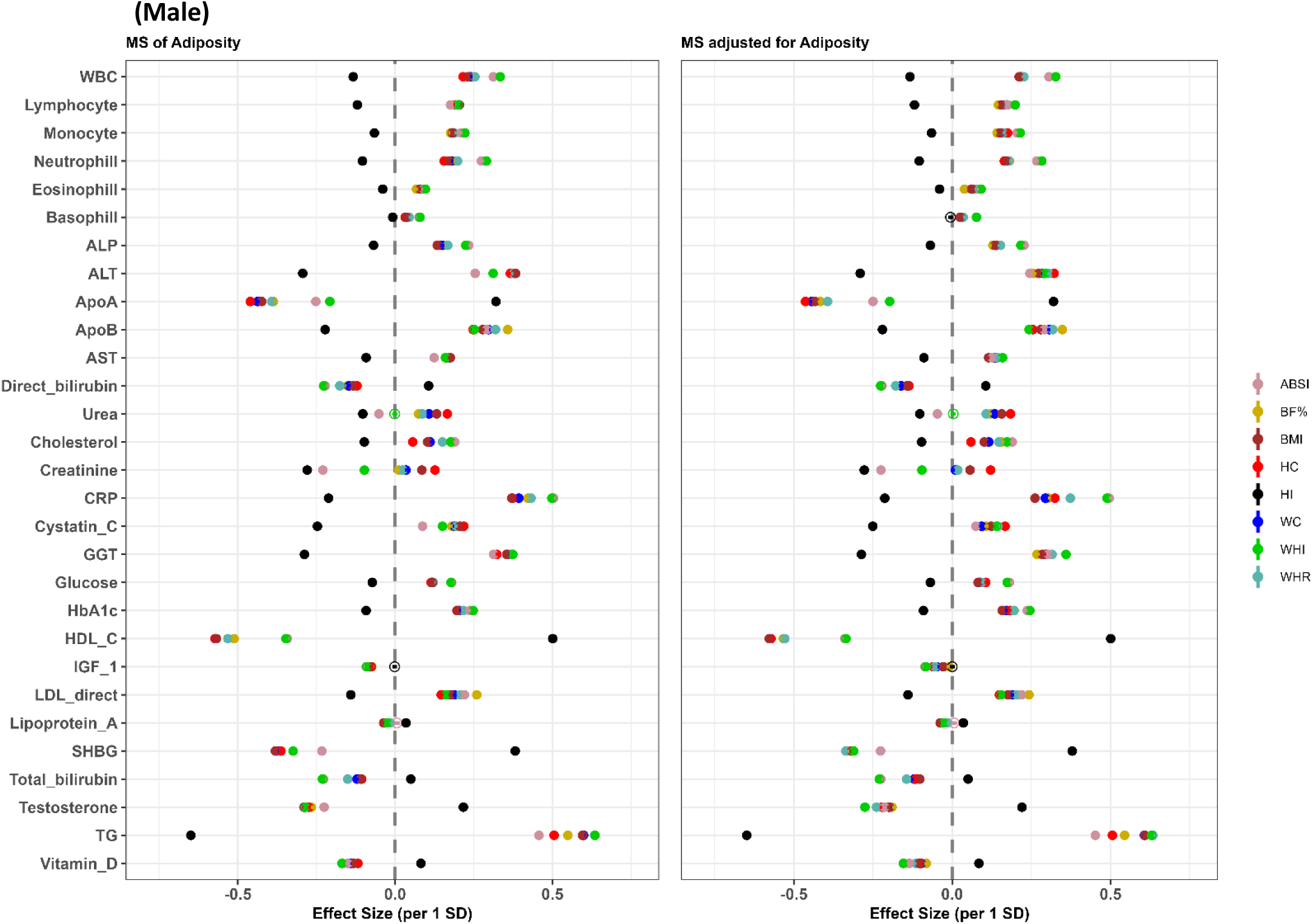
Associations of standardized MSs and standardized MSs adjusted for the corresponding adiposity index with 29 clinical biomarkers, stratified by sex. Multivariable-adjusted model, based on age, alcohol consumption, smoking status, metabolic equivalent of task, townsend deprivation index, paracetamol use, NSAID, weight change, fasting time, and in females also menopausal status, oral contraceptives use and HRT. Coefficients are expressed as 1/SD increase. <u>Abbreviations</u>: ABSI, a body shape index; ALP, alkaline phosphatase; ALT, alanine aminotransferase; Apo, apolipoprotein; AST, aspartate aminotransferase; BF%; body fat %; BMI, body mass index; CRP, C-Reactive protein; GGT, gamma glutamyltransferase; HbA1c, glycated hemoglobin; HC, hip circumference; HI, hip index; MS, metabolic signature; TG, triglycerides; WC, waist circumference; WHI, waist to hip index; WHR, waist to hip ratio; WBC, white blood cells.

## Discussion

Leveraging metabolomics data from 151,526 healthy UKBB participants, we identified sex-specific MSs of eight adiposity indices and validated them in an external population. Our analysis revealed notably sexual dimorphism in the metabolism of adiposity indices, with a median percentage of common metabolites between men and women of 25%. The non-allometric MS_BMI_, MS_BF%_, MS_WC_, MS_HC_, and MS_WHR_ had the highest overlap in metabolites, whereas the allometric MS_ABSI_, MS_HI_ and MS_WHI_ had the highest percentage of distinct metabolites compared to the non-allometric MSs. This study also revealed that the identified MSs were independently associated with over 25 biomarkers with differences observed by sex and adiposity index, and the strength of these associations was greater compared to the associations with the respective adiposity indices. Specifically, the associations of MS_HI_ with most biomarkers differed qualitatively compared to the rest of the MS associations.,

The relationship between blood metabolome and adiposity indices has been extensively investigated. Several observational studies using NMR or mass spectrometry, combined with Mendelian randomization or univariate associations, have identified numerous circulating metabolites associated with adiposity indices (13, 23–26). These metabolites were mostly lipoprotein particles of different sizes and subclass concentrations, ketone bodies, fatty acids, hexoses, glycerophospholipids, glycerolipids, biogenic amines, sphingolipids, and phosphatidylcholine species (13, 23–26). A recent cohort of 30,000 healthy Mexicans performed a sex-specific metabolome-wide association study to associate 139 NMR metabolites with four adiposity indices (BMI, WC, HC, WHR) (13). They found total cholines, L-VLDL-CE and LA positively, whereas ApoA1 and L-HDL-Pl negatively associated with these adiposity indices and did not detect sexual dimorphism. Furthermore, a Finnish study of 12,664 healthy adults, investigated the association between 82 NMR metabolites and BMI using Mendelian randomization (21). This study showed positive associations with leucine and PUFA, while inverse associations with ApoA1 and, similarly, did not detect sexual dimorphism.

Our study contradicts some of these findings, indicating opposite directions for certain metabolites, such as very large and large VLDL rich in CE, and identifying sexual dimorphism in adiposity metabolism. Beyond population-specific characteristics and lifestyle factors (e.g. dietary habits) that could have affected metabolite concentrations, these previous studies used statistical techniques that could not control for the high multicollinearity in metabolomics data and associations independencies between metabolites, and these reasons might explain these discrepancies. To the best of our knowledge, this is the first study to identify MSs of the eight adiposity indices using a multi-step estimation algorithm built upon adaptive elastic-net regularization with stability selection, reducing false positives and improving the stability of feature selection, while maintaining estimation accuracy (27). Furthermore, we externally validated the identified signatures enhancing reproducibility and minimizing type I errors, indicating their potential as biomarkers for adiposity.

Besides traditional adiposity indices, we identified MSs for ABSI, HI and WHI that have never been explored before. WC, HC and WHR are dependent on overall body size, while ABSI, HI and WHI are measures of abdominal and gluteofemoral fat, respectively, and are independent of weight and height. The MSs of these allometric exhibited a high percentage of distinct metabolites and differed in composition and association with biomarkers compared to those of WC, HC and WHR. MS_ABSI_ was associated with a greater atherogenic profile, characterized by higher concentrations of IDL-C, TG-rich LDL particles and inflammatory GlycA in males, and higher concentrations of lipoprotein particles rich in TG such as LDL and VLDL and GlycA in females. On the other hand, several studies have reported that a greater fat accumulation in the hips might be associated with a more favourable metabolic profile (28, 29). Our results support these findings and further demonstrate sex-dependent differences in the metabolomic profile of HI. In females, HI was associated with higher concentrations of HDL particles and lower concentrations of saturated fatty acids. In males, MS_HI_ consisted of a higher number of HDL subclass particles, and higher concentrations of valine, while lower concentrations of TG-rich IDL particles.

In our study, the amino acids glycine, leucine and tyrosine were consistently present across all MSs in males. In females, tyrosine was included in six MSs, glycine in five, and leucine was present in four. Leucine was inversely associated with the adiposity indices, a finding that agrees with a previous metabolomic investigation (30) in Spanish and Danish participants with overweight/obesity. In contrast, studies involving US, Finnish, and Mexican populations found elevated circulating leucine concentrations associated with obesity (21, 22, 31). Further research is needed to understand the tissue-specific BCAA oxidation dynamics in obesity (32). Tyrosine was positively associated with adiposity indices, consistent with previous reports (23, 33). Furthermore, tyrosine has been linked to an increased risk of T2D (34, 35) and cardiovascular diseases (36).

In previous cross-sectional studies of adults with and without obesity, GlycA concentrations were positively associated with obesity. Similarly, in our study, GlycA was the only common inflammatory marker positively associated with all adiposity indices in females, and with WHR, ABSI and WHI in males. Based on these findings, GlycA may better reflect inflammatory processes that are driving adipocyte dysfunction in females, whereas in males, abdominal fat accumulation may promote systemic inflammation. Previous studies have demonstrated the utility of GlycA for predicting incident cardiovascular diseases (37), type 2 diabetes (38), cancer (39) and Alzheimer’s disease (40).

Although, numerous studies have investigated the relationship between traditional adiposity indices and clinical biomarkers, few have explored ABSI, HI and WHI (41–43). Positive associations have been reported between ABSI, BMI and TG, fasting glucose, and inverse associations for HDL-C in a European adult population (42). In Chinese adults, positive associations of BMI and WC with ALT and gamma-glutamyl transferase were found (44). Higher ABSI and BMI values have also been associated with higher CRP levels (45). Previous research using UKBB data revealed opposite directions between HI vs. BMI and HI vs. ABSI associations with liver, metabolic and inflammatory related biomarkers (43). Our results are in line with these findings and expand on them by examining the MS of adiposity indices, revealing stronger associations with most clinical biomarkers compared to the associations of phenotypic adiposity indices. Notably, the MSs of ABSI and in particular HI, showed stronger associations in most biomarkers compared to the respective non-allometric adiposity indices. This suggests that the MS of these allometric adiposity indices may capture residual effects of adiposity indices along with other metabolic influences. The largest magnitude of associations was observed between all MSs and TG, HDL-C, ApoA, ApoB, SHBG, CRP, and LDL. This might be because MSs were constructed based on an NMR platform that mainly measures atherogenic lipid metabolites (46). The stronger associations of MS_ABSI_ with most of liver and inflammatory related biomarkers in males, and metabolic-related biomarkers in females, compared to MS_BMI_ and MS_WC_, confirm its more atherogenic profile. On the other hand, MS_HI_ showed favourable associations compared to other MSs of overall and abdominal adiposity, supporting its health benefits. Our findings provide novel evidence by shedding light on associations between adiposity indices and biomarkers relevant to a spectrum of chronic diseases, such as cardiovascular, metabolic, renal, and liver conditions.

To the best of our knowledge, this study represents the first comprehensive assessment of NMR-based metabolites associated with eight adiposity indices and extends the analyses to include an atlas of associations with clinical biomarkers in a large population. The inclusion of an external population strengthens the robustness and generalizability of our MS findings. Many exclusion criteria such as comorbidities and medication use were applied to minimize their effects on metabolites’ and biomarkers’ concentrations. There are also limitations in this study. First, the NMR platform identified metabolites mainly involved in lipid metabolism. Other techniques like mass spectrometry and untargeted approaches could advance our understanding of the metabolic effect of adiposity on health outcomes. Second, for interpretation purposes, we employed a machine learning model that assumes linear associations between adiposity indices and metabolites and due to the inability to assess interactions among metabolites, some information may have been lost potentially affecting its performance. Third, the human metabolome is dynamic and constantly in flux, necessitating long-term repeated metabolomics data to understand how changes in metabolites at different time points affect health outcomes.

## Conclusions

In summary, our findings suggest that different regions of the adipose tissue undergo distinct metabolic processes, each having a unique impact on health. We observed differences in the composition of the MS of different adiposity indices by sex. Comparisons between the allometric and non-allometric MSs revealed the lowest counts of common metabolites in both sexes. GlycA, albumin and HDL particle sizes were the only common metabolites associated with all adiposity indices except with WHI in females, whereas in males glycine, tyrosine and leucine were common in all MSs. Furthermore, MS_HI_ qualitatively differed from the rest of MSs in most biomarkers associations. Based on the metabolite content of different MSs by adiposity index and their associations with biomarkers, MS_ABSI_ was found to be more atherogenic, whereas MS_HI_ was more favourable for health. In addition, MSs showed stronger associations with clinical biomarkers compared to respective associations with traditional adiposity indices, highlighting the importance of considering metabolic factors beyond simple measures of adiposity in assessing health risk.

## Methods

### Study design and populations

#### Discovery population

This is a cross-sectional study leveraging data from the UKBB. UKBB is a population-based, multicenter prospective cohort study comprising 502,359 participants aged 37 to 73 years old living in the UK, recruited between 2006 and 2010 (47). Metabolites in plasma (46) were measured in 248,286 participants using NMR provided by the Nightingales’ Health platform. Exclusions were defined based on previous literature (**Figure 4**) (43). We excluded participants with self-reported non-white ancestry (N=12,841) due to insufficient numbers from other ethnicities. Additionally, participants with missing data on any adiposity index or extreme anthropometric measurements (height <130 cm; WC <50 or >160 cm; BMI <18.5 or ≥45 kg/m^2^) were excluded (N=6,902). We further excluded participants with mismatched self-reported genetic sex or aneuploidy (N=408) and pregnant females at enrollment (N=1). To minimize the impact of comorbidities and medications on metabolites’ concentrations, we excluded participants with self-reported prevalent cancer at enrolment or prevalent date of cancer diagnosis from baseline (N=21,464), those who developed cancer or died within two years after enrollment (N=4,943), those with self-reported endocrine (N=410), thyroid (N=11,461) and liver-related diseases or kidney failure (N=1,662) at enrollment, diabetes or received anti-diabetic medication (N=8,660), inflammatory bowel disease (N=1,551), chronic respiratory illness or heart failure (N=4,857). Participants who were using lipid-lowering medications or exogenous glucocorticoids at the time of enrollment were also excluded (N=21,499) (**Supplementary Text S1**). Finally, the study included 151,526 participants (80,935 females and 70,591 males).

**Figure 4.**
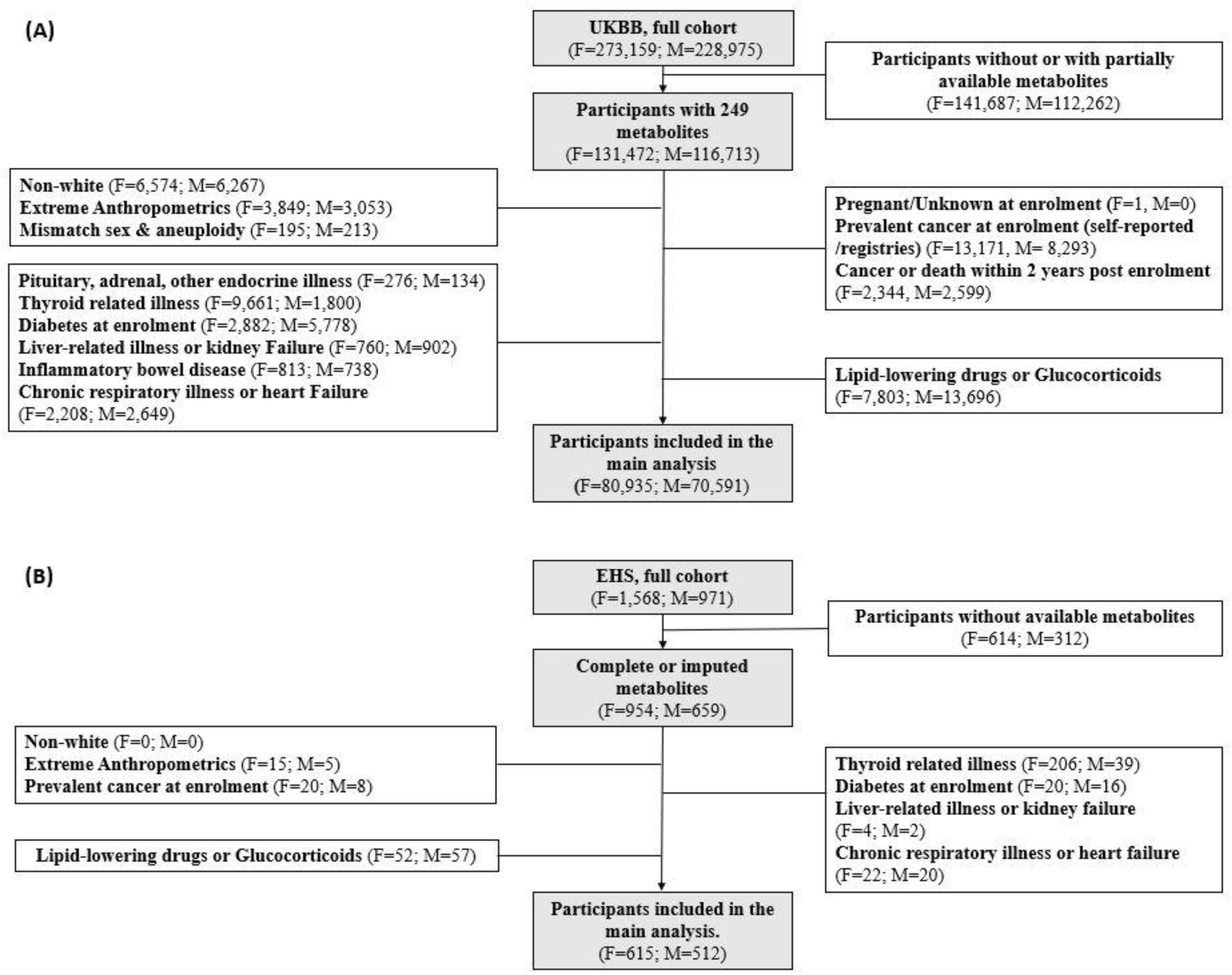
Flowchart of the UK Biobank (A) and Epirus Health Study (B). Boxes with dark color represent included participants, while boxes with white color represent excluded participants. <u>Abbreviations</u>: ABSI, a body shape index; BMI, body mass index; EHS, Epirus Health Study; F, females; HI, hip index; M, males; UKBB, UK Biobank; WC, waist circumference.

#### External validation population

We sought to validate the associations between MS and adiposity indices from the UKBB in the Epirus Health study (EHS) (48–50). EHS recruited over 2,500 participants aged 20 to 74 years old from 2019 to 2023 where 1,613 had available metabolomic information (**Figure 4**). Similar to the UKBB exclusions, we removed participants with missing or extreme anthropometric measurements (N=20), participants with prevalent cancer at enrollment (N=28), those with thyroid (N=245), diabetes (N=36), liver-related diseases or kidney failure (N=6), chronic respiratory illness or heart failure (N=42), and participants who were receiving lipid-lowering medications or exogenous glucocorticoids at enrollment (N=109). This resulted in a cohort of 1,127 participants (615 females and 512 males).

## Measurements

### Metabolomic profiling

#### Discovery population

Metabolomic data were analyzed in ethylenediaminetetraacetic acid (EDTA) plasma samples generally drawn non-fasting participants, with a median of 3 hours since the last meal (**Table 1**). They were quantified using Nightingale Health’s high-throughput proton NMR (1H-NMR), which allows for the simultaneous quantification of 168 circulating metabolic measures. Apart from the 168 directly quantified metabolites, 81 ratio measures were also computed, providing a comprehensive targeted analysis of 249 metabolites (**Supplementary Text S2**). These measures encompass lipids, lipoprotein subclass, fatty acid composition, and various low-molecular-weight metabolites, such as amino acids, ketone bodies, and glycolysis metabolites, quantified in molar concentration units (51).

#### External validation population

Metabolomic analyses were performed on 8-hour fasting plasma EDTA samples. These samples were analyzed using the same Nightingale Health’s NMR-based metabolomic profiling platform as in the UKBB (52). Descriptive statistics for the metabolomics data in UKBB and EHS are presented in **Supplementary Table S7**.

### Adiposity indices

Adiposity indices, including BF%, BMI (kg/m^2^), HC (cm), WC (cm), and WHR, were used for the estimation of total body, gluteofemoral, and abdominal fat mass. We also computed ABSI, HI and WHI allometric indices, to address the correlation of traditional abdominal and gluteofemoral indices with BMI using coefficients from the National Health and Nutrition Examination Survey (NHANES) (53, 54). HI calculated with coefficients from NHANES was uncorrelated with BMI in women, but was inversely correlated with BMI in men probably due to black ethnic participants in NHANES (54). To calculate HI for men, we used simple-fraction coefficients based on UK Biobank data (7, 55). **Supplementary Figure S2** presents the correlations among the study’s adiposity indices. The assessment methods for adiposity indices in both studies are presented in **Supplementary Table S8**.

### Covariates

Extensive health information was gathered for each participant in UKBB through interviews, primarily in the form of questionnaires. This information covered various aspects, including demographics (age, sex), socio-economic (Townsend deprivation index [TDI]) and lifestyle factors (smoking status, alcohol consumption, energy expenditure), anthropometric (weight change within the last year preceding enrollment), recent medication use (paracetamol, NSAID), fasting time and for females also menopausal status, ever oral contraceptives use, and ever hormone replacement therapy use (HRT). Biological data from blood samples were collected and subjected to a well-described handling and storage protocol (56). **Supplementary Figure S3** presents the percentages of UKBB covariates missing values (median = 0%; range 0%–15%).

### Biomarkers and clinical variables

Blood samples were collected from participants in UKBB at various times throughout the day, from 8 a.m. to 9 p.m. There were no specific requirements for fasting prior to sampling. These samples were used to measure health-related biomarkers. We selected 29 biomarkers linked to cardiometabolic, endocrine, bone, liver and renal health. Among these, 28 biomarkers were measured in serum (SST tube), and one (HbA1c) in red blood cells (EDTA tube). Detailed information on assay performance can be found elsewhere (57, 58). **Supplementary Figure S4** presents the percentages of missing values for these biomarkers (median: 4.6%; range: 2.5%–22.9%).

### Ethical approval

Both UKBB and EHS studies underwent ethical review and were approved by the Northwest Multi-Centre Research Ethics Committee and the Research Ethics Committee of the University of Ioannina, respectively. All participants provided written informed consent before participation.

### Statistical analyses

#### Metabolic data and missing values

In the discovery population, we used the complete case metabolomic data without excluding metabolites, as the median percentage of missingness was 0% (range: 0%–4.3%). In the external validation population, one of the 249 plasma metabolites, namely β-hydroxybutyrate, was removed due to the high number of missing values (>20%). The remaining 248 metabolites had a median percentage of missingness of 1% (range: 0%–8.8%). To avoid loss of power in EHS, missing values of these metabolites were sex-stratified imputed using the non-parametric random imputation method (missForest) based on a random forest (59, 60). **Supplementary Figure S5** presents the sex-specific distributions of observed and imputed metabolites that had the highest percentage of missingness compared to others. Only the metabolite data were used as variables for imputation. The rank-based inverse normal transformation was applied to metabolites in each study to distribute their weights evenly and approximate normality. During the validation phase, β-hydroxybutyrate was omitted from the identified MSs, to ensure consistency in metabolomics data across both studies.

#### Identification of metabolic signatures correlated with adiposity indices

We regressed each adiposity index on 249 metabolites to identify the MSs stratified by sex. Prior to regression, adiposity indices were standardized into z-scores. **Supplementary Figure S6** presents adiposity indices distributions. Due to the high dimensionality and multicollinearity of the metabolomics data, we used the MSA-Elnet (27), an iterative procedure of Adaptive Elastic Net, which is a combination of Elastic Net and Adaptive Lasso. It performs different amounts of shrinkage to different features, aiming to penalize the smaller coefficients more severely. Feature selection was performed utilizing stability selection of 100 iterations keeping a subset of identified metabolites >60 times (61). We then split the dataset into train (80%) and test (20%) sets and applied grid searching with five iterations of the algorithm, tuning our model for the 1) hyperparameters L1 and L2 norms, for 2) both minimum and 1 standard errors metrics, and 3) the positive constant gamma which defines the data-driven weighting parameter (0.1,0.5,1,1.5,1.8,2,2.5,3), using 10-fold cross-validation (CV) techniques (**Supplementary Figure S7**). Our criteria to choose the optimal combination was the 1000-bootstrapped root of mean square error (RMSE). SE of coefficients derived after 100-bootstrap samples of the final coefficients. Metabolic scores were constructed as the weighted sum of the coefficients for each MS separately. To assess both internal and external validity, MSs were standardized in each population based on MS z-scores derived from the UKBB. The 1000-bootstrapped PCC was calculated to evaluate the performance of the identified metabolic signatures. We then applied the discovery model to the external validation set to evaluate the performance of each MS using the bootstrapped RMSE and PCC.

#### Common and distinct metabolomics

Across all adiposity indices, we identified common and distinct sex-specific metabolites derived from MS, comparing them pairwise or simultaneously. Sex- and adiposity index-specific Venn diagrams were created to visualize these findings.

#### Associations of the metabolic signatures with clinical biomarkers

We examined the associations of the standardized MSs, the respective standardized adiposity indices and their mutual adjustments with each biomarker fitting four multivariable linear regression models stratified by sex. Each model was adjusted for age at enrollment (continuous), alcohol consumption (categorical), smoking status (categorical), energy expenditure measured in metabolic equivalents of task (METs; continuous), TDI (continuous), paracetamol use regularly in last four weeks (binary), NSAID (binary), weight change within the last year preceding enrollment (binary), fasting time (continuous), and in females also menopausal status (binary), oral contraceptives use (binary), and HRT (binary). More information regarding covariates categories can be found in **Supplementary Text S3**. All biomarker measurements were log-transformed (natural logarithm log_e_) to mitigate the influence of right-skewed distributions. To ensure meaningful comparisons, all adiposity indices, MS and biomarkers were standardized into z-scores stratified by sex. To assess heterogeneity between sexes for each adiposity measure in biomarker associations, we computed the Cochran’s Q test. To account for multiple testing, we adjusted p-values in all associations using Bonferroni’s correction (0.05/number of biomarkers). All analyses were performed using R version 4.4.0.

## Acknowledgement

This research has been conducted using the UK Biobank Resource under Application Number 79696.

## Data availability

All data used in this work derived from UK Biobank Resource under Application Number 79696.

## Author contributions

Conceptualization: CKP, CP, KKT

Methodology: CKP, KKT

Validation: CKP, KKT

Formal analysis: CKP, GM

Resources: CP, KKT

Writing - Original Draft: CKP, CP

Visualization: CKP

Writing - Review & Editing: All

Supervision: CP, KKT

Funding acquisition: KKT, RMM

## Disclaimer

Where authors are identified as personnel of the International Agency for Research on Cancer / World Health Organization, the authors alone are responsible for the views expressed in this article and they do not necessarily represent the decisions, policy, or views of the International Agency for Research on Cancer / World Health Organization.

