## Supplementary Files for "Sex-specific metabolic signatures of adiposity associated with clinical biomarkers in the UK Biobank"

**Affiliations**

*: Shared last authorship

[†]**Corresponding author**

*Christos Papagiannopoulos, MSc, PhDc, Department of Hygiene and Epidemiology, School of Medicine, University of Ioannina, Ioannina, Greece, T.Th.1186, Campus Ioannina University, Zip code: 451 10

### Supplementary Text Information

**Supplementary Text S1**. Exclusion criteria in UK Biobank population.
**Supplementary Text S2**. Metabolic biomarker measurements by Nuclear Magnetic Resonance.
**Supplementary Text S3.** Definition of covariates.

### Supplementary Table Descriptions

**Supplementary Table S1.** Overall and sex-specific baseline characteristics of Epirus Health Study.
**Supplementary Table S2**. Overlapped metabolites of the eight metabolic signatures. Coefficients expressed per 1 SD increment of respective metabolite, sorted from most common to most distinct across metabolic signatures. With pink color presented the most common, and orange the most distinct.
**Supplementary Table S3**. Overlapped metabolites of the eight metabolic signatures of body adiposity indices in pairs and by sex in the form of Y/X/C (X; Distinct in vertical, Y; Distinct in horizontal; C, Common).
**Supplementary Table S4**. Overall and sex-specific descriptives of the clinical biomarkers in the UK Biobank.
**Supplementary Table S5**. P-values from q-test for heterogeneity between sexes for each metabolic signature and biomarker. Bonferroni correction was performed (0.05/number of biomarkers).
**Supplementary Table S6.** Sex-specific associations of the standardized adiposity indices, the respective standardized metabolic signatures and the associations of their mutual adjustments with 29 clinical biomarkers in z-score. Models are adjusted for age, alcohol consumption, smoking status, metabolic equivalent of task, town deprivation index, paracetamol use, NSAID, weight change, fasting time, and in females also menopausal status, oral contraceptives use and HRT. Coefficients are expressed as 1/SD increase.
**Supplementary Table S7**. Overall and sex-specific descriptives in UK Biobank and Epirus Health Study for 249 nuclear magnetic resonance metabolites. Data are presented as Mean ± SD and Median (25o, 75o). Abbreviations: IQR; interquartile range; SD, standard deviation.
**Supplementary Table S8**. General information and assessment of adiposity indices in UK Biobank and Epirus Health Study

### Supplementary Figure Legends

**Supplementary Figure S1.** Overlaps of the metabolic signatures between females (red) and males (light blue).
**Supplementary Figure S2.** Pearson correlation coefficients between eight body adiposity indices by sex. Green and brown colors present higher or lower correlations, respectively. Color intensity presents the magnitude of the association.
**Supplementary Figure S3.** Missing values percentages of the covariates used in analyses
**Supplementary Figure S4.** Missing values percentages of the UK Biobank clinical biomarkers.
**Supplementary Figure S5.** Sex-specific distributions of observed and imputed nuclear magnetic resonance metabolites in the Epirus Health Study. Metabolites that are presented had the highest percentage of missingness compared to others.
**Supplementary Figure S6.** Normal and log-transformed distributions for each body adiposity index by sex in UK Biobank. Red and blue colors present the normal and log-transformed distributions, respectively.
**Supplementary Figure S7**. Hyperparameter tuning and selection of the sex-stratified MSA-net for all adiposity indices. Red and blue colors present lambda min and lambda 1SE criterion, respectively. Values in circles present the number of identified metabolites after selecting the best combination of L1 and L2 norms. Colored boxes present the respective parameter selection of gamma, and lambda criterion.

### Supplementary Text

**Supplementary Text S1.** Exclusion criteria in UK Biobank population**.**

**The exclusion criteria were applied sequentially in the following order, with each participant counted only once, following previous literature (1).**

**a)** Field [21000-0.0] "**Ethnic background**"; in the study were retained participants with codes: 1 "White", 1001 "British", 1002 "Irish", 1003 "Any other white background". Answer: -1 "Do not know" OR Answer: -3 "Prefer not to answer" replaced with missing. Excluded participants with missing values.

**b) Anthropometric measurements** were obtained from Fields [48-0.0] "Waist circumference", Field [49-0.0] "Hip circumference", Field [50-0.0] "Standing height" and Field [21002-0.0] "Weight". Excluded participants with missing values.

**c)** Excluded were participants with code 1 for Field [22019-0.0] "**Sex chromosome aneuploidy**" OR with a **mismatch** between Field [22001-0.0] "Genetic sex" and Field [34-0.0] "Sex (self-reported)". Missing "Genetic sex" replaced with self-reported sex.

**d)** Field [3140-0.0] "**Pregnant**"; Answer: 1 "Yes", OR Answer 2: "Unsure", OR Missing. Excluded participants with Answer: 1 or missing.

**e**) Excluded participants with **prevalent cancer cases at enrolment**. Field [134.0.0] "**Self-reported cancers entered**", Answer: >0 "Yes" or Answer:0 "No" OR Field [40005.0.0] "**Date of cancer diagnosis**" with prevalent dates than Field [53.0.0] "**Date of attending assessment centre**".
Participants with missing values were assigned the median sex-specific category (No).

**f**) Excluded participants with **incident cancer cases or deaths within 2 years post-enrollment**. Field [40005.0.0] "date of cancer diagnosis" and Field [40000.0.0] "date of death, acquired from central registry" with Field [53.0.0] "date of assessment". They were included if the differences in the dates were < 365.25*2.

**g**) Exclusions comprised **1) pituitary, adrenal, and other endocrine non-cancer illness self-reported at enrolment, 2) thyroid-related non-cancer illness, 3) diabetes status at enrolment and anti-diabetic medication, 4) liver-related non-cancer illness or kidney failure self-reported at enrolment, 5) inflammatory bowel disease, 6) chronic respiratory non-cancer illness or heart failure** defined as described in the literature (1).
Participants with missing values were retained in the analyses.

<u>Note from UK Biobank</u>: If the participant was uncertain of the type of illness they had, then they described it to the interviewer (a trained nurse), who attempted to place it within the coding tree. If the disease could not be in the coding tree, the interviewer entered a free-text description. These free-text descriptions were subsequently examined by a doctor and, where possible, matched to entries in the coding tree. Free-text descriptions that could not be matched with very high probability have been marked as "unclassifiable".

**h**) **Lipid-lowering drugs or glucocorticoids** exclusions defined as described in the literature (1).
Participants with missing values retained in the analyses.

**Supplementary Text S2**. Metabolic biomarker measurements by Nuclear Magnetic Resonance.

Ethylenediaminetetraacetic acid (EDTA) plasma samples from aliquot 3 were measured using Nightingale Health's NMR-based metabolic biomarker profiling platform. If comparisons are made to the clinical biochemistry markers available in the UK Biobank, please note that those were measured from serum samples, primarily from aliquot1. The phase 1 and 2 samples were a random subset of the full cohort. The samples were prepared directly in 96 well-plates by UK Biobank. At least 85 µL plasma was aliquoted in each well using TECAN freedom EVO 150 robotic liquid handlers, which have coefficients of variation in pipetting volume at <0.75% across eight tips. Plasma samples were shipped to Nightingale Health's laboratories in the 96-well plates on dry ice in sample batches of ~5,000-20,000. Details of the metabolic biomarker profiling platform and experimentation have been described elsewhere (2). In brief, EDTA plasma samples were stored in a freezer at -80°C. Before preparation, frozen samples were slowly thawed at +4°C overnight and then mixed gently and centrifuged (3 min, 3400´g, +4°C) to remove the possible precipitate. Aliquots of each sample were transferred into 3-mm outer-diameter NMR tubes and mixed in 1:1 ratio with a phosphate buffer (75mM Na2HPO4 in 80%/20% H2O/D2O, pH 7.4, also including 0.08% sodium 3-(trimethylsilyl) propionate-2,2,3,3-d4 and 0.04% sodium azide) automatically with an automated liquid handler (PerkinElmer Janus Automated Workstation). Nightingale Health performed real-time monitoring of the measurement consistency within and between spectrometers throughout the UK Biobank samples. Two control samples provided by Nightingale Health were included in each 96-well plate to track consistency across multiple spectrometers. Furthermore, two blind duplicate samples provided by the UK Biobank were included in each well plate, with the position information unlocked only after the results were delivered. Coefficient of variation (CV) targets across the metabolic biomarker profile were pre-specified for both Nightingale Health's internal control samples and UK Biobank's blind duplicates. The targets were met for each consecutively measured batch of ~25,000 samples. For most of the metabolic biomarkers the CVs are below 5%. For more details on metabolic biomarker measurements by NMR, see the document download from: https://biobank.ndph.ox.ac.uk/ukb/ukb/docs/nmrm_companion_doc.pdf

**Supplementary Text S3**. Definition of covariates.

**Alcohol consumption** was derived from Field “About how often do you drink alcohol?” [1558.0.0]. Answer 5 “Special occasions only” and 4 “One to three times a month” coded as “1-3 times/month”.

**Smoking Status** was derived from Field “Smoking Status” [20116.0.0].

**Metabolic equivalent of task** (MET) was derived by summing up the minutes per week of the Fields “MET per week for moderate activity” [22038.0.0], Fields “MET per week for vigorous activity” [22039.0.0] and Fields “MET minutes per week for walking” [22037.0.0].

**Weight change compared with 1 year ago** was derived from Field “Compared with one year ago, has your weight changed?” [2306.0.0]. Answers 0: “No - weigh about the same” coded as “No”. Answers 2: “Yes - gained weight” and 3: “Yes - lost weight” coded as “Yes”.

**Townsend deprivation index** as described in the literature (3).

**Paracetamol use** (regularly in last four weeks), **Nonsteroidal anti-inflammatory drugs** (NSAID), and **ever contraceptive use** were defined as described in the literature (1).

**Hormone Replacement Therapy**: was derived from Field “Have you ever used hormone replacement therapy (HRT)” [2814.0.0]. Field [6153-0/3] “Medication for cholesterol, blood pressure, diabetes, or take exogenous hormones”, Answer 4 “Hormone Replacement Therapy” coded as “Yes”.

**Menopausal status** was derived from Field “Had menopause” [2724.0.0]. Values 2: “Not sure - had a hysterectomy”, 3: ” Not sure - other reason” replaced with missing.

At each covariate values, -3: “Prefer not to answer” and -1: “Do not know” replaced with missing. Missing values were excluded from the analysis (**Supplementary Figure S6**).

### Supplementary Tables

**Supplementary Table S1.** Overall and sex-specific baseline characteristics of Epirus Health Study.

| Characteristics | All<br>N = 1,127 | EHS Cohort<br>Females<br>N = 615 (54.70) | Males<br>N = 512 (45.30) |
|---|---|---|---|
| **Age at enrollment** (years) | 44.70 ± 10.65 | 44.43 ± 10.46 | 45.03 ± 10.89 |
| **Sex** | | 615 (54.60) | 512 (45.40) |
| **Education** | | | |
| Primary and secondary school [a] | 58 (5.15) | 34 (5.53) | 24 (4.70) |
| High school [b] | 309 (27.44) | 144 (23.41) | 165 (32.29) |
| Higher education [c] | 759 (67.41) | 437 (71.06) | 322 (63.01) |
| **Smoking status** | | | |
| Never | 506 (44.90) | 293 (47.64) | 213 (41.60) |
| Previous | 217 (19.25) | 99 (16.10) | 118 (23.05) |
| Current | 404 (35.85) | 223 (36.26) | 181 (35.35) |
| **Alcohol status** | | | |
| Never | 120 (10.65) | 83 (13.50) | 37 (7.23) |
| Less than once/month | 316 (28.04) | 216 (35.12) | 100 (19.53) |
| 1-3 times/month | 184 (16.33) | 104 (16.91) | 80 (15.62) |
| 1-2 times/week | 359 (31.85) | 160 (26.02) | 199 (38.87) |
| 3-4 times/week | 91 (8.07) | 34 (5.53) | 57 (11.13) |
| Almost everyday | 57 (5.06) | 18 (2.93) | 39 (7.62) |
| **METs** (minutes/week) | 630 (0, 1,530) | 540 (0, 1,260) | 720 (90, 1,800) |
| **Height** (cm) | 169 (162, 176) | 163 (159, 167) | 176.85 ± 6.38 |
| **Adiposity indices** | | | |
| Weight (kg) | 75.10 (64.55, 85.60) | 66.10 (59.10, 74.90) | 84.10 (76.70, 93.83) |
| Body fat (%) | 26.80 (21.70, 32.90) | 31.11 ± 6.73 | 22.64 ± 6.03 |
| Waist circumference (cm) | 90 (80.50, 99) | 84 (76, 92) | 96 (88.88, 105) |
| Hip circumference (cm) | 99 (95, 105) | 99 (94, 105) | 100 (95, 104) |
| Waist to hip ratio | 0.89 (0.83, 0.97) | 0.84 (0.79, 0.89) | 0.96 (0.91, 1.02) |
| BMI (kg/m$^2$) | 25.79 (23.33, 28.79) | 27.78 (22.20, 27.94) | 26.84 (25.08, 29.37) |
| ABSI | 78.60 (73.57, 83.34) | 76.44 ± 6.57 | 81.47 (75.71, 84.68) |
| HI | 57.73 (47.84, 64.34) | 63.80 (61.02, 66.25) | 47.66 ± 1.98 |
| Waist to HI ratio | 3.96 (3.70, 4.23) | 3.75 (3.55, 3.98) | 4.19 (3.99, 4.46) |

Mean ± standard deviation or median [25o, 75o] and frequency (percentage) are presented for continuous and categorical variables, respectively.

Abbreviations: a Elementary school or junior high school, up to 9 years of education. b High school, up to 12 years of education. c University degree/MSc/PhD/Postdoc, more than 13 years of education; ABSI, a body shape index; BMI, body mass index; HI, hip index; METs, metabolic equivalents of task.

**Supplementary Table S2.** Overlapped metabolites of the eight metabolic signatures. Coefficients expressed per 1 SD increment of respective metabolite, sorted from most common to most distinct across metabolic signatures. Abbreviations: ABSI, a body shape index; BMI, body mass index; -C, cholesterol; HI, hip index; -TG, triglycerides; -PL, Phospholipids; -CE, Cholesteryl esters; -FC, Free cholesterol; -L, Total lipids; -P, Lipoprotein particle concentrations; XXL-, Chylomicrons and extremely large; XL-, Very large; L-, Large; M-, Medium; S-, Small; XS- Very small; LA, Linoleic Acid; DHA, docosahexaenoic acid; FA, fatty acids; HDL, high-density lipoprotein; IDL, intermediate-density lipoproteins; LDL, low-density lipoprotein; MUFA, monounsaturated fatty acid; PUFA, polyunsaturated fatty acid; SFA, saturated fatty acid; VLDL, very low-density lipoprotein; VHDL, very high-density lipoprotein.

**(Females)**

| Metabolites | BF% | WC | HC | WHR | BMI | ABSI | HI | WHI |
|---|---|---|---|---|---|---|---|---|
| Albumin | -0.104 | -0.091 | -0.105 | -0.046 | -0.093 | -0.048 | -0.064 | |
| Average Diameter for HDL Particles | -0.454 | -0.211 | -0.013 | -0.308 | -0.633 | -0.059 | 0.113 | |
| Glycoprotein Acetyls | 0.229 | 0.202 | 0.166 | 0.154 | 0.177 | 0.087 | 0.056 | |
| Tyrosine | 0.058 | 0.07 | 0.034 | | 0.055 | 0.058 | 0.046 | |
| Creatinine | | | 0.011 | | 0.036 | -0.057 | -0.055 | -0.033 |
| Glutamine | -0.068 | -0.068 | -0.049 | | -0.063 | -0.016 | | |
| Glycine | | | | -0.05 | -0.012 | -0.037 | 0.039 | -0.038 |
| Linoleic Acid to Total Fatty Acids % | -0.292 | -0.253 | -0.176 | -0.078 | -0.338 | | | |
| Monounsaturated Fatty Acids to Total Fatty Acids % | 0.281 | 0.298 | 0.698 | | 0.832 | | | 0.253 |
| 3-Hydroxybutyrate | -0.105 | -0.031 | -0.075 | | -0.089 | | | |
| Apolipoprotein A1 | 0.421 | 1.829 | 0.507 | | 1.099 | | | |
| Cholesterol to Total Lipids in Medium VLDL % | -0.448 | -0.548 | -0.262 | | -0.478 | | | |
| Cholesteryl Esters in Large VLDL | | -0.852 | -0.632 | -0.647 | -0.837 | | | |
| Concentration of Small HDL Particles | 1.192 | | 1.179 | | 1.116 | 0.035 | | |
| Free Cholesterol in Very Small VLDL | 0.834 | | 0.538 | 0.441 | 0.512 | | | |
| Leucine | -0.027 | | -0.111 | | -0.108 | | -0.042 | |
| Omega-6 Fatty Acids | 0.724 | 0.775 | 0.474 | | 1.191 | | | |
| Phospholipids in Large HDL | 0.552 | 0.548 | 0.045 | | 0.591 | | | |
| Total Cholines | -1.347 | -0.894 | -0.87 | | -1.073 | | | |
| Valine | 0.076 | 0.065 | 0.201 | | 0.183 | | | |
| Free Cholesterol in Very Large HDL | -0.266 | | -0.177 | | -0.108 | | | |
| Free Cholesterol to Total Lipids in Medium VLDL % | 0.174 | 0.347 | | | 0.359 | | | |
| Phospholipids to Total Lipids in Medium LDL % | -0.118 | | -0.105 | | -0.012 | | | |
| Polyunsaturated Fatty Acids to Monounsaturated Fatty Acids ratio | | 0.241 | 0.789 | | 0.969 | | | |
| Polyunsaturated Fatty Acids to Total Fatty Acids % | | | -0.391 | | -0.256 | | | 0.297 |
| Triglycerides in Large HDL | | -0.471 | -0.313 | | -0.418 | | | |
| Acetone | | | | | | 0.037 | 0.025 | |
| Concentration of IDL Particles | | | 0.201 | | 0.175 | | | |
| Cholesterol in Small HDL | | | -1.283 | | -1.372 | | | |
| Free Cholesterol to Total Lipids in Large VLDL % | | | | | | 0.075 | | 0.044 |
| Free Cholesterol to Total Lipids in Medium LDL % | 0.113 | | 0.137 | | | | | |
| Polyunsaturated Fatty Acids | | -0.438 | | | -0.741 | | | |
| Saturated Fatty Acids to Total Fatty Acids % | | | | | | | -0.055 | 0.2 |
| Total Lipids in IDL | 0.590 | 0.343 | | | | | | |
| HDL Cholesterol | | -0.885 | | | -0.604 | | | |
| Phospholipids in Chylomicrons and Extremely Large VLDL | | | | | | 0.056 | | 0.171 |
| Acetoacetate | | | | | | | 0.024 | |
| Average Diameter for VLDL Particles | | | | 0.448 | | | | |
| Cholesteryl Esters in Medium VLDL | 0.261 | | | | | | | |
| Cholesteryl Esters in Small HDL | -1.065 | | | | | | | |
| Cholesteryl Esters in Very Large VLDL | | | | | 0.348 | | | |
| Cholesteryl Esters in Very Small VLDL | -0.431 | | | | | | | |
| Cholesteryl Esters to Total Lipids in Large VLDL % | | | | | | -0.073 | | |
| Cholesteryl Esters to Total Lipids in Small HDL % | | -0.256 | | | | | | |
| Cholesteryl Esters to Total Lipids in Very Large VLDL % | | | | | | -0.062 | | |

| | | | | | | | | |
|---|---|---|---|---|---|---|---|---|
| **Citrate** | | | | | | | 0.023 | |
| **Concentration of Medium VLDL Particles** | -0.512 | | | | | | | |
| **Concentration of Very Large HDL Particles** | | -0.181 | | | | | | |
| **Docosahexaenoic Acid** | | | -0.063 | | | | | |
| **Free Cholesterol to Total Lipids in IDL %** | | | | | | | 0.039 | |
| **Free Cholesterol to Total Lipids in Medium HDL %** | | | | | 0.301 | | | |
| **Histidine** | | | | | | | -0.013 | |
| **Isoleucine** | | | | | | | | 0.038 |
| **Phospholipids in Large LDL** | -0.241 | | | | | | | |
| **Phospholipids in Small LDL** | | | | | | 0.077 | | |
| **Phospholipids to Total Lipids in Small VLDL %** | | | 0.143 | | | | | |
| **Total Lipids in Medium HDL** | | -0.798 | | | | | | |
| **Total Triglycerides** | 0.234 | | | | | | | |
| **Triglycerides in HDL** | -0.502 | | | | | | | |
| **Triglycerides in Small VLDL** | -0.537 | | | | | | | |
| **VLDL Cholesterol** | | 0.751 | | | | | | |

### (Male)

| **Metabolites** | **BF%** | **WC** | **HC** | **WHR** | **BMI** | **ABSI** | **HI** | **WHI** |
|---|---|---|---|---|---|---|---|---|
| **Glycine** | -0.152 | -0.106 | -0.082 | -0.107 | -0.117 | -0.027 | 0.033 | -0.064 |
| **Leucine** | -0.139 | -0.179 | -0.124 | -0.047 | -0.188 | -0.164 | -0.067 | -0.086 |
| **Tyrosine** | 0.180 | 0.175 | 0.145 | 0.141 | 0.170 | 0.075 | 0.018 | 0.063 |
| **Average Diameter for HDL Particles** | -0.817 | -0.824 | -0.552 | -0.779 | -1.137 | | 0.053 | |
| **Total Cholines** | -1.169 | -0.941 | -0.718 | -0.442 | -0.691 | -0.055 | | |
| **Creatinine** | | | 0.056 | | 0.044 | -0.076 | -0.047 | -0.038 |
| **Free Cholesterol in Very Small VLDL** | 1.055 | 0.873 | 0.465 | 0.749 | 0.735 | | | |
| **Monounsaturated Fatty Acids to Total Fatty Acids %** | 0.217 | 0.245 | | 0.231 | | 0.07 | | 0.056 |
| **Omega-6 Fatty Acids** | 1.157 | 1.037 | 0.536 | 0.91 | 0.97 | | | |
| **Phospholipids in Large HDL** | 0.856 | 0.807 | 0.539 | 0.739 | 1.151 | | | |
| **Albumin** | | | | -0.085 | | -0.103 | -0.023 | -0.101 |
| **Cholesteryl Esters in Large VLDL** | -1.303 | -1.285 | -1 | | -1.287 | | | |
| **Cholesteryl Esters in Very Large VLDL** | 0.792 | 0.984 | 0.505 | | 1.063 | | | |
| **Cholesteryl Esters in Very Small VLDL** | -0.473 | -0.304 | | -0.47 | -0.233 | | | |
| **Linoleic Acid** | -0.66 | -0.636 | | -0.657 | -0.699 | | | |
| **Linoleic Acid to Total Fatty Acids %** | | | -0.174 | | | -0.05 | 0.036 | -0.119 |
| **Glutamine** | | -0.045 | -0.066 | | -0.061 | | | |
| **Glycoprotein Acetyls** | | | | 0.044 | | 0.124 | | 0.122 |
| **Phospholipids in Small HDL** | 0.266 | 0.201 | 0.353 | | | | | |
| **Total Concentration of Branched-Chain Amino Acids (Leucine + Isoleucine + Valine)** | 0.066 | 0.172 | | | 0.207 | | | |
| **Valine** | | | 0.160 | | | | 0.055 | -0.078 |
| **3-Hydroxybutyrate** | | | | | | 0.039 | | 0.051 |
| **Acetone** | | | | | | 0.028 | 0.028 | |
| **Cholesterol in Very Large VLDL** | | | 0.306 | 0.122 | | | | |
| **Cholesterol to Total Lipids in Medium VLDL %** | | | -0.262 | | -0.177 | | | |
| **Cholesteryl Esters to Total Lipids in Large VLDL %** | | | | -0.184 | | | | -0.028 |
| **Concentration of Small HDL Particles** | | | 0.335 | | | | | 0.056 |
| **Docosahexaenoic Acid to Total Fatty Acids %** | | | | | -0.106 | | 0.021 | |
| **HDL Cholesterol** | -0.117 | -0.160 | | | | | | |
| **Isoleucine** | | | | | | 0.094 | | 0.096 |
| **Phenylalanine** | | | | | | 0.031 | | 0.03 |
| **Triglycerides in Large LDL** | | | | | | 0.101 | | 0.16 |
| **Triglycerides in Small LDL** | -0.595 | -0.194 | | | | | | |
| **Lactate** | | | | | | 0.041 | | 0.028 |
| **Free Cholesterol to Total Lipids in Medium HDL %** | | 0.254 | 0.148 | | | | | |
| **Free Cholesterol to Total Lipids in Small HDL %** | | | | | | -0.076 | | -0.085 |
| **Glucose** | | | | | | 0.027 | | 0.022 |
| **Acetoacetate** | | | | | | 0.026 | | |
| **Cholesterol in IDL** | | | | | | 0.076 | | |
| **Cholesterol to Total Lipids in Very Large VLDL %** | | -0.172 | | | | | | |
| **Cholesteryl Esters in Chylomicrons and Extremely Large VLDL** | | | | | | -0.093 | | |
| **Cholesteryl Esters in Medium VLDL** | | | | | | | 0.03 | |
| **Cholesteryl Esters in Small HDL** | | | -0.583 | | | | | |

| | | | | | | | | |
|---|---|---|---|---|---|---|---|---|
| **Cholesteryl Esters to Total Lipids in Medium LDL %** | | | -0.135 | | | | | |
| **Cholesteryl Esters to Total Lipids in Very Large HDL %** | | | | | | | -0.042 | |
| **Citrate** | | | | | | | 0.029 | |
| **Concentration of Medium VLDL Particles** | | | | -0.520 | | | | |
| **Degree of Unsaturation** | | | | | | | | 0.130 |
| **Free Cholesterol in Very Large HDL** | | | -0.095 | | | | | |
| **Free Cholesterol to Total Lipids in IDL %** | | | | | | | 0.020 | |
| **Free Cholesterol to Total Lipids in Large LDL %** | | | | 0.010 | | | | |
| **Free Cholesterol to Total Lipids in Very Small VLDL %** | | | | | | 0.036 | | |
| **Omega-6 Fatty Acids to Omega-3 Fatty Acids ratio** | | | | | | | | 0.117 |
| **Phospholipids in IDL** | | | | 0.349 | | | | |
| **Phospholipids to Total Lipids in Chylomicrons and Extremely Large VLDL %** | | | | | | 0.011 | | |
| **Phospholipids to Total Lipids in Medium LDL %** | | | -0.141 | | | | | |
| **Phospholipids to Total Lipids in Small LDL %** | | | | | | | | 0.091 |
| **Phospholipids to Total Lipids in Small VLDL %** | | | 0.270 | | | | | |
| **Polyunsaturated Fatty Acids to Total Fatty Acids %** | | | -0.210 | | | | | |
| **Saturated Fatty Acids** | | | -0.475 | | | | | |
| **Sphingomyelins** | 0.320 | | | | | | | |
| **Total Triglycerides** | 0.714 | | | | | | | |
| **Triglycerides in IDL** | | | | | | | -0.037 | |
| **Triglycerides in Large HDL** | | | -0.135 | | | | | |
| **Triglycerides in Very Large HDL** | | | | -0.163 | | | | |
| **Triglycerides to Total Lipids in IDL %** | -0.111 | | | | | | | |
| **Triglycerides to Total Lipids in Medium HDL %** | -0.125 | | | | | | | |
| **Triglycerides to Total Lipids in Medium LDL %** | 0.202 | | | | | | | |

**Supplementary Table S3.** Overlapped metabolites of the eight metabolic signatures of body adiposity indices in pairs and by sex **in the form of Y/X/C** (X; Distinct in vertical, Y; Distinct in horizontal; C, Common). Abbreviations: ABSI, a body shape index; BF%, body fat %; BMI, body mass index; HC; hip circumference; HI, hip index; WC, waist circumference; WHI, waist to HI index; WHR, waist to hip ratio.

**(Females)**

| | **BF%** | **WC** | **HC** | **WHR** | **BMI** | **ABSI** | **HI** | **WHI** |
|---|---|---|---|---|---|---|---|---|
| **BF%** | 0/0/30 | 9/14/16 | 9/10/20 | 3/25/5 | 12/10/20 | 8/24/6 | 8/25/5 | 7/29/1 |
| **WC** | 14/9/16 | 0/0/25 | 12/8/17 | 3/20/5 | 12/5/20 | 9/20/5 | 9/21/4 | 7/24/1 |
| **HC** | 10/9/20 | 8/12/17 | 0/0/29 | 2/23/6 | 6/3/26 | 7/22/7 | 7/23/6 | 5/26/3 |
| **WHR** | 25/3/5 | 20/3/5 | 23/2/6 | 0/0/8 | 25/1/7 | 10/4/4 | 9/4/4 | 7/7/1 |
| **BMI** | 10/12/20 | 5/12/20 | 3/6/26 | 1/25/7 | 0/0/32 | 6/24/8 | 6/25/7 | 4/28/4 |
| **ABSI** | 24/8/6 | 20/9/5 | 22/7/7 | 4/10/4 | 24/6/8 | 0/0/14 | 6/7/7 | 4/10/4 |
| **HI** | 25/8/5 | 21/9/4 | 23/7/6 | 4/9/4 | 25/6/7 | 7/6/7 | 0/0/13 | 5/10/3 |
| **WHI** | 29/7/1 | 24/7/1 | 26/5/3 | 7/7/1 | 28/4/4 | 10/4/4 | 10/5/3 | 0/0/8 |

**(Males)**

| | **BF** | **WC** | **HC** | **WHR** | **BMI** | **ABSI** | **HI** | **WHI** |
|---|---|---|---|---|---|---|---|---|
| **BF** | 0/0/22 | 3/5/17 | 16/11/11 | 8/11/11 | 4/9/13 | 17/17/5 | 11/18/4 | 17/18/4 |
| **WC** | 5/3/17 | 0/0/20 | 14/7/13 | 8/9/11 | 3/6/14 | 17/15/5 | 11/16/4 | 17/16/4 |
| **HC** | 11/16/11 | 7/14/13 | 0/0/27 | 10/18/9 | 4/14/13 | 16/21/6 | 8/20/7 | 14/20/7 |
| **WHR** | 11/8/11 | 9/8/11 | 18/10/9 | 0/0/19 | 7/9/10 | 15/12/7 | 10/14/5 | 14/12/7 |
| **BMI** | 9/4/13 | 6/3/14 | 14/4/13 | 9/7/10 | 0/0/17 | 17/12/5 | 9/11/6 | 17/13/4 |
| **ABSI** | 17/17/5 | 15/17/5 | 21/16/6 | 12/15/7 | 12/17/5 | 0/0/22 | 8/15/7 | 6/7/15 |
| **HI** | 18/11/4 | 16/11/4 | 20/8/7 | 14/10/5 | 11/9/6 | 15/8/7 | 0/0/15 | 14/8/7 |
| **WHI** | 18/17/4 | 16/17/4 | 20/14/7 | 12/14/7 | 13/17/4 | 7/6/15 | 8/14/7 | 0/0/21 |

**Supplementary Table S4.** Overall and sex-specific descriptives of the clinical biomarkers in the UK Biobank.

| Biomarkers | All<br>N = 151,601 | Female<br>80,978 (53.4%) | Male<br>70,623 (46.6%) |
|---|---|---|---|
| **WBC** | 6.58 (5.6, 7.7) | 6.6 (5.61, 7.75) | 6.53 (5.58, 7.68) |
| **Lymphocyte (10^9 cells/L)** | 1.87 (1.51, 2.26) | 1.9 (1.6, 2.31) | 1.8 (1.48, 2.2) |
| **Monocyte (10^9 cells/L)** | 0.44 (0.36, 0.55) | 0.4 (0.33, 0.5) | 0.5 (0.4, 0.6) |
| **Neutrophil (10^9 cells/L)** | 3.99 (3.23, 4.88) | 4 (3.24, 4.89) | 3.97 (3.22, 4.86) |
| **Eosinophil (10^9 cells/L)** | 0.13 (0.1, 0.2) | 0.12 (0.1, 0.2) | 0.15 (0.1, 0.22) |
| **Basophil (10^9 cells/L)** | 0.02 (0, 0.04) | 0.02 (0, 0.04) | 0.02 (0, 0.04) |
| **ALP (U/L)** | 79.3 (66.6, 94.2) | 80.4 (66.5, 96.6) | 78.2 (66.7, 91.7) |
| **ALT (U/L)** | 19.76 (15.16, 26.74) | 17.02 (13.58, 22.17) | 23.51 (18.3, 31.33) |
| **Apo A (g/L)** | 1.51 (1.35, 1.69) | 1.61 (1.45, 1.78) | 1.41 (1.28, 1.56) |
| **Apo B (g/L)** | 1.06 (0.91, 1.21) | 1.05 (0.9, 1.21) | 1.07 (0.93, 1.22) |
| **AST (U/L)** | 24.1 (20.8, 28.3) | 22.6 (19.7, 26.2) | 25.9 (22.5, 30.4) |
| **Direct bilirubin (umol/L)** | 1.59 (1.29, 2.04) | 1.44 (1.21, 1.82) | 1.75 (1.41, 2.23) |
| **Urea (mmol/L)** | 5.23 (4.49, 6.07) | 5.06 (4.33, 5.89) | 5.42 (4.68, 6.25) |
| **Cholesterol (mmol/L)** | 5.86 (5.19, 6.58) | 5.96 (5.27, 6.71) | 5.74 (5.11, 6.43) |
| **Creatinine (umol/L)** | 70.4 (61.6, 80.6) | 63 (57.1, 69.5) | 79.9 (72.8, 87.7) |
| **CRP (mg/L)** | 1.25 (0.63, 2.55) | 1.29 (0.62, 2.73) | 1.21 (0.64, 2.37) |
| **Cystatin C (mg/L)** | 0.87 (0.8, 0.96) | 0.84 (0.77, 0.93) | 0.9 (0.83, 0.99) |
| **GGT (U/L)** | 25.2 (18, 38.7) | 20.5 (15.6, 29.8) | 31.8 (23.1, 47.4) |
| **Glucose (mmol/L)** | 4.89 (4.58, 5.24) | 4.88 (4.58, 5.22) | 4.9 (4.57, 5.25) |
| **HbA1c (mmol/moL)** | 34.6 (32.3, 37) | 34.7 (32.4, 37) | 34.5 (32.27, 36.9) |
| **HDL Cholesterol (mmol/L)** | 1.41 (1.19, 1.68) | 1.57 (1.35, 1.82) | 1.26 (1.09, 1.47) |
| **IGF 1 (nmol/L)** | 21.66 (18.07, 25.16) | 21.17 (17.43, 24.83) | 22.19 (18.81, 25.49) |
| **LDL direct (mmol/L)** | 3.69 (3.18, 4.25) | 3.69 (3.16, 4.28) | 3.69 (3.2, 4.22) |
| **Lipoprotein A (nmol/L)** | 19.6 (9.3, 58.2) | 20.6 (9.6, 58.1) | 18.6 (8.97, 58.41) |
| **SHBG (mmol/L)** | 45.87 (33.06, 64.24) | 57.78 (41.89, 77.83) | 37.04 (28.12, 48.02) |
| **Total bilirubin (IU/mL)** | 8.09 (6.46, 10.42) | 7.27 (5.91, 9.15) | 9.14 (7.38, 11.64) |
| **Testosterone (nmol/L)** | 5.17 (1.03, 11.97) | 1.03 (0.73, 1.39) | 11.94 (9.81, 14.4) |
| **TRG (mmol/L)** | 1.48 (1.07, 2.12) | 1.33 (0.98, 1.85) | 1.7 (1.2, 2.44) |
| **Vitamin D (nmol/L)** | 48 (33.9, 62.9) | 48 (33.9, 62.9) | 47.9 (33.9, 62.8) |

Median [interquartile range] is presented for continuous variables. Missing values (**Supplementary Figure S4**)

Abbreviations: ALP, alkaline phosphatase; ALT, alanine aminotransferase; Apo, apolipoprotein; AST, aspartate aminotransferase; CRP, c-reactive protein; GGT, gamma glutamyltransferase; HbA1c, glycated hemoglobin; TRG, triglycerides; WBC, white blood cells.

**Supplementary Table S5**. P-values from q-test for heterogeneity between sexes for each metabolic signature and biomarker. Bonferroni correction was performed (0.05/number of biomarkers = 0.00172). Abbreviations: ALP, alkaline phosphatase; ALT, alanine aminotransferase; ABSI, a body shape index; Apo, apolipoprotein; AST, aspartate aminotransferase; BF%, body fat %; BMI, body mass index; CRP, c-reactive protein; GGT, gamma glutamyltransferase; HbA1c, glycated hemoglobin; HI, hip index; TRG, triglycerides; WBC, white blood cells; *, different directions on effect estimates between sexes.

| Biomarkers | BF% | WC | HC | WHR | BMI | ABSI | HI | WHI |
|---|---|---|---|---|---|---|---|---|
| **WBC** | 3,18E-05 | 1,24E-09 | 9,97E-09 | 4,0E-01 | 2,25E-05 | 1,07E-03 | 1,53E-05 | 1,01E-08 |
| **Lymphocyte** | >0,99 | >0,99 | >0,99 | >0,99 | >0,99 | >0,99 | >0,99 | >0,99 |
| **Monocyte** | >0,99 | >0,99 | >0,99 | 1,50E-04 | >0,99 | 2,18E-12 | 2,39E-04 | 5,65E-17 |
| **Neutrophil** | 1,76E-07 | 2,5E-13 | 1,05E-14 | 1,44E-02 | 6,3E-09 | 2,68E-01 | 1,48E-05 | 4,76E-09 |
| **Eosinophil** | 5,63E-02 | 5,61E-01 | 2,10E-01 | >0,99 | 4,82E-01 | >0,99 | >0,99 | >0,99 |
| **Basophil** | >0,99 | >0,99 | 3,53E-01 | >0,99 | 8,34E-01 | 2,17E-01 | >0,99 | 1,24E-01 |
| **ALP** | 4,89E-17 | 1,85E-11 | 7,08E-23 | 1,86E-02 | 1,02E-14 | 5,57E-01 | **5,39E-12*** | 6,97E-09 |
| **ALT** | 8,02E-11 | 7,48E-22 | 1,94E-31 | 1,83E-15 | 1,9E-18 | >0,99 | 5,77E-37 | 4,32E-11 |
| **ApoA** | 3,51E-02 | >0,99 | 2,32E-05 | 9,35E-01 | >0,99 | 6,40E-05 | >0,99 | 1,36E-08 |
| **ApoB** | 4,39E-01 | 5,79E-03 | >0,99 | 8,52E-18 | >0,99 | 1,54E-63 | >0,99 | 4,27E-11 |
| **AST** | 3,18E-08 | 1,81E-14 | 3,01E-16 | 2,32E-17 | 6,03E-08 | 7,64E-08 | 1,80E-10 | 3,89E-13 |
| **Direct bilirubin** | 4,63E-01 | 2,31E-02 | 8,09E-03 | >0,99 | 3,34E-02 | >0,99 | 7,33E-07 | 2,58E-02 |
| **Urea** | >0,99 | >0,99 | >0,99 | 1,61E-04 | >0,99 | **1,23E-16*** | >0,99 | 1,20E-05 |
| **Cholesterol** | >0,99 | >0,99 | >0,99 | 1,82E-05 | >0,99 | 6,78E-28 | 3,01E-01 | 8,89E-04 |
| **Creatinine** | 2,16E-01 | >0,99 | 2,11E-05 | >0,99 | >0,99 | 4,60E-16 | 3,99E-08 | >0,99 |
| **CRP** | 8,91E-32 | 7,46E-35 | 4,89E-55 | 3,69E-06 | 2,3E-55 | 3,17E-08 | 6,92E-37 | 1,68E-66 |
| **Cystatin C** | 1,03E-01 | 1,36E-03 | >0,99 | 2,09E-04 | 1,19E-02 | 1,92E-07 | 3,51E-17 | >0,99 |
| **GGT** | >0,99 | 7,33E-03 | 2,50E-04 | 1,10E-03 | 2,87E-03 | >0,99 | 3,71E-36 | 1,63E-24 |
| **Glucose** | >0,99 | >0,99 | >0,99 | >0,99 | >0,99 | 1,28E-04 | >0,99 | 9,53E-07 |
| **HbA1c** | 7,98E-04 | 6,63E-01 | 5,48E-03 | >0,99 | 1,91E-03 | 2,41E-01 | 1,97E-03 | 1,27E-19 |
| **HDL C** | 1,75E-01 | 1,92E-01 | 2,37E-05 | 2,78E-22 | >0,99 | 2,38E-64 | >0,99 | 2,63E-43 |
| **IGF_1** | >0,99 | >0,99 | >0,99 | >0,99 | 5,55E-01 | 6,79E-01 | >0,99 | 8,28E-02 |
| **LDL direct** | 4,68E-04 | 2,36E-07 | >0,99 | 6,09E-34 | 1,24E-01 | 1,14E-70 | 9,28E-05 | 4,40E-35 |
| **Lipoprotein A** | >0,99 | >0,99 | 1,76E-01 | 3,09E-02 | >0,99 | **2,02E-4*** | >0,99 | 8,30E-03 |
| **SHBG** | >0,99 | >0,99 | 6,86E-03 | 6,02E-02 | >0,99 | 1,48E-55 | 1,18E-07 | 7,55E-01 |
| **Total bilirubin** | 1,33E-09 | 2,65E-12 | 2,50E-20 | >0,99 | 8,05E-14 | 6,15E-04 | 6,71E-04 | 1,01E-02 |
| **Testosterone** | 3,16E-36 | 1,71E-34 | **3,30E-72*** | **1,76E-76*** | 1,9E-34 | **4,83E-75*** | 2,31E-70 | 3,41E-114 |
| **TG** | 1,38E-01 | 7,24E-02 | 8,25E-04 | 2,90E-35 | 6,08E-12 | 9,88E-324 | 1,93E-70 | 2,39E-172 |
| **Vitamin D** | 9,98E-04 | 5,46E-04 | 1,29E-01 | 7,20E-02 | 1,77E-05 | >0,99 | 2,96E-03 | >0,99 |

**Supplementary Table S6.** Sex-specific associations of the standardized adiposity indices, the respective standardized metabolic signatures and the associations of their mutual adjustments with 29 clinical biomarkers in z-score. Models are adjusted for age, alcohol consumption, smoking status, metabolic equivalent of task, town deprivation index, paracetamol use, NSAID, weight change, fasting time, and in females also menopausal status, oral contraceptives use and HRT. Coefficients are expressed as 1/SD increase. Abbreviations: ABSI, a body shape index; ALP, alkaline phosphatase; ALT, alanine aminotransferase; Apo, apolipoprotein; AST, aspartate aminotransferase; BF%, body fat%; BMI, body mass index; CRP, c-reactive protein; GGT, gamma glutamyltransferase; HbA1c, glycated hemoglobin; TRG, triglycerides; MS, metabolic signature; HC, hip circumference; HI, hip index; WBC, white blood cells; WC, waist circumference; WHI, waist to HI ratio; WHR, waist to hip ratio; *, Bonferroni correction significant (0.05/number of biomarkers = 0.00172).

| BF% | Female | | | | Male | | | |
|---|---|---|---|---|---|---|---|---|
| **Biomarkers** | **Adiposity** | **Adiposity adjusted for MS** | **MS** | **MS adjusted for Adiposity** | **Adiposity** | **Adiposity adjusted for MS** | **MS** | **MS adjusted for Adiposity** |
| **WBC** | 0.168 (0.16, 0.176) * | 0.008 (-0.001, 0.017) | 0.282 (0.274, 0.289) * | 0.277 (0.267, 0.286) * | 0.174 (0.167, 0.182) * | 0.051 (0.042, 0.06) * | 0.242 (0.235, 0.25) * | 0.214 (0.206, 0.223) * |
| **Lymphocyte** | 0.132 (0.124, 0.14) * | 0.04 (0.031, 0.05) * | 0.182 (0.174, 0.19) * | 0.159 (0.149, 0.168) * | 0.169 (0.161, 0.176) * | 0.085 (0.076, 0.094) * | 0.192 (0.185, 0.2) * | 0.146 (0.137, 0.155) * |
| **Monocyte** | 0.097 (0.089, 0.105) * | 0.025 (0.016, 0.035) * | 0.138 (0.13, 0.146) * | 0.124 (0.114, 0.133) * | 0.144 (0.137, 0.152) * | 0.063 (0.054, 0.072) * | 0.176 (0.169, 0.184) * | 0.142 (0.133, 0.151) * |
| **Neutrophil** | 0.138 (0.13, 0.146) * | -0.01 (-0.02, -0.001) | 0.25 (0.243, 0.258) * | 0.256 (0.247, 0.266) * | 0.12 (0.113, 0.128) * | 0.017 (0.008, 0.026) * | 0.19 (0.182, 0.197) * | 0.181 (0.172, 0.19) * |
| **Eosinophil** | 0.091 (0.082, 0.099) * | 0.044 (0.034, 0.054) * | 0.107 (0.099, 0.115) * | 0.081 (0.071, 0.091) * | 0.075 (0.067, 0.083) * | 0.053 (0.043, 0.062) * | 0.068 (0.06, 0.075) * | 0.039 (0.029, 0.048) * |
| **Basophil** | 0.023 (0.015, 0.031) * | -0.012 (-0.022, -0.002) | 0.053 (0.045, 0.061) * | 0.06 (0.05, 0.07) * | 0.028 (0.02, 0.035) * | 0.008 (-0.002, 0.017) | 0.039 (0.031, 0.046) * | 0.034 (0.025, 0.043) * |
| **ALP** | 0.22 (0.213, 0.228) * | 0.081 (0.073, 0.09) * | 0.288 (0.28, 0.295) * | 0.24 (0.231, 0.249) * | 0.121 (0.113, 0.128) * | 0.047 (0.038, 0.057) * | 0.154 (0.146, 0.162) * | 0.128 (0.119, 0.138) * |
| **ALT** | 0.23 (0.222, 0.238) * | 0.132 (0.123, 0.142) * | 0.246 (0.238, 0.254) * | 0.168 (0.159, 0.178) * | 0.355 (0.348, 0.363) * | 0.208 (0.199, 0.216) * | 0.371 (0.364, 0.378) * | 0.257 (0.249, 0.266) * |
| **ApoA** | -0.184 (-0.192, -0.176) * | 0.034 (0.025, 0.043) * | -0.356 (-0.364, -0.349) * | -0.376 (-0.385, -0.367) * | -0.185 (-0.192, -0.177) * | 0.054 (0.045, 0.063) * | -0.387 (-0.394, -0.38) * | -0.417 (-0.425, -0.408) * |
| **ApoB** | 0.216 (0.208, 0.224) * | -0.003 (-0.012, 0.006) | 0.376 (0.369, 0.384) * | 0.378 (0.369, 0.387) * | 0.218 (0.21, 0.226) * | 0.018 (0.009, 0.027) * | 0.358 (0.35, 0.365) * | 0.348 (0.339, 0.357) * |
| **AST** | 0.034 (0.026, 0.042) * | 0.008 (-0.001, 0.018) | 0.05 (0.042, 0.058) * | 0.045 (0.035, 0.055) * | 0.145 (0.137, 0.153) * | 0.072 (0.062, 0.081) * | 0.166 (0.158, 0.174) * | 0.127 (0.118, 0.136) * |
| **Direct bilirubi** | -0.095 (-0.104, -0.085) * | 0.021 (0.01, 0.032) * | -0.187 (-0.196, -0.178) * | -0.199 (-0.211, -0.188) * | -0.072 (-0.08, -0.064) * | 0.023 (0.013, 0.033) * | -0.151 (-0.159, -0.143) * | -0.164 (-0.173, -0.154) * |
| **Urea** | 0.051 (0.044, 0.059) * | -0.025 (-0.034, -0.016) * | 0.117 (0.109, 0.125) * | 0.132 (0.122, 0.141) * | -0.015 (-0.023, -0.008) * | -0.085 (-0.094, -0.075) * | 0.075 (0.067, 0.082) * | 0.121 (0.112, 0.13) * |
| **Cholesterol** | 0.125 (0.117, 0.132) * | 0.025 (0.016, 0.034) * | 0.187 (0.179, 0.194) * | 0.172 (0.163, 0.181) * | 0.132 (0.125, 0.14) * | 0.043 (0.034, 0.053) * | 0.179 (0.172, 0.187) * | 0.156 (0.147, 0.165) * |
| **Creatinine** | 0.033 (0.025, 0.041) * | 0.006 (-0.004, 0.016) | 0.051 (0.043, 0.059) * | 0.048 (0.037, 0.058) * | 0.008 (0, 0.016) * | 0.002 (-0.008, 0.012) | 0.012 (0.004, 0.019) * | 0.011 (0.001, 0.02) |
| **CRP** | 0.481 (0.474, 0.488) * | 0.226 (0.218, 0.234) * | 0.573 (0.566, 0.579) * | 0.44 (0.432, 0.448) * | 0.39 (0.383, 0.398) * | 0.217 (0.208, 0.225) * | 0.422 (0.415, 0.429) * | 0.303 (0.295, 0.312) * |
| **Cystatin** | 0.257 (0.25, 0.264) * | 0.175 (0.167, 0.184) * | 0.244 (0.237, 0.252) * | 0.142 (0.133, 0.15) * | 0.196 (0.189, 0.203) * | 0.135 (0.126, 0.144) * | 0.18 (0.173, 0.187) * | 0.106 (0.098, 0.115) * |
| **GGT** | 0.238 (0.23, 0.245) * | 0.092 (0.082, 0.101) * | 0.306 (0.298, 0.314) * | 0.252 (0.243, 0.262) * | 0.33 (0.323, 0.337) * | 0.177 (0.168, 0.185) * | 0.364 (0.357, 0.372) * | 0.268 (0.259, 0.276) * |
| **Glucose** | 0.088 (0.08, 0.097) * | 0.028 (0.018, 0.039) * | 0.12 (0.112, 0.129) * | 0.103 (0.093, 0.114) * | 0.104 (0.096, 0.113) * | 0.055 (0.045, 0.065) * | 0.116 (0.108, 0.124) * | 0.086 (0.077, 0.096) * |
| **HbA1c** | 0.111 (0.104, 0.119) * | -0.033 (-0.043, -0.024) * | 0.23 (0.222, 0.237) * | 0.25 (0.24, 0.259) * | 0.127 (0.119, 0.135) * | 0.015 (0.005, 0.024) | 0.204 (0.196, 0.211) * | 0.196 (0.187, 0.205) * |
| **HDL_C** | -0.268 (-0.276, -0.261) * | 0.022 (0.013, 0.03) * | -0.488 (-0.495, -0.481) * | -0.501 (-0.51, -0.492) * | -0.266 (-0.273, -0.258) * | 0.039 (0.031, 0.047) * | -0.512 (-0.518, -0.505) * | -0.533 (-0.541, -0.525) * |
| **IGF_1** | -0.112 (-0.12, -0.105) * | -0.094 (-0.103, -0.085) * | -0.087 (-0.095, -0.08) * | -0.032 (-0.041, -0.023) * | -0.125 (-0.133, -0.118) * | -0.12 (-0.129, -0.111) * | -0.075 (-0.082, -0.067) * | -0.009 (-0.018, 0) |
| **LDL direct** | 0.19 (0.182, 0.197) * | 0.017 (0.008, 0.026) * | 0.308 (0.301, 0.315) * | 0.298 (0.289, 0.307) * | 0.17 (0.162, 0.178) * | 0.031 (0.022, 0.04) * | 0.259 (0.252, 0.267) * | 0.243 (0.233, 0.252) * |
| **Lipoprotein A** | 0.035 (0.026, 0.044) * | 0.038 (0.027, 0.049) * | 0.017 (0.008, 0.026) * | -0.005 (-0.017, 0.006) | -0.009 (-0.018, 0) * | 0.007 (-0.003, 0.018) | -0.025 (-0.033, -0.016) * | -0.029 (-0.039, -0.018) * |
| **SHBG** | -0.387 (-0.394, -0.379) * | -0.191 (-0.2, -0.182) * | -0.45 (-0.457, -0.442) * | -0.338 (-0.347, -0.328) * | -0.265 (-0.272, -0.257) * | -0.081 (-0.089, -0.072) * | -0.366 (-0.373, -0.359) * | -0.322 (-0.33, -0.313) * |
| **Total bilirubin** | -0.111 (-0.119, -0.103) * | 0.006 (-0.004, 0.015) | -0.197 (-0.206, -0.189) * | -0.201 (-0.211, -0.191) * | -0.064 (-0.072, -0.056) * | 0 (-0.01, 0.009) | -0.112 (-0.119, -0.104) * | -0.112 (-0.121, -0.102) * |
| **Testosterone** | 0.105 (0.096, 0.113) * | 0.111 (0.1, 0.121) * | 0.055 (0.046, 0.063) * | -0.01 (-0.021, 0) | -0.248 (-0.256, -0.241) * | -0.139 (-0.149, -0.13) * | -0.266 (-0.274, -0.259) * | -0.19 (-0.199, -0.181) * |
| **TG** | 0.309 (0.302, 0.317) * | 0.013 (0.004, 0.021) | 0.52 (0.513, 0.526) * | 0.512 (0.504, 0.52) * | 0.32 (0.313, 0.328) * | 0.008 (0, 0.016) | 0.549 (0.542, 0.555) * | 0.544 (0.537, 0.552) * |
| **Vitamin D** | -0.143 (-0.151, -0.135) * | -0.128 (-0.138, -0.118) * | -0.1 (-0.109, -0.092) * | -0.025 (-0.035, -0.015) * | -0.138 (-0.146, -0.13) * | -0.091 (-0.101, -0.082) * | -0.132 (-0.14, -0.124) * | -0.082 (-0.091, -0.073) * |

| WC | Female | | | | Male | | | |
|---|---|---|---|---|---|---|---|---|
| Biomarkers | Adiposity | Adiposity adjusted for MS | MS | MS adjusted for Adiposity | Adiposity | Adiposity adjusted for MS | MS | Adiposity adjusted for MS |
| WBC | 0.18 (0.173, 0.188) * | 0.007 (-0.002, 0.016) | 0.305 (0.297, 0.312) * | 0.301 (0.291, 0.31) * | 0.158 (0.151, 0.166) * | 0.04 (0.031, 0.049) * | 0.239 (0.231, 0.246) * | 0.218 (0.209, 0.226) * |
| Lymphocyte | 0.142 (0.134, 0.149) * | 0.033 (0.023, 0.043) * | 0.207 (0.2, 0.215) * | 0.188 (0.179, 0.198) * | 0.155 (0.148, 0.163) * | 0.062 (0.053, 0.071) * | 0.204 (0.196, 0.211) * | 0.171 (0.162, 0.18) * |
| Monocyte | 0.101 (0.093, 0.109) * | 0.014 (0.005, 0.024) | 0.158 (0.15, 0.166) * | 0.15 (0.14, 0.16) * | 0.14 (0.133, 0.148) * | 0.055 (0.046, 0.064) * | 0.186 (0.179, 0.194) * | 0.158 (0.149, 0.167) * |
| Neutrophil | 0.147 (0.139, 0.155) * | -0.009 (-0.018, 0.001) | 0.265 (0.257, 0.273) * | 0.27 (0.261, 0.279) * | 0.106 (0.099, 0.114) * | 0.014 (0.005, 0.022) | 0.178 (0.17, 0.185) * | 0.171 (0.162, 0.179) * |
| Eosinophil | 0.103 (0.095, 0.112) * | 0.048 (0.038, 0.058) * | 0.124 (0.116, 0.133) * | 0.097 (0.087, 0.107) * | 0.074 (0.066, 0.081) * | 0.038 (0.029, 0.047) * | 0.085 (0.078, 0.093) * | 0.065 (0.056, 0.074) * |
| Basophil | 0.03 (0.021, 0.038) * | -0.004 (-0.014, 0.005) | 0.056 (0.048, 0.064) * | 0.059 (0.049, 0.069) * | 0.03 (0.022, 0.038) * | 0.012 (0.003, 0.022) | 0.039 (0.031, 0.047) * | 0.032 (0.023, 0.041) * |
| ALP | 0.195 (0.188, 0.203) * | 0.063 (0.054, 0.071) * | 0.266 (0.259, 0.274) * | 0.23 (0.221, 0.239) * | 0.092 (0.084, 0.1) * | 0.016 (0.007, 0.025) * | 0.148 (0.14, 0.156) * | 0.14 (0.131, 0.149) * |
| ALT | 0.272 (0.264, 0.28) * | 0.177 (0.168, 0.187) * | 0.267 (0.259, 0.275) * | 0.164 (0.155, 0.173) * | 0.337 (0.329, 0.344) * | 0.18 (0.172, 0.189) * | 0.383 (0.376, 0.39) * | 0.288 (0.28, 0.296) * |
| ApoA | -0.231 (-0.239, -0.223) * | 0.02 (0.012, 0.029) * | -0.424 (-0.431, -0.417) * | -0.436 (-0.445, -0.427) * | -0.227 (-0.234, -0.219) * | 0.014 (0.005, 0.022) * | -0.437 (-0.443, -0.43) * | -0.444 (-0.452, -0.436) * |
| ApoB | 0.204 (0.197, 0.212) * | 0 (-0.009, 0.009) | 0.354 (0.347, 0.361) * | 0.354 (0.345, 0.363) * | 0.147 (0.139, 0.154) * | -0.02 (-0.029, -0.011) * | 0.297 (0.289, 0.304) * | 0.307 (0.298, 0.316) * |
| AST | 0.072 (0.064, 0.08) * | 0.056 (0.046, 0.065) * | 0.06 (0.052, 0.068) * | 0.028 (0.018, 0.037) * | 0.148 (0.141, 0.156) * | 0.075 (0.066, 0.085) * | 0.174 (0.167, 0.182) * | 0.135 (0.126, 0.144) * |
| Direct bilirubin | -0.093 (-0.102, -0.084) * | 0.022 (0.011, 0.034) * | -0.187 (-0.196, -0.178) * | -0.2 (-0.211, -0.189) * | -0.06 (-0.069, -0.052) * | 0.028 (0.018, 0.037) * | -0.146 (-0.154, -0.138) * | -0.161 (-0.17, -0.151) * |
| Urea | 0.055 (0.047, 0.063) * | -0.022 (-0.031, -0.013) * | 0.121 (0.113, 0.128) * | 0.133 (0.124, 0.143) * | 0.023 (0.015, 0.03) * | -0.05 (-0.059, -0.041) * | 0.108 (0.1, 0.115) * | 0.134 (0.125, 0.143) * |
| Cholesterol | 0.096 (0.089, 0.104) * | 0.02 (0.011, 0.029) * | 0.143 (0.136, 0.151) * | 0.132 (0.122, 0.141) * | 0.055 (0.047, 0.063) * | -0.008 (-0.017, 0.002) | 0.111 (0.104, 0.119) * | 0.115 (0.106, 0.124) * |
| Creatinine | 0.061 (0.053, 0.069) * | 0.05 (0.04, 0.06) * | 0.048 (0.04, 0.057) * | 0.019 (0.009, 0.029) * | 0.051 (0.043, 0.059) * | 0.045 (0.036, 0.054) * | 0.034 (0.026, 0.042) * | 0.01 (0.001, 0.02) |
| CRP | 0.45 (0.443, 0.457) * | 0.197 (0.19, 0.205) * | 0.553 (0.546, 0.56) * | 0.438 (0.43, 0.446) * | 0.347 (0.34, 0.354) * | 0.187 (0.179, 0.195) * | 0.393 (0.386, 0.4) * | 0.294 (0.286, 0.302) * |
| Cystatin | 0.266 (0.259, 0.273) * | 0.185 (0.176, 0.193) * | 0.249 (0.242, 0.256) * | 0.142 (0.133, 0.15) * | 0.233 (0.225, 0.24) * | 0.182 (0.174, 0.19) * | 0.189 (0.182, 0.196) * | 0.093 (0.085, 0.102) * |
| GGT | 0.257 (0.249, 0.264) * | 0.113 (0.104, 0.122) * | 0.315 (0.307, 0.322) * | 0.249 (0.24, 0.258) * | 0.277 (0.27, 0.285) * | 0.117 (0.109, 0.126) * | 0.356 (0.349, 0.363) * | 0.294 (0.286, 0.303) * |
| Glucose | 0.113 (0.105, 0.122) * | 0.06 (0.05, 0.071) * | 0.127 (0.118, 0.135) * | 0.092 (0.081, 0.102) * | 0.105 (0.097, 0.113) * | 0.058 (0.048, 0.068) * | 0.118 (0.11, 0.126) * | 0.087 (0.078, 0.097) * |
| HbA1c | 0.167 (0.16, 0.175) * | 0.051 (0.042, 0.06) * | 0.231 (0.223, 0.238) * | 0.201 (0.192, 0.21) * | 0.155 (0.148, 0.163) * | 0.061 (0.053, 0.07) * | 0.204 (0.197, 0.212) * | 0.172 (0.163, 0.181) * |
| HDL_C | -0.335 (-0.343, -0.328) * | 0.013 (0.005, 0.021) * | -0.597 (-0.603, -0.59) * | -0.604 (-0.612, -0.596) * | -0.305 (-0.312, -0.297) * | 0.007 (-0.001, 0.014) | -0.571 (-0.577, -0.565) * | -0.575 (-0.582, -0.567) * |
| IGF_1 | -0.111 (-0.119, -0.104) * | -0.079 (-0.088, -0.069) * | -0.102 (-0.11, -0.095) * | -0.057 (-0.066, -0.047) * | -0.1 (-0.107, -0.092) * | -0.073 (-0.082, -0.064) * | -0.088 (-0.096, -0.08) * | -0.05 (-0.059, -0.041) * |
| LDL direct | 0.168 (0.16, 0.175) * | 0.014 (0.005, 0.023) | 0.274 (0.267, 0.282) * | 0.266 (0.257, 0.275) * | 0.096 (0.088, 0.104) * | -0.009 (-0.018, 0.001) | 0.187 (0.18, 0.195) * | 0.192 (0.183, 0.201) * |
| Lipoprotein A | 0.026 (0.017, 0.035) * | 0.048 (0.036, 0.059) * | -0.01 (-0.019, -0.001) * | -0.038 (-0.049, -0.027) * | -0.014 (-0.023, -0.005) * | 0.006 (-0.005, 0.016) | -0.034 (-0.043, -0.025) * | -0.037 (-0.047, -0.027) * |
| SHBG | -0.443 (-0.45, -0.435) * | -0.25 (-0.259, -0.241) * | -0.48 (-0.487, -0.472) * | -0.334 (-0.343, -0.325) * | -0.284 (-0.291, -0.276) * | -0.112 (-0.12, -0.104) * | -0.376 (-0.383, -0.369) * | -0.317 (-0.326, -0.309) * |
| Total bilirubin | -0.116 (-0.124, -0.108) * | 0.01 (0.001, 0.02) | -0.213 (-0.221, -0.205) * | -0.219 (-0.229, -0.209) * | -0.068 (-0.076, -0.06) * | -0.003 (-0.012, 0.006) | -0.121 (-0.129, -0.114) * | -0.12 (-0.129, -0.111) * |
| Testosterone | 0.084 (0.076, 0.093) * | 0.1 (0.089, 0.11) * | 0.032 (0.024, 0.041) * | -0.026 (-0.037, -0.016) * | -0.28 (-0.288, -0.273) * | -0.172 (-0.181, -0.163) * | -0.289 (-0.297, -0.282) * | -0.199 (-0.208, -0.19) * |
| TG | 0.377 (0.37, 0.384) * | 0.008 (0.001, 0.016) | 0.643 (0.637, 0.649) * | 0.639 (0.631, 0.646) * | 0.314 (0.307, 0.322) * | -0.016 (-0.023, -0.008) * | 0.599 (0.593, 0.605) * | 0.607 (0.6, 0.615) * |
| Vitamin D | -0.158 (-0.167, -0.15) * | -0.135 (-0.145, -0.125) * | -0.119 (-0.127, -0.111) * | -0.04 (-0.05, -0.03) * | -0.137 (-0.144, -0.129) * | -0.083 (-0.092, -0.074) * | -0.142 (-0.149, -0.134) * | -0.098 (-0.107, -0.089) * |

| HC | Female | | | | Male | | | |
|---|---|---|---|---|---|---|---|---|
| **Biomarkers** | **Adiposity** | **Adiposity adjusted for MS** | **MS** | **MS adjusted for Adiposity** | **Adiposity** | **Adiposity adjusted for MS** | **MS** | **MS adjusted for Adiposity** |
| **WBC** | 0.128 (0.12, 0.135) * | -0.004 (-0.013, 0.004) | 0.287 (0.28, 0.295) * | 0.289 (0.281, 0.298) * | 0.087 (0.08, 0.094) * | 0 (-0.008, 0.008) | 0.216 (0.209, 0.223) * | 0.216 (0.208, 0.224) * |
| **Lymphocyte** | 0.094 (0.086, 0.102) * | 0.015 (0.007, 0.024) * | 0.179 (0.172, 0.187) * | 0.172 (0.163, 0.181) * | 0.116 (0.108, 0.124) * | 0.046 (0.038, 0.054) * | 0.192 (0.184, 0.199) * | 0.174 (0.165, 0.182) * |
| **Monocyte** | 0.073 (0.065, 0.081) * | 0.001 (-0.008, 0.01) | 0.158 (0.15, 0.166) * | 0.157 (0.148, 0.166) * | 0.089 (0.081, 0.097) * | 0.018 (0.01, 0.027) * | 0.182 (0.175, 0.19) * | 0.175 (0.167, 0.183) * |
| **Neutrophil** | 0.107 (0.099, 0.115) * | -0.012 (-0.021, -0.004) | 0.255 (0.248, 0.263) * | 0.261 (0.253, 0.27) * | 0.043 (0.036, 0.051) * | -0.023 (-0.031, -0.015) * | 0.156 (0.148, 0.163) * | 0.165 (0.157, 0.173) * |
| **Eosinophil** | 0.079 (0.071, 0.087) * | 0.03 (0.021, 0.039) * | 0.122 (0.113, 0.13) * | 0.108 (0.099, 0.117) * | 0.051 (0.043, 0.058) * | 0.021 (0.012, 0.029) * | 0.083 (0.075, 0.09) * | 0.074 (0.066, 0.083) * |
| **Basophil** | 0.019 (0.011, 0.028) * | -0.01 (-0.019, -0.001) | 0.06 (0.052, 0.068) * | 0.065 (0.056, 0.074) * | 0.009 (0.002, 0.017) * | -0.005 (-0.013, 0.004) | 0.032 (0.024, 0.04) * | 0.034 (0.026, 0.042) * |
| **ALP** | 0.167 (0.16, 0.174) * | 0.049 (0.041, 0.057) * | 0.28 (0.273, 0.287) * | 0.257 (0.249, 0.265) * | 0.044 (0.036, 0.052) * | -0.012 (-0.021, -0.004) | 0.135 (0.127, 0.143) * | 0.14 (0.132, 0.148) * |
| **ALT** | 0.18 (0.172, 0.188) * | 0.096 (0.088, 0.105) * | 0.228 (0.22, 0.236) * | 0.183 (0.174, 0.192) * | 0.241 (0.233, 0.248) * | 0.111 (0.103, 0.119) * | 0.366 (0.359, 0.373) * | 0.322 (0.314, 0.329) * |
| **ApoA** | -0.159 (-0.167, -0.151) * | 0.027 (0.019, 0.035) * | -0.396 (-0.403, -0.389) * | -0.409 (-0.417, -0.401) * | -0.179 (-0.187, -0.172) * | 0.008 (0, 0.015) | -0.46 (-0.467, -0.454) * | -0.463 (-0.471, -0.456) * |
| **ApoB** | 0.112 (0.105, 0.12) * | -0.002 (-0.011, 0.006) | 0.249 (0.241, 0.256) * | 0.25 (0.241, 0.258) * | 0.085 (0.077, 0.093) * | -0.017 (-0.026, -0.009) * | 0.248 (0.24, 0.255) * | 0.255 (0.246, 0.263) * |
| **AST** | 0.028 (0.021, 0.036) * | 0.011 (0.002, 0.019) | 0.044 (0.036, 0.052) * | 0.039 (0.03, 0.048) * | 0.111 (0.103, 0.119) * | 0.053 (0.045, 0.062) * | 0.165 (0.157, 0.172) * | 0.143 (0.135, 0.152) * |
| **Direct bilirubi** | -0.056 (-0.065, -0.047) * | 0.03 (0.02, 0.041) * | -0.171 (-0.181, -0.162) * | -0.186 (-0.196, -0.175) * | -0.018 (-0.026, -0.01) * | 0.038 (0.029, 0.046) * | -0.121 (-0.129, -0.113) * | -0.136 (-0.145, -0.128) * |
| **Urea** | 0.056 (0.049, 0.064) * | -0.018 (-0.027, -0.01) * | 0.154 (0.147, 0.162) * | 0.163 (0.154, 0.171) * | 0.028 (0.02, 0.035) * | -0.046 (-0.055, -0.038) * | 0.166 (0.158, 0.173) * | 0.184 (0.176, 0.192) * |
| **Cholesterol** | 0.041 (0.034, 0.049) * | 0.018 (0.01, 0.026) * | 0.059 (0.052, 0.067) * | 0.051 (0.042, 0.059) * | 0.016 (0.008, 0.023) * | -0.008 (-0.017, 0) | 0.056 (0.048, 0.064) * | 0.059 (0.051, 0.068) * |
| **Creatinine** | 0.085 (0.077, 0.093) * | 0.058 (0.049, 0.067) * | 0.087 (0.079, 0.095) * | 0.06 (0.05, 0.069) * | 0.063 (0.055, 0.071) * | 0.014 (0.006, 0.023) * | 0.127 (0.119, 0.134) * | 0.121 (0.113, 0.129) * |
| **CRP** | 0.396 (0.389, 0.403) * | 0.171 (0.164, 0.179) * | 0.571 (0.564, 0.578) * | 0.491 (0.484, 0.498) * | 0.252 (0.245, 0.26) * | 0.122 (0.114, 0.13) * | 0.373 (0.366, 0.38) * | 0.324 (0.317, 0.332) * |
| **Cystatin** | 0.266 (0.259, 0.273) * | 0.185 (0.177, 0.193) * | 0.264 (0.257, 0.271) * | 0.177 (0.169, 0.185) * | 0.195 (0.188, 0.202) * | 0.128 (0.12, 0.136) * | 0.218 (0.211, 0.225) * | 0.167 (0.159, 0.175) * |
| **GGT** | 0.174 (0.166, 0.182) * | 0.061 (0.052, 0.069) * | 0.276 (0.268, 0.283) * | 0.247 (0.239, 0.256) * | 0.18 (0.172, 0.187) * | 0.059 (0.051, 0.067) * | 0.323 (0.315, 0.33) * | 0.299 (0.291, 0.307) * |
| **Glucose** | 0.075 (0.067, 0.084) * | 0.03 (0.021, 0.04) * | 0.112 (0.104, 0.121) * | 0.098 (0.089, 0.108) * | 0.064 (0.056, 0.072) * | 0.021 (0.013, 0.03) * | 0.114 (0.106, 0.122) * | 0.106 (0.097, 0.115) * |
| **HbA1c** | 0.11 (0.103, 0.118) * | 0.006 (-0.002, 0.015) | 0.23 (0.223, 0.237) * | 0.227 (0.219, 0.235) * | 0.107 (0.099, 0.114) * | 0.033 (0.024, 0.041) * | 0.197 (0.189, 0.204) * | 0.184 (0.175, 0.192) * |
| **HDL_C** | -0.22 (-0.228, -0.212) * | 0.018 (0.011, 0.026) * | -0.513 (-0.52, -0.506) * | -0.522 (-0.53, -0.514) * | -0.231 (-0.238, -0.223) * | 0 (-0.007, 0.007) | -0.573 (-0.579, -0.566) * | -0.572 (-0.579, -0.566) * |
| **IGF_1** | -0.102 (-0.11, -0.095) * | -0.067 (-0.075, -0.058) * | -0.108 (-0.116, -0.101) * | -0.077 (-0.086, -0.069) * | -0.064 (-0.071, -0.056) * | -0.04 (-0.048, -0.031) * | -0.076 (-0.083, -0.068) * | -0.06 (-0.068, -0.052) * |
| **LDL direct** | 0.092 (0.085, 0.1) * | 0.013 (0.004, 0.021) | 0.18 (0.172, 0.187) * | 0.174 (0.165, 0.182) * | 0.053 (0.045, 0.061) * | -0.007 (-0.016, 0.001) | 0.146 (0.138, 0.154) * | 0.149 (0.141, 0.157) * |
| **Lipoprotein A** | 0.019 (0.01, 0.028) * | 0.019 (0.008, 0.029) * | 0.01 (0.001, 0.019) * | 0.001 (-0.009, 0.012) | -0.013 (-0.021, -0.004) * | 0.002 (-0.007, 0.012) | -0.036 (-0.045, -0.028) * | -0.037 (-0.047, -0.028) * |
| **SHBG** | -0.326 (-0.333, -0.318) * | -0.153 (-0.162, -0.145) * | -0.449 (-0.456, -0.441) * | -0.377 (-0.386, -0.369) * | -0.204 (-0.211, -0.196) * | -0.069 (-0.076, -0.061) * | -0.363 (-0.37, -0.356) * | -0.336 (-0.343, -0.328) * |
| **Total bilirubin** | -0.084 (-0.092, -0.076) * | 0.022 (0.013, 0.031) * | -0.222 (-0.23, -0.214) * | -0.232 (-0.241, -0.223) * | -0.026 (-0.033, -0.018) * | 0.02 (0.012, 0.029) * | -0.106 (-0.114, -0.099) * | -0.114 (-0.123, -0.106) * |
| **Testosterone** | 0.092 (0.083, 0.101) * | 0.088 (0.078, 0.097) * | 0.051 (0.042, 0.059) * | 0.009 (-0.001, 0.019) | -0.223 (-0.231, -0.216) * | -0.134 (-0.142, -0.126) * | -0.275 (-0.282, -0.267) * | -0.221 (-0.229, -0.213) * |
| **TG** | 0.217 (0.21, 0.225) * | 0.006 (-0.002, 0.014) | 0.464 (0.457, 0.471) * | 0.461 (0.454, 0.469) * | 0.202 (0.194, 0.209) * | -0.002 (-0.009, 0.005) | 0.505 (0.498, 0.512) * | 0.506 (0.499, 0.513) * |
| **Vitamin_D** | -0.146 (-0.154, -0.138) * | -0.116 (-0.125, -0.107) * | -0.119 (-0.127, -0.111) * | -0.065 (-0.074, -0.056) * | -0.087 (-0.094, -0.079) * | -0.046 (-0.055, -0.038) * | -0.119 (-0.126, -0.111) * | -0.1 (-0.108, -0.092) * |

| WHR | Female | | | | Male | | | |
|---|---|---|---|---|---|---|---|---|
| **Biomarkers** | **Adiposity** | **Adiposity adjusted for MS** | **MS** | **MS adjusted for Adiposity** | **Adiposity** | **Adiposity adjusted for MS** | **MS** | **MS adjusted for Adiposity** |
| **WBC** | 0.147 (0.14, 0.155) * | 0.03 (0.022, 0.038) * | 0.27 (0.262, 0.278) * | 0.256 (0.247, 0.264) * | 0.168 (0.16, 0.175) * | 0.057 (0.049, 0.065) * | 0.253 (0.246, 0.261) * | 0.226 (0.218, 0.235) * |
| **Lymphocyte** | 0.122 (0.115, 0.13) * | 0.036 (0.028, 0.045) * | 0.206 (0.198, 0.214) * | 0.188 (0.179, 0.197) * | 0.134 (0.126, 0.141) * | 0.049 (0.04, 0.057) * | 0.197 (0.19, 0.205) * | 0.174 (0.166, 0.183) * |
| **Monocyte** | 0.081 (0.073, 0.089) * | 0.033 (0.025, 0.042) * | 0.119 (0.111, 0.127) * | 0.103 (0.094, 0.112) * | 0.136 (0.128, 0.144) * | 0.058 (0.049, 0.066) * | 0.188 (0.18, 0.195) * | 0.16 (0.152, 0.169) * |
| **Neutrophil** | 0.116 (0.109, 0.124) * | 0.015 (0.006, 0.023) * | 0.229 (0.221, 0.236) * | 0.221 (0.213, 0.23) * | 0.128 (0.12, 0.135) * | 0.04 (0.032, 0.049) * | 0.198 (0.191, 0.206) * | 0.179 (0.171, 0.188) * |
| **Eosinophil** | 0.078 (0.071, 0.086) * | 0.043 (0.034, 0.052) * | 0.099 (0.091, 0.107) * | 0.078 (0.069, 0.087) * | 0.068 (0.06, 0.076) * | 0.031 (0.022, 0.04) * | 0.09 (0.083, 0.098) * | 0.076 (0.067, 0.085) * |
| **Basophil** | 0.025 (0.017, 0.033) * | 0.008 (-0.001, 0.017) | 0.04 (0.032, 0.048) * | 0.036 (0.027, 0.046) * | 0.038 (0.03, 0.046) * | 0.021 (0.012, 0.03) * | 0.045 (0.037, 0.052) * | 0.035 (0.026, 0.043) * |
| **ALP** | 0.132 (0.124, 0.139) * | 0.043 (0.035, 0.051) * | 0.214 (0.207, 0.221) * | 0.193 (0.185, 0.201) * | 0.105 (0.097, 0.113) * | 0.031 (0.022, 0.04) * | 0.167 (0.16, 0.175) * | 0.152 (0.144, 0.161) * |
| **ALT** | 0.233 (0.226, 0.241) * | 0.139 (0.131, 0.148) * | 0.273 (0.265, 0.28) * | 0.205 (0.197, 0.214) * | 0.3 (0.293, 0.308) * | 0.153 (0.145, 0.161) * | 0.376 (0.369, 0.383) * | 0.304 (0.296, 0.312) * |
| **ApoA** | -0.195 (-0.203, -0.188) * | -0.004 (-0.012, 0.004) | -0.42 (-0.427, -0.413) * | -0.418 (-0.426, -0.41) * | -0.188 (-0.195, -0.18) * | 0.003 (-0.005, 0.011) | -0.392 (-0.399, -0.385) * | -0.393 (-0.401, -0.385) * |
| **ApoB** | 0.205 (0.198, 0.213) * | 0.012 (0.004, 0.02) | 0.428 (0.421, 0.435) * | 0.422 (0.414, 0.43) * | 0.156 (0.149, 0.164) * | 0.002 (-0.006, 0.011) | 0.319 (0.311, 0.326) * | 0.318 (0.309, 0.326) * |
| **AST** | 0.079 (0.071, 0.086) * | 0.064 (0.056, 0.073) * | 0.063 (0.055, 0.071) * | 0.032 (0.023, 0.04) * | 0.124 (0.117, 0.132) * | 0.055 (0.046, 0.064) * | 0.169 (0.161, 0.176) * | 0.142 (0.134, 0.151) * |
| **Direct bilirubi** | -0.088 (-0.097, -0.079) * | -0.004 (-0.014, 0.006) | -0.186 (-0.196, -0.177) * | -0.184 (-0.195, -0.174) * | -0.082 (-0.09, -0.073) * | 0.006 (-0.004, 0.015) | -0.176 (-0.184, -0.168) * | -0.179 (-0.188, -0.17) * |
| **Urea** | 0.027 (0.02, 0.035) * | 0.004 (-0.005, 0.012) | 0.053 (0.046, 0.061) * | 0.051 (0.043, 0.06) * | 0.008 (0.001, 0.016) * | -0.044 (-0.052, -0.035) * | 0.087 (0.079, 0.094) * | 0.107 (0.099, 0.116) * |
| **Cholesterol** | 0.11 (0.103, 0.117) * | 0.015 (0.007, 0.023) * | 0.215 (0.207, 0.222) * | 0.207 (0.199, 0.216) * | 0.078 (0.071, 0.086) * | 0.007 (-0.002, 0.016) | 0.15 (0.143, 0.158) * | 0.147 (0.138, 0.156) * |
| **Creatinine** | 0.005 (-0.003, 0.013) | 0.005 (-0.004, 0.014) | 0.002 (-0.006, 0.01) | -0.001 (-0.01, 0.009) | 0.021 (0.013, 0.029) * | 0.013 (0.004, 0.022) | 0.024 (0.016, 0.031) * | 0.018 (0.009, 0.027) * |
| **CRP** | 0.287 (0.28, 0.294) * | 0.089 (0.081, 0.097) * | 0.475 (0.468, 0.482) * | 0.432 (0.424, 0.44) * | 0.305 (0.297, 0.312) * | 0.123 (0.115, 0.131) * | 0.432 (0.425, 0.439) * | 0.373 (0.365, 0.381) * |
| **Cystatin** | 0.133 (0.126, 0.14) * | 0.044 (0.036, 0.052) * | 0.216 (0.209, 0.223) * | 0.195 (0.187, 0.203) * | 0.175 (0.168, 0.183) * | 0.105 (0.097, 0.114) * | 0.194 (0.187, 0.201) * | 0.144 (0.136, 0.152) * |
| **GGT** | 0.216 (0.208, 0.223) * | 0.093 (0.085, 0.102) * | 0.312 (0.305, 0.32) * | 0.267 (0.259, 0.276) * | 0.267 (0.26, 0.275) * | 0.114 (0.106, 0.122) * | 0.37 (0.362, 0.377) * | 0.315 (0.307, 0.323) * |
| **Glucose** | 0.096 (0.088, 0.105) * | 0.051 (0.042, 0.06) * | 0.124 (0.116, 0.133) * | 0.1 (0.09, 0.109) * | 0.104 (0.096, 0.112) * | 0.059 (0.05, 0.068) * | 0.121 (0.113, 0.129) * | 0.093 (0.084, 0.102) * |
| **HbA1c** | 0.141 (0.134, 0.149) * | 0.057 (0.049, 0.065) * | 0.211 (0.204, 0.219) * | 0.184 (0.175, 0.192) * | 0.139 (0.132, 0.147) * | 0.044 (0.035, 0.052) * | 0.216 (0.209, 0.224) * | 0.195 (0.187, 0.204) * |
| **HDL_C** | -0.295 (-0.302, -0.287) * | -0.006 (-0.012, 0.001) | -0.634 (-0.64, -0.628) * | -0.631 (-0.638, -0.624) * | -0.261 (-0.269, -0.254) * | -0.005 (-0.013, 0.002) | -0.532 (-0.538, -0.525) * | -0.529 (-0.536, -0.522) * |
| **IGF_1** | -0.061 (-0.068, -0.053) * | -0.039 (-0.048, -0.031) * | -0.066 (-0.073, -0.058) * | -0.047 (-0.055, -0.038) * | -0.091 (-0.098, -0.083) * | -0.064 (-0.073, -0.055) * | -0.085 (-0.093, -0.077) * | -0.055 (-0.063, -0.046) * |
| **LDL direct** | 0.169 (0.162, 0.177) * | 0.008 (0, 0.016) | 0.356 (0.349, 0.363) * | 0.352 (0.344, 0.36) * | 0.107 (0.099, 0.115) * | 0.008 (-0.001, 0.017) | 0.208 (0.2, 0.215) * | 0.204 (0.195, 0.212) * |
| **Lipoprotein A** | 0.021 (0.012, 0.03) * | 0.045 (0.035, 0.055) * | -0.03 (-0.039, -0.021) * | -0.051 (-0.062, -0.041) * | -0.009 (-0.018, 0) * | -0.002 (-0.012, 0.008) | -0.016 (-0.024, -0.007) * | -0.015 (-0.025, -0.005) |
| **SHBG** | -0.351 (-0.358, -0.343) * | -0.181 (-0.189, -0.172) * | -0.459 (-0.466, -0.451) * | -0.371 (-0.379, -0.363) * | -0.257 (-0.264, -0.249) * | -0.094 (-0.102, -0.086) * | -0.38 (-0.387, -0.373) * | -0.335 (-0.343, -0.327) * |
| **Total bilirubin** | -0.094 (-0.102, -0.086) * | -0.018 (-0.026, -0.009) * | -0.176 (-0.184, -0.168) * | -0.167 (-0.176, -0.158) * | -0.084 (-0.092, -0.076) * | -0.014 (-0.023, -0.005) | -0.151 (-0.159, -0.143) * | -0.144 (-0.153, -0.135) * |
| **Testosterone** | 0.035 (0.026, 0.043) * | 0.032 (0.023, 0.042) * | 0.02 (0.012, 0.029) * | 0.005 (-0.005, 0.014) | -0.225 (-0.233, -0.217) * | -0.109 (-0.117, -0.1) * | -0.291 (-0.298, -0.283) * | -0.239 (-0.248, -0.231) * |
| **TG** | 0.364 (0.357, 0.371) * | 0.021 (0.016, 0.027) * | 0.758 (0.753, 0.763) * | 0.748 (0.742, 0.753) * | 0.309 (0.301, 0.316) * | 0.001 (-0.006, 0.008) | 0.634 (0.628, 0.64) * | 0.633 (0.627, 0.64) * |
| **Vitamin D** | -0.092 (-0.1, -0.084) * | -0.057 (-0.067, -0.048) * | -0.103 (-0.111, -0.095) * | -0.076 (-0.085, -0.066) * | -0.132 (-0.14, -0.124) * | -0.076 (-0.085, -0.067) * | -0.15 (-0.158, -0.143) * | -0.114 (-0.123, -0.105) * |

| BMI | Female | | | | Male | | | |
|---|---|---|---|---|---|---|---|---|
| **Biomarkers** | **Adiposity** | **Adiposity adjusted for MS** | **MS** | **MS adjusted for Adiposity** | **Adiposity** | **Adiposity adjusted for MS** | **MS** | **MS adjusted for Adiposity** |
| **WBC** | 0.182 (0.174, 0.19) * | 0.022 (0.013, 0.032) * | 0.287 (0.28, 0.295) * | 0.274 (0.265, 0.284) * | 0.156 (0.149, 0.164) * | 0.038 (0.029, 0.046) * | 0.232 (0.225, 0.239) * | 0.211 (0.203, 0.22) * |
| **Lymphocyte** | 0.149 (0.141, 0.157) * | 0.059 (0.049, 0.068) * | 0.19 (0.182, 0.197) * | 0.156 (0.146, 0.165) * | 0.175 (0.168, 0.183) * | 0.088 (0.079, 0.097) * | 0.203 (0.196, 0.211) * | 0.156 (0.147, 0.165) * |
| **Monocyte** | 0.102 (0.094, 0.11) * | 0.018 (0.008, 0.028) * | 0.155 (0.147, 0.163) * | 0.144 (0.134, 0.154) * | 0.142 (0.134, 0.15) * | 0.057 (0.048, 0.066) * | 0.182 (0.174, 0.189) * | 0.151 (0.142, 0.16) * |
| **Neutrophil** | 0.146 (0.138, 0.154) * | -0.001 (-0.01, 0.009) | 0.252 (0.244, 0.26) * | 0.252 (0.243, 0.262) * | 0.097 (0.089, 0.104) * | 0.001 (-0.008, 0.01) | 0.171 (0.163, 0.178) * | 0.17 (0.161, 0.179) * |
| **Eosinophil** | 0.098 (0.09, 0.107) * | 0.044 (0.034, 0.054) * | 0.119 (0.111, 0.127) * | 0.094 (0.084, 0.104) * | 0.068 (0.06, 0.076) * | 0.033 (0.024, 0.043) * | 0.079 (0.072, 0.087) * | 0.061 (0.052, 0.071) * |
| **Basophil** | 0.034 (0.026, 0.042) * | 0.002 (-0.008, 0.012) | 0.056 (0.048, 0.064) * | 0.055 (0.045, 0.065) * | 0.032 (0.024, 0.04) * | 0.018 (0.009, 0.027) * | 0.035 (0.027, 0.043) * | 0.025 (0.016, 0.034) * |
| **ALP** | 0.191 (0.184, 0.198) * | 0.052 (0.043, 0.061) * | 0.268 (0.261, 0.275) * | 0.238 (0.229, 0.247) * | 0.073 (0.066, 0.081) * | -0.002 (-0.012, 0.007) | 0.134 (0.126, 0.141) * | 0.135 (0.126, 0.144) * |
| **ALT** | 0.257 (0.249, 0.264) * | 0.164 (0.154, 0.173) * | 0.254 (0.246, 0.262) * | 0.159 (0.15, 0.169) * | 0.355 (0.348, 0.362) * | 0.202 (0.193, 0.21) * | 0.383 (0.376, 0.39) * | 0.274 (0.266, 0.282) * |
| **ApoA** | -0.216 (-0.224, -0.208) * | 0.039 (0.03, 0.048) * | -0.415 (-0.423, -0.408) * | -0.438 (-0.447, -0.429) * | -0.225 (-0.232, -0.217) * | 0.018 (0.01, 0.026) * | -0.424 (-0.431, -0.418) * | -0.434 (-0.442, -0.426) * |
| **ApoB** | 0.181 (0.173, 0.189) * | 0.002 (-0.007, 0.011) | 0.308 (0.3, 0.315) * | 0.306 (0.297, 0.315) * | 0.153 (0.145, 0.161) * | -0.003 (-0.012, 0.006) | 0.278 (0.271, 0.286) * | 0.28 (0.271, 0.289) * |
| **AST** | 0.06 (0.052, 0.068) * | 0.039 (0.03, 0.049) * | 0.058 (0.051, 0.066) * | 0.036 (0.026, 0.045) * | 0.176 (0.168, 0.184) * | 0.112 (0.102, 0.121) * | 0.175 (0.168, 0.183) * | 0.115 (0.106, 0.124) * |
| **Direct bilirubi** | -0.091 (-0.1, -0.082) * | 0.015 (0.004, 0.027) | -0.172 (-0.181, -0.162) * | -0.18 (-0.192, -0.169) * | -0.062 (-0.07, -0.054) * | 0.019 (0.009, 0.029) * | -0.133 (-0.141, -0.125) * | -0.143 (-0.153, -0.134) * |
| **Urea** | 0.068 (0.061, 0.076) * | -0.032 (-0.041, -0.022) * | 0.153 (0.146, 0.161) * | 0.172 (0.162, 0.181) * | 0.043 (0.035, 0.051) * | -0.045 (-0.054, -0.036) * | 0.132 (0.124, 0.14) * | 0.156 (0.147, 0.165) * |
| **Cholesterol** | 0.08 (0.073, 0.088) * | 0.035 (0.026, 0.045) * | 0.097 (0.09, 0.105) * | 0.077 (0.068, 0.086) * | 0.06 (0.052, 0.068) * | 0.003 (-0.006, 0.012) | 0.103 (0.095, 0.111) * | 0.101 (0.092, 0.111) * |
| **Creatinine** | 0.091 (0.083, 0.099) * | 0.047 (0.037, 0.057) * | 0.102 (0.094, 0.11) * | 0.075 (0.065, 0.085) * | 0.085 (0.077, 0.092) * | 0.053 (0.044, 0.062) * | 0.085 (0.077, 0.093) * | 0.056 (0.047, 0.066) * |
| **CRP** | 0.48 (0.473, 0.487) * | 0.222 (0.214, 0.23) * | 0.571 (0.565, 0.578) * | 0.443 (0.435, 0.451) * | 0.349 (0.342, 0.356) * | 0.203 (0.194, 0.211) * | 0.371 (0.364, 0.378) * | 0.261 (0.253, 0.27) * |
| **Cystatin** | 0.284 (0.277, 0.291) * | 0.187 (0.179, 0.196) * | 0.274 (0.267, 0.281) * | 0.165 (0.157, 0.174) * | 0.223 (0.216, 0.23) * | 0.154 (0.145, 0.163) * | 0.206 (0.199, 0.213) * | 0.123 (0.115, 0.132) * |
| **GGT** | 0.246 (0.238, 0.254) * | 0.109 (0.099, 0.118) * | 0.298 (0.291, 0.306) * | 0.236 (0.226, 0.245) * | 0.289 (0.282, 0.297) * | 0.13 (0.121, 0.138) * | 0.354 (0.347, 0.361) * | 0.284 (0.276, 0.293) * |
| **Glucose** | 0.113 (0.104, 0.121) * | 0.065 (0.055, 0.076) * | 0.119 (0.111, 0.128) * | 0.081 (0.071, 0.092) * | 0.112 (0.104, 0.12) * | 0.067 (0.057, 0.076) * | 0.118 (0.11, 0.126) * | 0.082 (0.072, 0.091) * |
| **HbA1c** | 0.153 (0.146, 0.161) * | 0.028 (0.019, 0.037) * | 0.231 (0.224, 0.239) * | 0.215 (0.206, 0.224) * | 0.16 (0.153, 0.168) * | 0.071 (0.062, 0.081) * | 0.197 (0.189, 0.204) * | 0.158 (0.149, 0.167) * |
| **HDL_C** | -0.304 (-0.312, -0.296) * | 0.032 (0.024, 0.041) * | -0.559 (-0.566, -0.552) * | -0.578 (-0.586, -0.569) * | -0.305 (-0.312, -0.298) * | 0.018 (0.01, 0.026) * | -0.568 (-0.574, -0.562) * | -0.578 (-0.585, -0.57) * |
| **IGF_1** | -0.125 (-0.132, -0.117) * | -0.091 (-0.1, -0.081) * | -0.111 (-0.118, -0.103) * | -0.058 (-0.068, -0.049) * | -0.112 (-0.12, -0.105) * | -0.096 (-0.106, -0.087) * | -0.08 (-0.088, -0.073) * | -0.028 (-0.037, -0.019) * |
| **LDL direct** | 0.148 (0.14, 0.155) * | 0.023 (0.014, 0.032) * | 0.227 (0.219, 0.234) * | 0.213 (0.204, 0.223) * | 0.099 (0.091, 0.107) * | 0 (-0.009, 0.009) | 0.176 (0.169, 0.184) * | 0.176 (0.167, 0.185) * |
| **Lipoprotein A** | 0.03 (0.02, 0.039) * | 0.035 (0.024, 0.047) * | 0.01 (0.001, 0.02) * | -0.01 (-0.021, 0.001) | -0.015 (-0.024, -0.006) * | 0.006 (-0.005, 0.016) | -0.035 (-0.043, -0.026) * | -0.038 (-0.048, -0.027) * |
| **SHBG** | -0.43 (-0.437, -0.422) * | -0.238 (-0.247, -0.229) * | -0.467 (-0.474, -0.459) * | -0.329 (-0.338, -0.32) * | -0.294 (-0.302, -0.287) * | -0.117 (-0.125, -0.108) * | -0.381 (-0.388, -0.374) * | -0.318 (-0.326, -0.309) * |
| **Total bilirubin** | -0.122 (-0.13, -0.113) * | 0 (-0.01, 0.01) | -0.208 (-0.216, -0.2) * | -0.208 (-0.218, -0.198) * | -0.069 (-0.077, -0.061) * | -0.012 (-0.021, -0.003) | -0.109 (-0.116, -0.101) * | -0.102 (-0.111, -0.093) * |
| **Testosterone** | 0.11 (0.102, 0.119) * | 0.126 (0.116, 0.137) * | 0.047 (0.038, 0.056) * | -0.027 (-0.038, -0.016) * | -0.276 (-0.283, -0.268) * | -0.164 (-0.173, -0.155) * | -0.288 (-0.296, -0.281) * | -0.2 (-0.209, -0.191) * |
| **TG** | 0.322 (0.314, 0.329) * | 0.014 (0.006, 0.022) * | 0.535 (0.529, 0.542) * | 0.527 (0.519, 0.535) * | 0.317 (0.31, 0.325) * | -0.023 (-0.031, -0.016) * | 0.595 (0.589, 0.602) * | 0.608 (0.601, 0.615) * |
| **Vitamin D** | -0.159 (-0.167, -0.15) * | -0.137 (-0.147, -0.127) * | -0.116 (-0.124, -0.108) * | -0.036 (-0.046, -0.026) * | -0.112 (-0.119, -0.104) * | -0.053 (-0.062, -0.044) * | -0.132 (-0.14, -0.125) * | -0.104 (-0.113, -0.095) * |

| ABSI | Female | | | | Male | | | |
|---|---|---|---|---|---|---|---|---|
| **Biomarkers** | **Adiposity** | **Adiposity adjusted for MS** | **MS** | **MS adjusted for Adiposity** | **Adiposity** | **Adiposity adjusted for MS** | **MS** | **MS adjusted for Adiposity** |
| **WBC** | 0.09 (0.083, 0.098) * | 0.018 (0.01, 0.026) * | 0.266 (0.258, 0.274) * | 0.261 (0.253, 0.269) * | 0.092 (0.085, 0.1) * | 0.032 (0.025, 0.04) * | 0.312 (0.305, 0.319) * | 0.305 (0.298, 0.313) * |
| **Lymphocyte** | 0.061 (0.054, 0.069) * | 0.008 (0, 0.016) | 0.194 (0.187, 0.202) * | 0.192 (0.184, 0.2) * | 0.032 (0.024, 0.04) * | -0.003 (-0.01, 0.005) | 0.176 (0.168, 0.183) * | 0.176 (0.168, 0.184) * |
| **Monocyte** | 0.048 (0.041, 0.056) * | 0.015 (0.007, 0.023) * | 0.124 (0.116, 0.132) * | 0.119 (0.111, 0.128) * | 0.065 (0.057, 0.073) * | 0.025 (0.017, 0.033) * | 0.21 (0.202, 0.218) * | 0.205 (0.197, 0.213) * |
| **Neutrophil** | 0.077 (0.07, 0.085) * | 0.015 (0.007, 0.023) * | 0.228 (0.221, 0.236) * | 0.224 (0.216, 0.232) * | 0.087 (0.08, 0.095) * | 0.035 (0.028, 0.043) * | 0.274 (0.267, 0.282) * | 0.267 (0.26, 0.275) * |
| **Eosinophil** | 0.051 (0.043, 0.059) * | 0.027 (0.019, 0.036) * | 0.093 (0.085, 0.101) * | 0.085 (0.077, 0.093) * | 0.04 (0.032, 0.048) * | 0.023 (0.015, 0.031) * | 0.091 (0.083, 0.099) * | 0.087 (0.079, 0.095) * |
| **Basophil** | 0.01 (0.003, 0.018) * | 0 (-0.008, 0.008) | 0.037 (0.029, 0.045) * | 0.037 (0.029, 0.046) * | 0.014 (0.006, 0.021) * | -0.001 (-0.009, 0.007) | 0.076 (0.068, 0.084) * | 0.076 (0.068, 0.084) * |
| **ALP** | 0.087 (0.08, 0.094) * | 0.031 (0.024, 0.038) * | 0.211 (0.203, 0.218) * | 0.201 (0.194, 0.209) * | 0.079 (0.071, 0.087) * | 0.035 (0.027, 0.042) * | 0.234 (0.226, 0.241) * | 0.227 (0.219, 0.235) * |
| **ALT** | 0.129 (0.121, 0.136) * | 0.059 (0.052, 0.067) * | 0.267 (0.259, 0.274) * | 0.249 (0.241, 0.257) * | 0.093 (0.085, 0.101) * | 0.045 (0.037, 0.052) * | 0.254 (0.247, 0.262) * | 0.245 (0.238, 0.253) * |
| **ApoA** | -0.102 (-0.109, -0.094) * | -0.017 (-0.025, -0.01) * | -0.308 (-0.315, -0.3) * | -0.302 (-0.31, -0.295) * | -0.06 (-0.067, -0.052) * | -0.01 (-0.018, -0.003) | -0.252 (-0.26, -0.245) * | -0.25 (-0.258, -0.242) * |
| **ApoB** | 0.124 (0.117, 0.132) * | -0.007 (-0.014, 0) | 0.47 (0.463, 0.477) * | 0.472 (0.465, 0.479) * | 0.058 (0.05, 0.066) * | 0.001 (-0.007, 0.009) | 0.292 (0.284, 0.299) * | 0.291 (0.283, 0.299) * |
| **AST** | 0.048 (0.04, 0.056) * | 0.032 (0.024, 0.04) * | 0.068 (0.06, 0.076) * | 0.058 (0.05, 0.067) * | 0.01 (0.002, 0.018) * | -0.015 (-0.023, -0.007) * | 0.124 (0.116, 0.132) * | 0.127 (0.119, 0.135) * |
| **Direct bilirubi** | -0.054 (-0.063, -0.045) * | -0.001 (-0.01, 0.008) | -0.197 (-0.206, -0.187) * | -0.196 (-0.206, -0.187) * | -0.046 (-0.055, -0.038) * | -0.003 (-0.011, 0.005) | -0.222 (-0.231, -0.214) * | -0.222 (-0.23, -0.214) * |
| **Urea** | 0.001 (-0.007, 0.008) | -0.013 (-0.021, -0.006) * | 0.047 (0.039, 0.055) * | 0.051 (0.043, 0.059) * | -0.032 (-0.04, -0.025) * | -0.023 (-0.031, -0.015) * | -0.051 (-0.059, -0.044) * | -0.047 (-0.055, -0.039) * |
| **Cholesterol** | 0.072 (0.065, 0.08) * | -0.014 (-0.022, -0.007) * | 0.308 (0.301, 0.315) * | 0.313 (0.305, 0.32) * | 0.036 (0.028, 0.044) * | -0.001 (-0.009, 0.007) | 0.19 (0.182, 0.197) * | 0.19 (0.182, 0.198) * |
| **Creatinine** | -0.05 (-0.058, -0.043) * | -0.015 (-0.023, -0.007) * | -0.131 (-0.139, -0.123) * | -0.127 (-0.135, -0.118) * | -0.066 (-0.073, -0.058) * | -0.021 (-0.029, -0.014) * | -0.23 (-0.237, -0.222) * | -0.225 (-0.233, -0.217) * |
| **CRP** | 0.137 (0.129, 0.144) * | 0.016 (0.009, 0.023) * | 0.439 (0.432, 0.446) * | 0.434 (0.427, 0.441) * | 0.137 (0.129, 0.145) * | 0.04 (0.033, 0.046) * | 0.504 (0.497, 0.51) * | 0.496 (0.489, 0.503) * |
| **Cystatin** | 0.059 (0.052, 0.066) * | 0.021 (0.014, 0.028) * | 0.143 (0.135, 0.15) * | 0.136 (0.129, 0.144) * | 0.075 (0.068, 0.083) * | 0.061 (0.053, 0.068) * | 0.087 (0.079, 0.094) * | 0.075 (0.067, 0.082) * |
| **GGT** | 0.124 (0.116, 0.131) * | 0.041 (0.033, 0.048) * | 0.311 (0.304, 0.319) * | 0.299 (0.291, 0.307) * | 0.103 (0.095, 0.11) * | 0.043 (0.036, 0.051) * | 0.313 (0.305, 0.32) * | 0.304 (0.296, 0.312) * |
| **Glucose** | 0.044 (0.036, 0.052) * | 0.009 (0.001, 0.018) | 0.128 (0.12, 0.137) * | 0.125 (0.117, 0.134) * | 0.029 (0.02, 0.037) * | -0.007 (-0.015, 0.001) | 0.179 (0.171, 0.188) * | 0.181 (0.173, 0.189) * |
| **HbA1c** | 0.084 (0.077, 0.091) * | 0.026 (0.019, 0.034) * | 0.216 (0.208, 0.223) * | 0.208 (0.2, 0.216) * | 0.051 (0.044, 0.059) * | 0.005 (-0.003, 0.013) | 0.238 (0.23, 0.245) * | 0.237 (0.229, 0.245) * |
| **HDL_C** | -0.167 (-0.175, -0.16) * | -0.024 (-0.03, -0.017) * | -0.524 (-0.531, -0.517) * | -0.517 (-0.524, -0.51) * | -0.084 (-0.092, -0.077) * | -0.018 (-0.025, -0.01) * | -0.343 (-0.35, -0.336) * | -0.339 (-0.347, -0.332) * |
| **IGF_1** | -0.035 (-0.043, -0.028) * | -0.019 (-0.026, -0.011) * | -0.066 (-0.074, -0.059) * | -0.061 (-0.069, -0.053) * | -0.048 (-0.056, -0.041) * | -0.032 (-0.039, -0.024) * | -0.092 (-0.1, -0.084) * | -0.086 (-0.094, -0.078) * |
| **LDL direct** | 0.103 (0.096, 0.111) * | -0.011 (-0.018, -0.004) * | 0.41 (0.403, 0.417) * | 0.413 (0.406, 0.421) * | 0.045 (0.037, 0.052) * | 0.001 (-0.006, 0.009) | 0.22 (0.212, 0.228) * | 0.22 (0.212, 0.228) * |
| **Lipoprotein A** | 0.006 (-0.003, 0.014) | 0.018 (0.009, 0.028) * | -0.041 (-0.05, -0.032) * | -0.047 (-0.056, -0.037) * | -0.002 (-0.011, 0.007) | -0.003 (-0.012, 0.006) | 0.005 (-0.004, 0.014) | 0.006 (-0.003, 0.015) |
| **SHBG** | -0.188 (-0.195, -0.18) * | -0.076 (-0.083, -0.068) * | -0.425 (-0.432, -0.417) * | -0.402 (-0.41, -0.394) * | -0.077 (-0.085, -0.069) * | -0.032 (-0.04, -0.025) * | -0.233 (-0.241, -0.225) * | -0.226 (-0.234, -0.219) * |
| **Total bilirubin** | -0.057 (-0.065, -0.049) * | -0.008 (-0.016, 0) | -0.179 (-0.187, -0.171) * | -0.177 (-0.185, -0.169) * | -0.053 (-0.061, -0.045) * | -0.009 (-0.017, -0.001) | -0.227 (-0.235, -0.22) * | -0.226 (-0.234, -0.218) * |
| **Testosterone** | -0.007 (-0.015, 0.002) | -0.009 (-0.018, -0.001) | 0.006 (-0.003, 0.015) | 0.009 (0, 0.018) | -0.101 (-0.109, -0.093) * | -0.059 (-0.067, -0.051) * | -0.226 (-0.234, -0.218) * | -0.214 (-0.222, -0.206) * |
| **TG** | 0.234 (0.227, 0.241) * | 0.01 (0.006, 0.015) * | 0.808 (0.804, 0.813) * | 0.805 (0.8, 0.81) * | 0.114 (0.106, 0.122) * | 0.025 (0.018, 0.033) * | 0.457 (0.45, 0.464) * | 0.452 (0.445, 0.459) * |
| **Vitamin D** | -0.06 (-0.068, -0.052) * | -0.028 (-0.036, -0.02) * | -0.128 (-0.136, -0.12) * | -0.119 (-0.128, -0.111) * | -0.102 (-0.11, -0.094) * | -0.075 (-0.083, -0.067) * | -0.151 (-0.159, -0.143) * | -0.136 (-0.144, -0.128) * |

| HI | Female | | | | Male | | | |
|---|---|---|---|---|---|---|---|---|
| Biomarkers | Adiposity | Adiposity adjusted for MS | MS | MS adjusted for Adiposity | Adiposity | Adiposity adjusted for MS | MS | MS adjusted for Adiposity |
| WBC | -0.003 (-0.011, 0.004) | 0.01 (0.002, 0.017) | -0.078 (-0.086, -0.071) * | -0.08 (-0.088, -0.072) * | -0.016 (-0.023, -0.009) * | 0 (-0.007, 0.008) | -0.133 (-0.141, -0.126) * | -0.133 (-0.141, -0.126) * |
| Lymphocyte | -0.026 (-0.033, -0.018) * | -0.008 (-0.016, -0.001) | -0.11 (-0.118, -0.103) * | -0.109 (-0.117, -0.101) * | -0.021 (-0.028, -0.013) * | -0.006 (-0.014, 0.001) | -0.12 (-0.128, -0.113) * | -0.119 (-0.127, -0.112) * |
| Monocyte | -0.001 (-0.009, 0.006) | 0.001 (-0.006, 0.009) | -0.016 (-0.024, -0.008) * | -0.016 (-0.024, -0.008) * | -0.013 (-0.021, -0.006) * | -0.005 (-0.013, 0.002) | -0.066 (-0.074, -0.058) * | -0.065 (-0.073, -0.058) * |
| Neutrophil | 0.008 (0, 0.015) | 0.016 (0.009, 0.024) * | -0.047 (-0.055, -0.04) * | -0.05 (-0.058, -0.042) * | -0.01 (-0.017, -0.003) * | 0.003 (-0.005, 0.01) | -0.104 (-0.111, -0.096) * | -0.104 (-0.112, -0.097) * |
| Eosinophil | -0.001 (-0.009, 0.006) | 0.003 (-0.004, 0.011) | -0.029 (-0.037, -0.022) * | -0.03 (-0.038, -0.022) * | 0 (-0.007, 0.008) | 0.005 (-0.002, 0.013) | -0.039 (-0.047, -0.032) * | -0.04 (-0.048, -0.032) * |
| Basophil | -0.006 (-0.014, 0.002) | -0.008 (-0.016, -0.001) | 0.012 (0.004, 0.02) * | 0.014 (0.006, 0.021) * | -0.018 (-0.025, -0.01) * | -0.017 (-0.025, -0.009) * | -0.008 (-0.015, 0) | -0.006 (-0.013, 0.002) |
| ALP | 0.021 (0.014, 0.028) * | 0.019 (0.012, 0.026) * | 0.012 (0.005, 0.019) * | 0.009 (0.002, 0.016) | -0.005 (-0.013, 0.002) | 0.003 (-0.005, 0.011) | -0.069 (-0.076, -0.061) * | -0.069 (-0.077, -0.061) * |
| ALT | -0.067 (-0.074, -0.059) * | -0.042 (-0.049, -0.034) * | -0.16 (-0.168, -0.153) * | -0.153 (-0.161, -0.146) * | -0.06 (-0.067, -0.052) * | -0.025 (-0.032, -0.018) * | -0.294 (-0.301, -0.287) * | -0.291 (-0.298, -0.283) * |
| ApoA | 0.073 (0.065, 0.08) * | 0.021 (0.013, 0.028) * | 0.32 (0.313, 0.328) * | 0.317 (0.31, 0.324) * | 0.04 (0.033, 0.047) * | 0.002 (-0.005, 0.009) | 0.32 (0.313, 0.328) * | 0.32 (0.313, 0.327) * |
| ApoB | -0.066 (-0.074, -0.059) * | -0.03 (-0.037, -0.022) * | -0.229 (-0.237, -0.222) * | -0.225 (-0.232, -0.217) * | -0.043 (-0.051, -0.035) * | -0.017 (-0.024, -0.009) * | -0.223 (-0.23, -0.215) * | -0.221 (-0.228, -0.213) * |
| AST | -0.032 (-0.04, -0.025) * | -0.03 (-0.038, -0.023) * | -0.019 (-0.027, -0.012) * | -0.014 (-0.022, -0.006) * | -0.035 (-0.043, -0.028) * | -0.025 (-0.032, -0.017) * | -0.092 (-0.1, -0.085) * | -0.089 (-0.097, -0.081) * |
| Direct bilirubi | 0.013 (0.005, 0.022) * | 0.007 (-0.002, 0.016) | 0.04 (0.031, 0.049) * | 0.039 (0.03, 0.048) * | 0.015 (0.007, 0.023) * | 0.003 (-0.005, 0.011) | 0.106 (0.098, 0.114) * | 0.106 (0.098, 0.114) * |
| Urea | -0.003 (-0.01, 0.004) | 0.017 (0.01, 0.024) * | -0.12 (-0.127, -0.112) * | -0.123 (-0.13, -0.115) * | -0.016 (-0.023, -0.008) * | -0.004 (-0.011, 0.004) | -0.103 (-0.11, -0.095) * | -0.102 (-0.11, -0.095) * |
| Cholesterol | -0.033 (-0.04, -0.026) * | -0.022 (-0.029, -0.014) * | -0.073 (-0.08, -0.066) * | -0.069 (-0.076, -0.062) * | -0.023 (-0.031, -0.016) * | -0.012 (-0.019, -0.004) | -0.098 (-0.106, -0.09) * | -0.097 (-0.104, -0.089) * |
| Creatinine | -0.041 (-0.049, -0.034) * | -0.007 (-0.014, 0.001) | -0.213 (-0.221, -0.206) * | -0.212 (-0.22, -0.205) * | -0.045 (-0.052, -0.037) * | -0.012 (-0.019, -0.004) | -0.279 (-0.287, -0.272) * | -0.278 (-0.285, -0.27) * |
| CRP | 0.001 (-0.007, 0.008) | 0.013 (0.005, 0.02) * | -0.072 (-0.08, -0.065) * | -0.074 (-0.082, -0.067) * | -0.013 (-0.021, -0.006) * | 0.012 (0.005, 0.019) * | -0.212 (-0.219, -0.204) * | -0.213 (-0.221, -0.206) * |
| Cystatin | 0.022 (0.015, 0.029) * | 0.049 (0.042, 0.056) * | -0.155 (-0.162, -0.148) * | -0.163 (-0.17, -0.156) * | 0 (-0.007, 0.007) | 0.03 (0.023, 0.037) * | -0.247 (-0.254, -0.24) * | -0.251 (-0.258, -0.244) * |
| GGT | -0.044 (-0.052, -0.037) * | -0.02 (-0.027, -0.012) * | -0.155 (-0.162, -0.147) * | -0.151 (-0.159, -0.144) * | -0.048 (-0.055, -0.04) * | -0.014 (-0.021, -0.007) * | -0.288 (-0.296, -0.281) * | -0.287 (-0.294, -0.279) * |
| Glucose | -0.039 (-0.047, -0.031) * | -0.032 (-0.04, -0.024) * | -0.053 (-0.061, -0.045) * | -0.047 (-0.056, -0.039) * | -0.038 (-0.046, -0.03) * | -0.03 (-0.038, -0.022) * | -0.073 (-0.081, -0.065) * | -0.069 (-0.077, -0.061) * |
| HbA1c | -0.037 (-0.044, -0.03) * | -0.029 (-0.037, -0.022) * | -0.053 (-0.061, -0.046) * | -0.049 (-0.056, -0.041) * | -0.023 (-0.031, -0.016) * | -0.012 (-0.02, -0.005) | -0.092 (-0.1, -0.085) * | -0.091 (-0.099, -0.083) * |
| HDL_C | 0.099 (0.092, 0.107) * | 0.018 (0.011, 0.024) * | 0.5 (0.494, 0.507) * | 0.497 (0.491, 0.504) * | 0.06 (0.053, 0.068) * | 0.001 (-0.005, 0.008) | 0.501 (0.494, 0.507) * | 0.501 (0.494, 0.507) * |
| IGF_1 | -0.027 (-0.034, -0.02) * | -0.024 (-0.031, -0.017) * | -0.024 (-0.032, -0.017) * | -0.02 (-0.028, -0.013) * | -0.015 (-0.022, -0.008) * | -0.015 (-0.022, -0.008) * | -0.002 (-0.009, 0.006) | 0 (-0.008, 0.008) |
| LDL direct | -0.054 (-0.061, -0.046) * | -0.023 (-0.03, -0.015) * | -0.193 (-0.201, -0.186) * | -0.19 (-0.197, -0.182) * | -0.026 (-0.034, -0.018) * | -0.01 (-0.017, -0.002) | -0.141 (-0.149, -0.133) * | -0.14 (-0.148, -0.132) * |
| Lipoprotein A | -0.012 (-0.02, -0.003) * | -0.017 (-0.025, -0.008) * | 0.029 (0.02, 0.037) * | 0.031 (0.022, 0.04) * | -0.001 (-0.01, 0.007) | -0.005 (-0.014, 0.003) | 0.035 (0.026, 0.043) * | 0.035 (0.026, 0.044) * |
| SHBG | 0.082 (0.074, 0.09) * | 0.03 (0.022, 0.037) * | 0.323 (0.315, 0.33) * | 0.318 (0.31, 0.325) * | 0.065 (0.058, 0.073) * | 0.02 (0.014, 0.027) * | 0.382 (0.375, 0.389) * | 0.379 (0.372, 0.386) * |
| Total bilirubin | -0.003 (-0.011, 0.005) | -0.004 (-0.011, 0.004) | 0.003 (-0.005, 0.01) | 0.003 (-0.005, 0.011) | 0.004 (-0.004, 0.011) | -0.002 (-0.01, 0.006) | 0.05 (0.042, 0.058) * | 0.05 (0.042, 0.058) * |
| Testosterone | 0.001 (-0.007, 0.009) | -0.001 (-0.01, 0.007) | 0.015 (0.007, 0.023) * | 0.015 (0.007, 0.024) * | -0.004 (-0.011, 0.004) | -0.03 (-0.037, -0.022) * | 0.217 (0.209, 0.224) * | 0.22 (0.213, 0.228) * |
| TG | -0.104 (-0.112, -0.097) * | -0.024 (-0.031, -0.018) * | -0.494 (-0.501, -0.488) * | -0.49 (-0.497, -0.484) * | -0.082 (-0.09, -0.075) * | -0.005 (-0.011, 0.001) | -0.649 (-0.655, -0.644) * | -0.649 (-0.655, -0.643) * |
| Vitamin D | -0.029 (-0.036, -0.021) * | -0.035 (-0.043, -0.027) * | 0.035 (0.027, 0.043) * | 0.041 (0.033, 0.049) * | -0.01 (-0.018, -0.003) * | -0.02 (-0.028, -0.012) * | 0.082 (0.074, 0.089) * | 0.084 (0.076, 0.092) * |

| WHI | Female | | | | Male | | | |
|---|---|---|---|---|---|---|---|---|
| **Biomarkers** | **Adiposity** | **Adiposity adjusted for MS** | **MS** | **MS adjusted for adiposity** | **Adiposity** | **Adiposity adjusted for MS** | **MS** | **MS adjusted for Adiposity** |
| **WBC** | 0.068 (0.061, 0.076) * | 0.008 (0, 0.015) | 0.262 (0.254, 0.269) * | 0.26 (0.252, 0.268) * | 0.11 (0.102, 0.117) * | 0.027 (0.02, 0.034) * | 0.334 (0.327, 0.341) * | 0.327 (0.32, 0.334) * |
| **Lymphocyte** | 0.056 (0.048, 0.064) * | 0.006 (-0.001, 0.014) | 0.215 (0.208, 0.223) * | 0.214 (0.206, 0.221) * | 0.054 (0.047, 0.062) * | 0.004 (-0.004, 0.012) | 0.2 (0.192, 0.208) * | 0.199 (0.191, 0.207) * |
| **Monocyte** | 0.036 (0.028, 0.044) * | 0.009 (0.001, 0.017) | 0.118 (0.11, 0.126) * | 0.116 (0.108, 0.124) * | 0.08 (0.072, 0.087) * | 0.025 (0.018, 0.033) * | 0.221 (0.214, 0.229) * | 0.215 (0.207, 0.223) * |
| **Neutrophil** | 0.054 (0.046, 0.061) * | 0.004 (-0.004, 0.012) | 0.214 (0.207, 0.222) * | 0.214 (0.206, 0.221) * | 0.099 (0.091, 0.106) * | 0.027 (0.02, 0.035) * | 0.29 (0.283, 0.297) * | 0.283 (0.275, 0.291) * |
| **Eosinophil** | 0.036 (0.028, 0.043) * | 0.016 (0.007, 0.024) * | 0.09 (0.082, 0.098) * | 0.086 (0.078, 0.095) * | 0.041 (0.033, 0.049) * | 0.018 (0.01, 0.026) * | 0.096 (0.088, 0.104) * | 0.091 (0.083, 0.099) * |
| **Basophil** | 0.011 (0.003, 0.018) * | 0 (-0.008, 0.008) | 0.044 (0.036, 0.052) * | 0.044 (0.036, 0.052) * | 0.027 (0.02, 0.035) * | 0.008 (0, 0.016) | 0.079 (0.071, 0.087) * | 0.077 (0.069, 0.085) * |
| **ALP** | 0.046 (0.039, 0.053) * | 0.011 (0.004, 0.019) | 0.151 (0.144, 0.158) * | 0.148 (0.141, 0.155) * | 0.083 (0.076, 0.091) * | 0.029 (0.021, 0.037) * | 0.224 (0.216, 0.232) * | 0.217 (0.209, 0.225) * |
| **ALT** | 0.122 (0.114, 0.129) * | 0.071 (0.064, 0.079) * | 0.233 (0.226, 0.241) * | 0.216 (0.208, 0.223) * | 0.145 (0.137, 0.152) * | 0.071 (0.064, 0.079) * | 0.311 (0.304, 0.319) * | 0.293 (0.285, 0.301) * |
| **ApoA** | -0.101 (-0.109, -0.093) * | -0.039 (-0.046, -0.031) * | -0.276 (-0.284, -0.268) * | -0.266 (-0.274, -0.259) * | -0.088 (-0.096, -0.081) * | -0.039 (-0.047, -0.031) * | -0.208 (-0.215, -0.2) * | -0.198 (-0.205, -0.19) * |
| **ApoB** | 0.128 (0.121, 0.135) * | 0.054 (0.047, 0.061) * | 0.333 (0.326, 0.34) * | 0.32 (0.312, 0.327) * | 0.093 (0.086, 0.101) * | 0.033 (0.025, 0.041) * | 0.251 (0.243, 0.259) * | 0.243 (0.235, 0.251) * |
| **AST** | 0.055 (0.048, 0.063) * | 0.039 (0.031, 0.046) * | 0.081 (0.073, 0.089) * | 0.072 (0.064, 0.08) * | 0.043 (0.035, 0.05) * | 0.003 (-0.005, 0.011) | 0.16 (0.152, 0.168) * | 0.159 (0.151, 0.167) * |
| **Direct bilirubi** | -0.048 (-0.057, -0.04) * | -0.007 (-0.016, 0.002) | -0.186 (-0.195, -0.177) * | -0.184 (-0.194, -0.175) * | -0.062 (-0.07, -0.054) * | -0.006 (-0.014, 0.002) | -0.228 (-0.236, -0.219) * | -0.226 (-0.234, -0.218) * |
| **Urea** | -0.006 (-0.014, 0.001) | -0.02 (-0.028, -0.013) * | 0.055 (0.047, 0.062) * | 0.06 (0.052, 0.067) * | -0.018 (-0.026, -0.01) * | -0.019 (-0.027, -0.011) * | -0.002 (-0.009, 0.006) | 0.003 (-0.005, 0.011) |
| **Cholesterol** | 0.076 (0.069, 0.083) * | 0.025 (0.018, 0.032) * | 0.225 (0.218, 0.232) * | 0.219 (0.211, 0.226) * | 0.056 (0.049, 0.064) * | 0.013 (0.005, 0.021) * | 0.177 (0.169, 0.184) * | 0.173 (0.165, 0.181) * |
| **Creatinine** | -0.043 (-0.051, -0.036) * | -0.024 (-0.032, -0.016) * | -0.089 (-0.097, -0.081) * | -0.083 (-0.091, -0.075) * | -0.031 (-0.039, -0.024) * | -0.007 (-0.015, 0.001) | -0.098 (-0.106, -0.09) * | -0.096 (-0.104, -0.088) * |
| **CRP** | 0.067 (0.059, 0.075) * | -0.005 (-0.012, 0.002) | 0.308 (0.301, 0.316) * | 0.309 (0.302, 0.317) * | 0.157 (0.149, 0.164) * | 0.034 (0.027, 0.041) * | 0.498 (0.491, 0.505) * | 0.489 (0.482, 0.496) * |
| **Cystatin** | 0.002 (-0.005, 0.009) | -0.029 (-0.036, -0.021) * | 0.125 (0.118, 0.132) * | 0.132 (0.125, 0.14) * | 0.077 (0.07, 0.084) * | 0.042 (0.034, 0.049) * | 0.15 (0.143, 0.158) * | 0.14 (0.132, 0.147) * |
| **GGT** | 0.109 (0.102, 0.117) * | 0.052 (0.044, 0.059) * | 0.259 (0.252, 0.267) * | 0.247 (0.239, 0.254) * | 0.147 (0.139, 0.154) * | 0.056 (0.049, 0.064) * | 0.374 (0.367, 0.381) * | 0.359 (0.352, 0.367) * |
| **Glucose** | 0.048 (0.04, 0.056) * | 0.023 (0.014, 0.031) * | 0.113 (0.105, 0.121) * | 0.108 (0.099, 0.116) * | 0.058 (0.05, 0.066) * | 0.014 (0.006, 0.023) * | 0.177 (0.169, 0.185) * | 0.174 (0.165, 0.182) * |
| **HbA1c** | 0.077 (0.07, 0.085) * | 0.044 (0.037, 0.052) * | 0.154 (0.147, 0.161) * | 0.143 (0.136, 0.151) * | 0.072 (0.065, 0.08) * | 0.011 (0.003, 0.018) | 0.248 (0.24, 0.255) * | 0.245 (0.238, 0.253) * |
| **HDL_C** | -0.164 (-0.172, -0.157) * | -0.052 (-0.059, -0.045) * | -0.493 (-0.5, -0.486) * | -0.48 (-0.487, -0.473) * | -0.129 (-0.136, -0.121) * | -0.045 (-0.052, -0.037) * | -0.347 (-0.354, -0.34) * | -0.336 (-0.343, -0.328) * |
| **IGF_1** | -0.007 (-0.014, 0) | 0.005 (-0.002, 0.012) | -0.049 (-0.057, -0.042) * | -0.051 (-0.058, -0.043) * | -0.046 (-0.054, -0.039) * | -0.026 (-0.033, -0.018) * | -0.09 (-0.098, -0.082) * | -0.083 (-0.091, -0.076) * |
| **LDL direct** | 0.105 (0.098, 0.113) * | 0.037 (0.03, 0.044) * | 0.302 (0.295, 0.309) * | 0.293 (0.285, 0.3) * | 0.067 (0.059, 0.075) * | 0.028 (0.02, 0.036) * | 0.161 (0.154, 0.169) * | 0.154 (0.146, 0.162) * |
| **Lipoprotein A** | 0.008 (-0.001, 0.016) | 0.025 (0.016, 0.034) * | -0.068 (-0.077, -0.059) * | -0.074 (-0.083, -0.065) * | -0.002 (-0.011, 0.007) | 0.005 (-0.004, 0.014) | -0.026 (-0.034, -0.017) * | -0.027 (-0.036, -0.018) * |
| **SHBG** | -0.16 (-0.168, -0.152) * | -0.082 (-0.089, -0.074) * | -0.356 (-0.363, -0.348) * | -0.336 (-0.343, -0.328) * | -0.128 (-0.136, -0.121) * | -0.05 (-0.058, -0.043) * | -0.324 (-0.332, -0.317) * | -0.311 (-0.319, -0.304) * |
| **Total bilirubin** | -0.04 (-0.048, -0.032) * | 0.004 (-0.004, 0.012) | -0.189 (-0.197, -0.181) * | -0.19 (-0.198, -0.182) * | -0.062 (-0.069, -0.054) * | -0.004 (-0.012, 0.004) | -0.231 (-0.239, -0.224) * | -0.23 (-0.238, -0.222) * |
| **Testosterone** | -0.02 (-0.028, -0.011) * | -0.018 (-0.027, -0.01) * | -0.01 (-0.018, -0.001) * | -0.005 (-0.014, 0.003) | -0.104 (-0.112, -0.097) * | -0.035 (-0.043, -0.027) * | -0.285 (-0.292, -0.277) * | -0.276 (-0.284, -0.268) * |
| **TG** | 0.229 (0.222, 0.237) * | 0.033 (0.029, 0.037) * | 0.85 (0.846, 0.854) * | 0.842 (0.838, 0.846) * | 0.178 (0.17, 0.186) * | 0.02 (0.014, 0.026) * | 0.635 (0.629, 0.641) * | 0.63 (0.623, 0.636) * |
| **Vitamin D** | -0.02 (-0.028, -0.012) * | 0.014 (0.006, 0.022) * | -0.149 (-0.157, -0.141) * | -0.152 (-0.16, -0.144) * | -0.095 (-0.103, -0.087) * | -0.057 (-0.065, -0.049) * | -0.169 (-0.177, -0.161) * | -0.154 (-0.162, -0.146) * |

**Supplementary Table S7.** Overall and sex-specific descriptives in UK Biobank and Epirus Health Study for 249 nuclear magnetic resonance metabolites. Data are presented as Mean ± SD and Median (25º, 75º). Abbreviations: ABSI, a body shape index; BMI, body mass index; -C, cholesterol; HI, hip index; -TG, triglycerides; -PL, Phospholipids; -CE, Cholesteryl esters; -FC, Free cholesterol; -L, Total lipids; -P, Lipoprotein particle concentrations; XXL-, Chylomicrons and extremely large; XL-, Very large; L-, Large; M-, Medium; S-, Small; XS- Very small; SD, standard deviation, LA, Linoleic Acid; DHA, docosahexaenoic acid; FA, fatty acids; HDL, high-density lipoprotein; IDL, intermediate-density lipoproteins; LDL, low-density lipoprotein; MUFA, monounsaturated fatty acid; PUFA, polyunsaturated fatty acid; SFA, saturated fatty acid; VLDL, very low-density lipoprotein; VHDL, very high-density lipoprotein.

| | | **UK Biobank** | | **Epirus Health Study** | |
|---|---|---|---|---|---|
| **Metabolites** | **Category** | **Female N = 80,978 (53.4%)** | **Male N = 70,623 (46.6%)** | **Females N = 615 (54.6%)** | **Males N = 512 (45.4%)** |
| Total Cholesterol | Cholesterol | 4,92 (4,35, 5,52) | 4,59 (4,06, 5,15) | 5.19 (4.58, 5.82) | 5.19 (4.53, 5.83) |
| Total Cholesterol Minus HDL, C | Cholesterol | 3,44 (2,93, 4,01) | 3,4 (2,91, 3,92) | 3.49 (2.94, 4.11) | 3.75 (3.19, 4.43) |
| Remnant Cholesterol (Non, HDL, Non, LDL , Cholesterol) | Cholesterol | 1,63 (1,37, 1,92) | 1,6 (1,37, 1,86) | 1.56 (1.31, 1.85) | 1.68 (1.39, 2.03) |
| VLDL Cholesterol | Cholesterol | 0,72 (0,56, 0,9) | 0,77 (0,63, 0,93) | 0.59 (0.46, 0.77) | 0.73 (0.56, 0.94) |
| Clinical LDL Cholesterol | Cholesterol | 2,68 (2,23, 3,16) | 2,63 (2,21, 3,09) | 2.85 (2.36, 3.33) | 3.06 (2.61, 3.6) |
| LDL Cholesterol | Cholesterol | 1,81 (1,54, 2,1) | 1,79 (1,54, 2,07) | 1.93 (1.63, 2.24) | 2.09 (1.79, 2.44) |
| HDL Cholesterol | Cholesterol | 1,43 (1,24, 1,65) | 1,15 (1,01, 1,33) | 1.68 (1.46, 1.89) | 1.35 (1.2, 1.52) |
| Total Triglycerides | Triglycerides | 1,11 (0,84, 1,49) | 1,38 (1,03, 1,81) | 0.89 (0.71, 1.14) | 1.2 (0.89, 1.66) |
| Triglycerides in VLDL | Triglycerides | 0,73 (0,52, 1,06) | 1 (0,7, 1,36) | 0.54 (0.41, 0.74) | 0.84 (0.58, 1.25) |
| Triglycerides in LDL | Triglycerides | 0,14 (0,12, 0,16) | 0,14 (0,12, 0,17) | 0.14 (0.12, 0.16) | 0.14 (0.12, 0.17) |
| Triglycerides in HDL | Triglycerides | 0,14 (0,11, 0,17) | 0,14 (0,11, 0,17) | 0.11 (0.09, 0.14) | 0.12 (0.1, 0.15) |
| Total Phospholipids in Lipoprotein Particles | Phospholipids | 3,08 (2,8, 3,38) | 2,85 (2,59, 3,14) | 3.17 (2.9, 3.49) | 3.1 (2.79, 3.39) |
| Phospholipids in VLDL | Phospholipids | 0,44 (0,34, 0,57) | 0,51 (0,4, 0,63) | 0.35 (0.27, 0.47) | 0.47 (0.35, 0.63) |
| Phospholipids in LDL | Phospholipids | 0,63 (0,54, 0,72) | 0,63 (0,55, 0,72) | 0.66 (0.57, 0.76) | 0.71 (0.62, 0.82) |
| Phospholipids in HDL | Phospholipids | 1,66 (1,47, 1,87) | 1,4 (1,24, 1,57) | 1.83 (1.61, 2.03) | 1.53 (1.4, 1.7) |
| Total Esterified Cholesterol | Cholesteryl esters | 3,58 (3,18, 4,01) | 3,33 (2,95, 3,72) | 3.8 (3.39, 4.27) | 3.8 (3.34, 4.26) |
| Cholesteryl Esters in VLDL | Cholesteryl esters | 0,44 (0,35, 0,54) | 0,46 (0,37, 0,54) | 0.37 (0.29, 0.47) | 0.44 (0.35, 0.56) |
| Cholesteryl Esters in LDL | Cholesteryl esters | 1,32 (1,12, 1,54) | 1,32 (1,13, 1,53) | 1.39 (1.17, 1.63) | 1.53 (1.31, 1.8) |
| Cholesteryl Esters in HDL | Cholesteryl esters | 1,11 (0,96, 1,29) | 0,89 (0,78, 1,04) | 1.31 (1.14, 1.47) | 1.06 (0.93, 1.2) |
| Total Free Cholesterol | Free cholesterol | 1,34 (1,18, 1,51) | 1,27 (1,11, 1,43) | 1.37 (1.2, 1.56) | 1.39 (1.21, 1.6) |
| Free Cholesterol in VLDL | Free cholesterol | 0,28 (0,21, 0,36) | 0,31 (0,25, 0,39) | 0.22 (0.17, 0.29) | 0.29 (0.22, 0.38) |
| Free Cholesterol in LDL | Free cholesterol | 0,49 (0,42, 0,57) | 0,47 (0,4, 0,55) | 0.54 (0.46, 0.62) | 0.56 (0.49, 0.65) |
| Free Cholesterol in HDL | Free cholesterol | 0,32 (0,28, 0,37) | 0,26 (0,23, 0,29) | 0.37 (0.32, 0.42) | 0.3 (0.26, 0.33) |
| Total Lipids in Lipoprotein Particles | Total lipids | 9,19 (8,19, 10,28) | 8,87 (7,91, 9,93) | 9.31 (8.33, 10.41) | 9.53 (8.36, 10.85) |
| Total Lipids in VLDL | Total lipids | 1,91 (1,44, 2,52) | 2,29 (1,76, 2,9) | 1.5 (1.15, 1.98) | 2.03 (1.5, 2.8) |
| Total Lipids in LDL | Total lipids | 2,58 (2,21, 2,98) | 2,57 (2,22, 2,96) | 2.71 (2.33, 3.15) | 2.93 (2.54, 3.44) |
| Total Lipids in HDL | Total lipids | 3,23 (2,85, 3,66) | 2,7 (2,4, 3,05) | 3.62 (3.21, 4.04) | 3.02 (2.73, 3.34) |
| Total Concentration of Lipoprotein Particles | Lipoprotein particle concentrations | 0,02 (0,02, 0,02) | 0,02 (0,01, 0,02) | 0.02 (0.02, 0.02) | 0.02 (0.02, 0.02) |
| Concentration of VLDL Particles | Lipoprotein particle concentrations | <0.1 (<0.1, <0.1) | <0.1 (<0.1, <0.1) | <0.01 (<0.01, <0.01) | <0.01 (<0.01, <0.01) |
| Concentration of LDL Particles | Lipoprotein particle concentrations | <0.1 (<0.1, <0.1) | <0.1 (<0.1, <0.1) | <0.01 (<0.01, <0.01) | <0.01 (<0.01, <0.01) |
| Concentration of HDL Particles | Lipoprotein particle concentrations | 0,02 (0,01, 0,02) | 0,01 (0,01, 0,02) | 0.02 (0.02, 0.02) | 0.02 (0.01, 0.02) |
| Average Diameter for VLDL Particles | Lipoprotein particle sizes | 38,2 (37,49, 39) | 39,17 (38,37, 39,92) | 37.61 (37.01, 38.23) | 38.73 (37.88, 39.77) |
| Average Diameter for LDL Particles | Lipoprotein particle sizes | 23,96 (23,9, 24,01) | 23,92 (23,86, 23,98) | 23.98 (23.91, 24.03) | 23.95 (23.89, 24) |
| Average Diameter for HDL Particles | Lipoprotein particle sizes | 9,71 (9,58, 9,85) | 9,52 (9,44, 9,63) | 9.81 (9.68, 9.94) | 9.57 (9.49, 9.69) |
| Phosphoglycerides | Other lipids | 2,41 (2,17, 2,66) | 2,19 (1,97, 2,43) | 2.51 (2.28, 2.79) | 2.43 (2.17, 2.69) |
| Triglycerides to Phosphoglycerides ratio | Other lipids | 0,47 (0,36, 0,61) | 0,64 (0,49, 0,81) | 0.36 (0.29, 0.44) | 0.5 (0.39, 0.66) |
| Total Cholines | Other lipids | 2,71 (2,47, 2,97) | 2,47 (2,25, 2,71) | 2.86 (2.62, 3.14) | 2.76 (2.49, 3.02) |
| Phosphatidylcholines | Other lipids | 2,23 (2, 2,47) | 2 (1,8, 2,22) | 2.35 (2.13, 2.6) | 2.23 (1.99, 2.46) |
| Sphingomyelins | Other lipids | 0,48 (0,43, 0,52) | 0,44 (0,4, 0,48) | 0.52 (0.47, 0.56) | 0.49 (0.45, 0.53) |
| Apolipoprotein B | Apolipoproteins | 0,87 (0,75, 1,01) | 0,88 (0,76, 1,01) | 0.86 (0.74, 1) | 0.95 (0.8, 1.12) |
| Apolipoprotein A1 | Apolipoproteins | 1,53 (1,39, 1,69) | 1,35 (1,23, 1,48) | 1.67 (1.51, 1.82) | 1.47 (1.37, 1.6) |
| Apolipoprotein B to Apolipoprotein A1 ratio | Apolipoproteins | 0,57 (0,47, 0,68) | 0,65 (0,55, 0,77) | 0.51 (0.43, 0.62) | 0.64 (0.53, 0.77) |
| Total Fatty Acids | Fatty acids | 12,06 (10,71, 13,63) | 11,92 (10,51, 13,57) | 11.95 (10.71, 13.43) | 12.39 (10.94, 14.35) |
| Degree of Unsaturation | Fatty acids | 1,37 (1,33, 1,42) | 1,33 (1,28, 1,38) | 1.36 (1.33, 1.39) | 1.33 (1.3, 1.37) |
| Omega, 3 Fatty Acids | Fatty acids | 0,52 (0,4, 0,68) | 0,46 (0,35, 0,6) | 0.38 (0.3, 0.48) | 0.4 (0.3, 0.5) |

| | | | | | |
|---|---|---|---|---|---|
| Omega, 6 Fatty Acids | Fatty acids | 4,65 (4,24, 5,1) | 4,47 (4,07, 4,89) | 4.82 (4.39, 5.35) | 4.87 (4.4, 5.45) |
| Polyunsaturated Fatty Acids | Fatty acids | 5,2 (4,7, 5,74) | 4,95 (4,49, 5,46) | 5.23 (4.74, 5.78) | 5.3 (4.75, 5.94) |
| Monounsaturated Fatty Acids | Fatty acids | 2,75 (2,34, 3,26) | 2,87 (2,4, 3,47) | 2.68 (2.34, 3.12) | 2.93 (2.47, 3.55) |
| Saturated Fatty Acids | Fatty acids | 4,07 (3,57, 4,67) | 4,04 (3,5, 4,72) | 4.01 (3.58, 4.51) | 4.14 (3.6, 4.8) |
| Linoleic Acid | Fatty acids | 3,59 (3,19, 4,04) | 3,45 (3,06, 3,88) | 3.83 (3.43, 4.39) | 3.88 (3.46, 4.51) |
| Docosahexaenoic Acid | Fatty acids | 0,24 (0,2, 0,3) | 0,2 (0,16, 0,25) | 0.21 (0.18, 0.24) | 0.2 (0.17, 0.23) |
| Omega, 3 Fatty Acids to Total Fatty Acids % | Fatty acids | 4,27 (3,43, 5,27) | 3,84 (3,1, 4,71) | 3.17 (2.62, 3.82) | 3.12 (2.61, 3.76) |
| Omega, 6 Fatty Acids to Total Fatty Acids % | Fatty acids | 38,91 (36,81, 40,63) | 37,89 (35,13, 40,12) | 40.61 (39.3, 41.76) | 39.74 (37.69, 41.17) |
| Polyunsaturated Fatty Acids to Total Fatty Acids % | Fatty acids | 43,37 (41,27, 45,13) | 41,93 (39,11, 44,23) | 43.84 (42.85, 44.92) | 42.92 (41.09, 44.32) |
| Monounsaturated Fatty Acids to Total Fatty Acids % | Fatty acids | 22,86 (21,51, 24,43) | 24,12 (22,49, 26,03) | 22.43 (21.57, 23.58) | 23.64 (22.47, 25.28) |
| Saturated Fatty Acids to Total Fatty Acids % | Fatty acids | 33,84 (32,83, 34,94) | 34 (32,78, 35,39) | 33.64 (32.97, 34.44) | 33.5 (32.62, 34.28) |
| Linoleic Acid to Total Fatty Acids % | Fatty acids | 29,88 (27,92, 31,72) | 29,12 (26,86, 31,2) | 32.32 (30.81, 33.81) | 31.63 (30.15, 33.1) |
| Docosahexaenoic Acid to Total Fatty Acids % | Fatty acids | 2,01 (1,66, 2,41) | 1,73 (1,39, 2,12) | 1.8 (1.58, 2.02) | 1.6 (1.37, 1.83) |
| Polyunsaturated Fatty Acids to Monounsaturated Fatty Acids ratio | Fatty acids | 1,9 (1,7, 2,09) | 1,74 (1,51, 1,96) | 1.96 (1.83, 2.07) | 1.82 (1.64, 1.97) |
| Omega, 6 Fatty Acids to Omega, 3 Fatty Acids ratio | Fatty acids | 8,98 (7,16, 11,38) | 9,69 (7,74, 12,22) | 12.79 (10.43, 15.68) | 12.59 (10.37, 15.41) |
| Alanine | Amino acids | 0,28 (0,23, 0,33) | 0,29 (0,25, 0,35) | 0.32 (0.28, 0.36) | 0.34 (0.3, 0.39) |
| Glutamine | Amino acids | 0,55 (0,49, 0,6) | 0,56 (0,51, 0,61) | 0.61 (0.55, 0.67) | 0.64 (0.58, 0.7) |
| Glycine | Amino acids | 0,18 (0,14, 0,24) | 0,14 (0,12, 0,17) | 0.26 (0.22, 0.31) | 0.22 (0.2, 0.25) |
| Histidine | Amino acids | 0,06 (0,06, 0,07) | 0,07 (0,06, 0,07) | 0.08 (0.07, 0.08) | 0.08 (0.07, 0.09) |
| Total Concentration of Branched, Chain Amino Acids (Leucine + Isoleucine + Valine) | Amino acids | 0,33 (0,29, 0,38) | 0,38 (0,33, 0,43) | 0.35 (0.31, 0.41) | 0.44 (0.39, 0.49) |
| Isoleucine | Amino acids | 0,04 (0,04, 0,06) | 0,05 (0,04, 0,06) | 0.04 (0.04, 0.05) | 0.06 (0.05, 0.07) |
| Leucine | Amino acids | 0,09 (0,08, 0,11) | 0,11 (0,09, 0,13) | 0.1 (0.09, 0.12) | 0.13 (0.11, 0.15) |
| Valine | Amino acids | 0,19 (0,17, 0,22) | 0,22 (0,19, 0,24) | 0.21 (0.18, 0.23) | 0.25 (0.22, 0.28) |
| Phenylalanine | Amino acids | 0,05 (0,04, 0,05) | 0,05 (0,04, 0,05) | 0.06 (0.06, 0.07) | 0.07 (0.06, 0.08) |
| Tyrosine | Amino acids | 0,06 (0,05, 0,07) | 0,06 (0,05, 0,07) | 0.06 (0.05, 0.06) | 0.06 (0.06, 0.07) |
| Glucose | Glycolysis related metabolites | 3,54 (3,12, 3,97) | 3,48 (3,05, 3,92) | 4.45 (4.06, 4.82) | 4.68 (4.26, 5) |
| Lactate | Glycolysis related metabolites | 3,74 (3,09, 4,49) | 3,9 (3,25, 4,64) | 2.19 (1.84, 2.64) | 2.44 (2.05, 3) |
| Pyruvate | Glycolysis related metabolites | 0,08 (0,06, 0,1) | 0,08 (0,06, 0,09) | 0.03 (0.02, 0.04) | 0.03 (0.02, 0.04) |
| Citrate | Glycolysis related metabolites | 0,07 (0,06, 0,07) | 0,06 (0,05, 0,07) | 0.06 (0.06, 0.07) | 0.06 (0.05, 0.07) |
| 3, Hydroxybutyrate | Ketone bodies | 0,04 (0,03, 0,07) | 0,04 (0,03, 0,06) | 0.02 (0.02, 0.03) | 0.02 (0.01, 0.03) |
| Acetate | Ketone bodies | 0,02 (0,01, 0,02) | 0,02 (0,01, 0,02) | 0.02 (0.01, 0.04) | 0.02 (0.01, 0.04) |
| Acetoacetate | Ketone bodies | 0,01 (0,01, 0,02) | 0,01 (0,01, 0,02) | 0.02 (0.01, 0.02) | 0.02 (0.01, 0.02) |
| Acetone | Ketone bodies | 0,01 (0,01, 0,02) | 0,01 (0,01, 0,02) | 62.97 (56.14, 69.3) | 77.69 (69.16, 86.9) |
| Creatinine | Fluid balance | 0,06 (0,05, 0,07) | 0,07 (0,07, 0,08) | 42.84 (40.72, 45.4) | 44.25 (41.93, 46.83) |
| Albumin | Fluid balance | 39,35 (37,35, 41,38) | 39,72 (37,71, 41,78) | 0.77 (0.7, 0.85) | 0.81 (0.73, 0.89) |
| Glycoprotein Acetyls | Inflammation | 0,8 (0,72, 0,88) | 0,8 (0,73, 0,88) | <0.01 (<0.01, <0.01) | <0.01 (<0.01, <0.01) |
| Concentration of Chylomicrons and Extremely Large VLDL Particles | Lipoprotein particle concentrations | <0.1 (<0.1, <0.1) | <0.1 (<0.1, <0.1) | 0.02 (0.01, 0.08) | 0.1 (0.03, 0.25) |
| Total Lipids in Chylomicrons and Extremely Large VLDL | Total lipids | 0,13 (0,05, 0,25) | 0,24 (0,12, 0,4) | <0.01 (<0.01, 0.01) | 0.01 (<0.01, 0.04) |
| Phospholipids in Chylomicrons and Extremely Large VLDL | Phospholipids | 0,02 (0,01, 0,04) | 0,04 (0,02, 0,06) | 0.01 (<0.01, 0.03) | 0.03 (0.01, 0.06) |
| Cholesterol in Chylomicrons and Extremely Large VLDL | Cholesterol | 0,04 (0,02, 0,07) | 0,06 (0,04, 0,09) | 0.01 (<0.01, 0.02) | 0.02 (0.01, 0.04) |
| Cholesteryl Esters in Chylomicrons and Extremely Large VLDL | Cholesteryl esters | 0,02 (0,01, 0,04) | 0,03 (0,02, 0,05) | <0.01 (<0.01, 0.01) | 0.01 (<0.01, 0.03) |
| Free Cholesterol in Chylomicrons and Extremely Large VLDL | Free cholesterol | 0,02 (0,01, 0,03) | 0,03 (0,01, 0,04) | 0.01 (<0.01, 0.04) | 0.05 (0.01, 0.16) |
| Triglycerides in Chylomicrons and Extremely Large VLDL | Triglycerides | 0,07 (0,03, 0,15) | 0,14 (0,06, 0,24) | <0.01 (<0.01, <0.01) | <0.01 (<0.01, <0.01) |
| Concentration of Very Large VLDL Particles | Lipoprotein particle concentrations | <0.1 (<0.1, <0.1) | <0.1 (<0.1, <0.1) | 0.09 (0.05, 0.15) | 0.18 (0.11, 0.3) |
| Total Lipids in Very Large VLDL | Total lipids | 0,15 (0,09, 0,24) | 0,23 (0,15, 0,33) | 0.02 (0.01, 0.03) | 0.03 (0.02, 0.05) |
| Phospholipids in Very Large VLDL | Phospholipids | 0,03 (0,02, 0,05) | 0,04 (0,03, 0,06) | 0.03 (0.02, 0.05) | 0.05 (0.03, 0.08) |
| Cholesterol in Very Large VLDL | Cholesterol | 0,05 (0,03, 0,07) | 0,06 (0,04, 0,08) | 0.02 (0.01, 0.03) | 0.03 (0.02, 0.04) |
| Cholesteryl Esters in Very Large VLDL | Cholesteryl esters | 0,03 (0,02, 0,04) | 0,03 (0,03, 0,04) | 0.01 (0.01, 0.02) | 0.02 (0.01, 0.03) |
| Free Cholesterol in Very Large VLDL | Free cholesterol | 0,02 (0,01, 0,03) | 0,03 (0,02, 0,04) | 0.04 (0.02, 0.07) | 0.09 (0.06, 0.16) |
| Triglycerides in Very Large VLDL | Triglycerides | 0,08 (0,04, 0,13) | 0,12 (0,07, 0,19) | <0.01 (<0.01, <0.01) | <0.01 (<0.01, <0.01) |
| Concentration of Large VLDL Particles | Lipoprotein particle concentrations | <0.1 (<0.1, <0.1) | <0.1 (<0.1, <0.1) | 0.2 (0.13, 0.29) | 0.34 (0.22, 0.5) |
| Total Lipids in Large VLDL | Total lipids | 0,27 (0,18, 0,4) | 0,38 (0,27, 0,5) | 0.03 (0.02, 0.06) | 0.06 (0.04, 0.1) |
| Phospholipids in Large VLDL | Phospholipids | 0,05 (0,03, 0,08) | 0,08 (0,05, 0,1) | 0.06 (0.04, 0.09) | 0.1 (0.07, 0.14) |
| Cholesterol in Large VLDL | Cholesterol | 0,09 (0,06, 0,12) | 0,11 (0,08, 0,14) | 0.03 (0.02, 0.05) | 0.05 (0.04, 0.07) |

| | | | | | |
|---|---|---|---|---|---|
| Cholesteryl Esters in Large VLDL | Cholesteryl esters | 0,05 (0,03, 0,07) | 0,06 (0,04, 0,07) | 0.03 (0.02, 0.04) | 0.05 (0.03, 0.07) |
| Free Cholesterol in Large VLDL | Free cholesterol | 0,04 (0,03, 0,06) | 0,05 (0,04, 0,07) | 0.1 (0.07, 0.15) | 0.17 (0.11, 0.27) |
| Triglycerides in Large VLDL | Triglycerides | 0,13 (0,09, 0,2) | 0,19 (0,13, 0,26) | <0.01 (<0.01, <0.01) | <0.01 (<0.01, <0.01) |
| Concentration of Medium VLDL Particles | Lipoprotein particle concentrations | <0.1 (<0.1, <0.1) | <0.1 (<0.1, <0.1) | 0.5 (0.38, 0.64) | 0.64 (0.49, 0.84) |
| Total Lipids in Medium VLDL | Total lipids | 0,56 (0,43, 0,71) | 0,63 (0,5, 0,77) | 0.11 (0.09, 0.15) | 0.14 (0.11, 0.19) |
| Phospholipids in Medium VLDL | Phospholipids | 0,13 (0,1, 0,16) | 0,14 (0,11, 0,17) | 0.16 (0.13, 0.21) | 0.19 (0.15, 0.25) |
| Cholesterol in Medium VLDL | Cholesterol | 0,18 (0,14, 0,23) | 0,18 (0,14, 0,22) | 0.1 (0.07, 0.12) | 0.1 (0.08, 0.13) |
| Cholesteryl Esters in Medium VLDL | Cholesteryl esters | 0,1 (0,08, 0,13) | 0,1 (0,07, 0,12) | 0.07 (0.05, 0.09) | 0.09 (0.07, 0.11) |
| Free Cholesterol in Medium VLDL | Free cholesterol | 0,08 (0,06, 0,1) | 0,08 (0,07, 0,1) | 0.21 (0.16, 0.27) | 0.3 (0.22, 0.41) |
| Triglycerides in Medium VLDL | Triglycerides | 0,24 (0,18, 0,33) | 0,3 (0,23, 0,39) | <0.01 (<0.01, <0.01) | <0.01 (<0.01, <0.01) |
| Concentration of Small VLDL Particles | Lipoprotein particle concentrations | <0.1 (<0.1, <0.1) | <0.1 (<0.1, <0.1) | 0.34 (0.27, 0.43) | 0.43 (0.34, 0.54) |
| Total Lipids in Small VLDL | Total lipids | 0,4 (0,32, 0,5) | 0,44 (0,36, 0,53) | 0.09 (0.07, 0.11) | 0.1 (0.08, 0.13) |
| Phospholipids in Small VLDL | Phospholipids | 0,1 (0,08, 0,12) | 0,1 (0,09, 0,12) | 0.13 (0.1, 0.17) | 0.17 (0.13, 0.2) |
| Cholesterol in Small VLDL | Cholesterol | 0,16 (0,12, 0,2) | 0,17 (0,14, 0,2) | 0.08 (0.06, 0.1) | 0.1 (0.08, 0.12) |
| Cholesteryl Esters in Small VLDL | Cholesteryl esters | 0,1 (0,08, 0,12) | 0,1 (0,09, 0,13) | 0.05 (0.04, 0.07) | 0.07 (0.05, 0.08) |
| Free Cholesterol in Small VLDL | Free cholesterol | 0,06 (0,05, 0,07) | 0,06 (0,05, 0,07) | 0.11 (0.09, 0.15) | 0.15 (0.11, 0.2) |
| Triglycerides in Small VLDL | Triglycerides | 0,14 (0,11, 0,18) | 0,17 (0,13, 0,21) | <0.01 (<0.01, <0.01) | <0.01 (<0.01, <0.01) |
| Concentration of Very Small VLDL Particles | Lipoprotein particle concentrations | <0.1 (<0.1, <0.1) | <0.1 (<0.1, <0.1) | 0.34 (0.28, 0.4) | 0.35 (0.29, 0.41) |
| Total Lipids in Very Small VLDL | Total lipids | 0,37 (0,32, 0,44) | 0,35 (0,31, 0,41) | 0.1 (0.08, 0.12) | 0.1 (0.08, 0.12) |
| Phospholipids in Very Small VLDL | Phospholipids | 0,11 (0,09, 0,13) | 0,1 (0,09, 0,12) | 0.18 (0.15, 0.22) | 0.18 (0.15, 0.22) |
| Cholesterol in Very Small VLDL | Cholesterol | 0,2 (0,17, 0,23) | 0,18 (0,15, 0,21) | 0.13 (0.11, 0.15) | 0.13 (0.1, 0.15) |
| Cholesteryl Esters in Very Small VLDL | Cholesteryl esters | 0,14 (0,12, 0,16) | 0,12 (0,1, 0,14) | 0.05 (0.05, 0.06) | 0.06 (0.05, 0.07) |
| Free Cholesterol in Very Small VLDL | Free cholesterol | 0,06 (0,05, 0,07) | 0,06 (0,05, 0,07) | 0.06 (0.05, 0.07) | 0.06 (0.05, 0.08) |
| Triglycerides in Very Small VLDL | Triglycerides | 0,07 (0,05, 0,08) | 0,07 (0,06, 0,08) | <0.01 (<0.01, <0.01) | <0.01 (<0.01, <0.01) |
| Concentration of IDL Particles | Lipoprotein particle concentrations | <0.1 (<0.1, <0.1) | <0.1 (<0.1, <0.1) | 1.37 (1.18, 1.58) | 1.35 (1.16, 1.56) |
| Total Lipids in IDL | Total lipids | 1,32 (1,15, 1,51) | 1,22 (1,06, 1,39) | 0.32 (0.28, 0.37) | 0.32 (0.27, 0.37) |
| Phospholipids in IDL | Phospholipids | 0,31 (0,27, 0,36) | 0,29 (0,25, 0,33) | 0.95 (0.82, 1.1) | 0.93 (0.8, 1.08) |
| Cholesterol in IDL | Cholesterol | 0,91 (0,78, 1,04) | 0,83 (0,71, 0,95) | 0.71 (0.61, 0.82) | 0.7 (0.6, 0.82) |
| Cholesteryl Esters in IDL | Cholesteryl esters | 0,67 (0,58, 0,77) | 0,61 (0,53, 0,7) | 0.24 (0.21, 0.28) | 0.24 (0.2, 0.27) |
| Free Cholesterol in IDL | Free cholesterol | 0,24 (0,2, 0,27) | 0,22 (0,19, 0,25) | 0.09 (0.08, 0.11) | 0.09 (0.08, 0.11) |
| Triglycerides in IDL | Triglycerides | 0,1 (0,08, 0,11) | 0,1 (0,08, 0,12) | <0.01 (<0.01, <0.01) | <0.01 (<0.01, <0.01) |
| Concentration of Large LDL Particles | Lipoprotein particle concentrations | <0.1 (<0.1, <0.1) | <0.1 (<0.1, <0.1) | 1.78 (1.52, 2.05) | 1.87 (1.61, 2.15) |
| Total Lipids in Large LDL | Total lipids | 1,66 (1,43, 1,91) | 1,61 (1,4, 1,85) | 0.39 (0.34, 0.45) | 0.41 (0.36, 0.47) |
| Phospholipids in Large LDL | Phospholipids | 0,37 (0,32, 0,43) | 0,36 (0,32, 0,42) | 1.29 (1.1, 1.49) | 1.36 (1.16, 1.58) |
| Cholesterol in Large LDL | Cholesterol | 1,19 (1,02, 1,38) | 1,15 (0,99, 1,33) | 0.95 (0.8, 1.1) | 1.01 (0.86, 1.17) |
| Cholesteryl Esters in Large LDL | Cholesteryl esters | 0,88 (0,75, 1,02) | 0,85 (0,73, 0,99) | 0.35 (0.29, 0.4) | 0.35 (0.3, 0.4) |
| Free Cholesterol in Large LDL | Free cholesterol | 0,32 (0,27, 0,36) | 0,3 (0,25, 0,35) | 0.09 (0.08, 0.11) | 0.1 (0.08, 0.11) |
| Triglycerides in Large LDL | Triglycerides | 0,09 (0,08, 0,11) | 0,1 (0,08, 0,11) | <0.01 (<0.01, <0.01) | <0.01 (<0.01, <0.01) |
| Concentration of Medium LDL Particles | Lipoprotein particle concentrations | <0.1 (<0.1, <0.1) | <0.1 (<0.1, <0.1) | 0.65 (0.54, 0.77) | 0.75 (0.64, 0.89) |
| Total Lipids in Medium LDL | Total lipids | 0,63 (0,52, 0,74) | 0,66 (0,56, 0,77) | 0.17 (0.15, 0.2) | 0.2 (0.17, 0.23) |
| Phospholipids in Medium LDL | Phospholipids | 0,16 (0,14, 0,19) | 0,17 (0,15, 0,2) | 0.44 (0.37, 0.53) | 0.52 (0.44, 0.61) |
| Cholesterol in Medium LDL | Cholesterol | 0,43 (0,36, 0,51) | 0,45 (0,38, 0,53) | 0.31 (0.25, 0.38) | 0.37 (0.31, 0.45) |
| Cholesteryl Esters in Medium LDL | Cholesteryl esters | 0,31 (0,25, 0,37) | 0,33 (0,27, 0,39) | 0.13 (0.11, 0.16) | 0.15 (0.13, 0.17) |
| Free Cholesterol in Medium LDL | Free cholesterol | 0,12 (0,1, 0,15) | 0,12 (0,11, 0,14) | 0.03 (0.03, 0.03) | 0.03 (0.03, 0.04) |
| Triglycerides in Medium LDL | Triglycerides | 0,03 (0,03, 0,04) | 0,03 (0,03, 0,04) | <0.01 (<0.01, <0.01) | <0.01 (<0.01, <0.01) |
| Concentration of Small LDL Particles | Lipoprotein particle concentrations | <0.1 (<0.1, <0.1) | <0.1 (<0.1, <0.1) | 0.3 (0.26, 0.35) | 0.33 (0.29, 0.39) |
| Total Lipids in Small LDL | Total lipids | 0,29 (0,25, 0,33) | 0,3 (0,26, 0,34) | 0.1 (0.08, 0.11) | 0.1 (0.09, 0.12) |
| Phospholipids in Small LDL | Phospholipids | 0,09 (0,08, 0,1) | 0,09 (0,08, 0,1) | 0.19 (0.16, 0.22) | 0.21 (0.19, 0.25) |
| Cholesterol in Small LDL | Cholesterol | 0,18 (0,16, 0,21) | 0,19 (0,16, 0,22) | 0.14 (0.12, 0.16) | 0.15 (0.13, 0.18) |
| Cholesteryl Esters in Small LDL | Cholesteryl esters | 0,13 (0,11, 0,15) | 0,14 (0,12, 0,16) | 0.06 (0.05, 0.07) | 0.06 (0.05, 0.07) |
| Free Cholesterol in Small LDL | Free cholesterol | 0,05 (0,04, 0,06) | 0,05 (0,04, 0,06) | 0.01 (0.01, 0.01) | 0.01 (0.01, 0.02) |
| Triglycerides in Small LDL | Triglycerides | 0,01 (0,01, 0,02) | 0,02 (0,01, 0,02) | <0.01 (<0.01, <0.01) | <0.01 (<0.01, <0.01) |
| Concentration of Very Large HDL Particles | Lipoprotein particle concentrations | <0.1 (<0.1, <0.1) | <0.1 (<0.1, <0.1) | 0.23 (0.18, 0.29) | 0.15 (0.12, 0.18) |
| Total Lipids in Very Large HDL | Total lipids | 0,18 (0,14, 0,24) | 0,13 (0,1, 0,16) | 0.11 (0.08, 0.15) | 0.06 (0.05, 0.08) |
| Phospholipids in Very Large HDL | Phospholipids | 0,09 (0,06, 0,12) | 0,05 (0,04, 0,07) | 0.11 (0.09, 0.14) | 0.08 (0.06, 0.09) |
| Cholesterol in Very Large HDL | Cholesterol | 0,09 (0,07, 0,11) | 0,06 (0,05, 0,08) | 0.08 (0.07, 0.1) | 0.05 (0.04, 0.07) |
| Cholesteryl Esters in Very Large HDL | Cholesteryl esters | 0,06 (0,05, 0,08) | 0,04 (0,04, 0,06) | 0.03 (0.02, 0.03) | 0.02 (0.02, 0.02) |
| Free Cholesterol in Very Large HDL | Free cholesterol | 0,02 (0,02, 0,03) | 0,02 (0,02, 0,02) | 0.01 (0.01, 0.01) | 0.01 (0, 0.01) |
| Triglycerides in Very Large HDL | Triglycerides | 0,01 (0,01, 0,01) | 0,01 (0,01, 0,01) | <0.01 (<0.01, <0.01) | <0.01 (<0.01, <0.01) |
| Concentration of Large HDL Particles | Lipoprotein particle concentrations | <0.1 (<0.1, <0.1) | <0.1 (<0.1, <0.1) | 0.97 (0.75, 1.18) | 0.57 (0.45, 0.76) |
| Total Lipids in Large HDL | Total lipids | 0,74 (0,55, 0,98) | 0,45 (0,34, 0,61) | 0.48 (0.37, 0.57) | 0.29 (0.23, 0.37) |
| Phospholipids in Large HDL | Phospholipids | 0,37 (0,28, 0,48) | 0,23 (0,18, 0,31) | 0.47 (0.36, 0.58) | 0.26 (0.2, 0.36) |
| Cholesterol in Large HDL | Cholesterol | 0,34 (0,24, 0,47) | 0,19 (0,14, 0,28) | 0.37 (0.27, 0.45) | 0.2 (0.16, 0.28) |
| Cholesteryl Esters in Large HDL | Cholesteryl esters | 0,26 (0,19, 0,36) | 0,15 (0,11, 0,21) | 0.1 (0.08, 0.13) | 0.06 (0.04, 0.08) |
| Free Cholesterol in Large HDL | Free cholesterol | 0,08 (0,06, 0,11) | 0,05 (0,03, 0,06) | 0.03 (0.02, 0.04) | 0.02 (0.02, 0.03) |
| Triglycerides in Large HDL | Triglycerides | 0,03 (0,02, 0,04) | 0,03 (0,02, 0,03) | <0.01 (<0.01, <0.011) | <0.01 (<0.01, <0.01) |

| | | | | | |
|---|---|---|---|---|---|
| Concentration of Medium HDL Particles | Lipoprotein particle concentrations | <0.1 (<0.1, <0.1) | <0.1 (<0.1, <0.1) | 1.21 (1.07, 1.34) | 1.03 (0.94, 1.17) |
| Total Lipids in Medium HDL | Total lipids | 1,1 (0,97, 1,25) | 0,94 (0,82, 1,07) | 0.55 (0.49, 0.61) | 0.48 (0.44, 0.54) |
| Phospholipids in Medium HDL | Phospholipids | 0,51 (0,46, 0,58) | 0,44 (0,39, 0,5) | 0.62 (0.53, 0.69) | 0.51 (0.45, 0.58) |
| Cholesterol in Medium HDL | Cholesterol | 0,54 (0,46, 0,61) | 0,44 (0,38, 0,51) | 0.51 (0.44, 0.56) | 0.42 (0.37, 0.48) |
| Cholesteryl Esters in Medium HDL | Cholesteryl esters | 0,44 (0,38, 0,5) | 0,36 (0,31, 0,42) | 0.11 (0.09, 0.13) | 0.09 (0.08, 0.1) |
| Free Cholesterol in Medium HDL | Free cholesterol | 0,1 (0,08, 0,11) | 0,08 (0,06, 0,09) | 0.04 (0.03, 0.05) | 0.04 (0.03, 0.06) |
| Triglycerides in Medium HDL | Triglycerides | 0,05 (0,04, 0,06) | 0,05 (0,04, 0,06) | 0.01 (0.01, 0.01) | 0.01 (0.01, 0.01) |
| Concentration of Small HDL Particles | Lipoprotein particle concentrations | 0,01 (0,01, 0,01) | 0,01 (0,01, 0,01) | 1.19 (1.1, 1.29) | 1.23 (1.14, 1.31) |
| Total Lipids in Small HDL | Total lipids | 1,17 (1,07, 1,28) | 1,15 (1,06, 1,25) | 0.68 (0.62, 0.73) | 0.69 (0.64, 0.74) |
| Phospholipids in Small HDL | Phospholipids | 0,67 (0,61, 0,73) | 0,65 (0,6, 0,71) | 0.47 (0.44, 0.52) | 0.49 (0.46, 0.52) |
| Cholesterol in Small HDL | Cholesterol | 0,45 (0,41, 0,49) | 0,44 (0,41, 0,48) | 0.35 (0.32, 0.38) | 0.36 (0.34, 0.39) |
| Cholesteryl Esters in Small HDL | Cholesteryl esters | 0,33 (0,3, 0,37) | 0,33 (0,3, 0,36) | 0.12 (0.11, 0.13) | 0.12 (0.12, 0.13) |
| Free Cholesterol in Small HDL | Free cholesterol | 0,12 (0,11, 0,13) | 0,11 (0,1, 0,12) | 0.04 (0.03, 0.05) | 0.05 (0.04, 0.06) |
| Triglycerides in Small HDL | Triglycerides | 0,05 (0,04, 0,06) | 0,06 (0,05, 0,07) | 11.81 (1.48, 14.63) | 13.72 (11.84, 15.24) |
| Phospholipids to Total Lipids in Chylomicrons and Extremely Large VLDL % | Relative lipoprotein lipid concentration | 15,5 (13,88, 16,71) | 15,65 (14,76, 16,54) | 33.04 (25.21, 44.02) | 28.8 (23.62, 38.55) |
| Cholesterol to Total Lipids in Chylomicrons and Extremely Large VLDL % | Relative lipoprotein lipid concentration | 27,59 (23,9, 34,09) | 25,46 (22,76, 29,8) | 20.88 (15.15, 30.01) | 17.69 (13.95, 25.75) |
| Cholesteryl Esters to Total Lipids in Chylomicrons and Extremely Large VLDL % | Relative lipoprotein lipid concentration | 15,29 (12,64, 19,8) | 14,29 (12,33, 17,35) | 11.73 (9.15, 14.56) | 10.74 (9.39, 12.94) |
| Free Cholesterol to Total Lipids in Chylomicrons and Extremely Large VLDL % | Relative lipoprotein lipid concentration | 12,18 (10,86, 14,55) | 11,06 (10,2, 12,55) | 54.67 (44.2, 65.4) | 57.33 (49.22, 63.12) |
| Triglycerides to Total Lipids in Chylomicrons and Extremely Large VLDL % | Relative lipoprotein lipid concentration | 57,02 (50,25, 61,37) | 58,79 (54,16, 62,02) | 16.76 (14.06, 18.38) | 17.97 (16.75, 18.82) |
| Phospholipids to Total Lipids in Very Large VLDL % | Relative lipoprotein lipid concentration | 18,89 (17,77, 19,76) | 19 (18,29, 19,71) | 33.43 (28.76, 39.02) | 28.55 (24.76, 32.62) |
| Cholesterol to Total Lipids in Very Large VLDL % | Relative lipoprotein lipid concentration | 30,08 (26,2, 34,91) | 26,91 (23,78, 30,72) | 21.74 (17.83, 26.91) | 17.52 (14.1, 20.93) |
| Cholesteryl Esters to Total Lipids in Very Large VLDL % | Relative lipoprotein lipid concentration | 17,83 (14,65, 21,81) | 15,22 (12,71, 18,41) | 11.63 (10.55, 12.68) | 11.14 (10.48, 11.84) |
| Free Cholesterol to Total Lipids in Very Large VLDL % | Relative lipoprotein lipid concentration | 12,21 (11,37, 13,22) | 11,64 (10,95, 12,4) | 50.28 (44.2, 55.1) | 53.59 (49.58, 57.53) |
| Triglycerides to Total Lipids in Very Large VLDL % | Relative lipoprotein lipid concentration | 51,26 (46,31, 55,42) | 54,11 (50,05, 57,59) | 17.55 (14.79, 19.35) | 19.34 (18.09, 20.18) |
| Phospholipids to Total Lipids in Large VLDL % | Relative lipoprotein lipid concentration | 19,42 (17,58, 20,58) | 20,21 (19,32, 21,02) | 29.26 (26.37, 31.75) | 28.41 (26.28, 30.75) |
| Cholesterol to Total Lipids in Large VLDL % | Relative lipoprotein lipid concentration | 31,05 (28,5, 33,66) | 29,54 (27,33, 31,98) | 16.26 (14.05, 18.16) | 15 (13.26, 16.75) |
| Cholesteryl Esters to Total Lipids in Large VLDL % | Relative lipoprotein lipid concentration | 17,2 (15,17, 19,31) | 15,46 (13,66, 17,48) | 13.08 (11.99, 13.96) | 13.47 (12.95, 14.08) |
| Free Cholesterol to Total Lipids in Large VLDL % | Relative lipoprotein lipid concentration | 13,93 (13,17, 14,64) | 14,11 (13,55, 14,69) | 53.71 (49.65, 58.38) | 52.52 (49.92, 54.95) |
| Triglycerides to Total Lipids in Large VLDL % | Relative lipoprotein lipid concentration | 49,95 (46,86, 53,14) | 50,43 (47,82, 52,87) | 23.05 (21.94, 23.97) | 22.3 (21.46, 23.33) |
| Phospholipids to Total Lipids in Medium VLDL % | Relative lipoprotein lipid concentration | 22,94 (21,95, 23,86) | 22,22 (21,23, 23,17) | 34.24 (30.4, 37.22) | 30.7 (26.73, 33.87) |
| Cholesterol to Total Lipids in Medium VLDL % | Relative lipoprotein lipid concentration | 32,83 (28,89, 36,48) | 29,28 (25,24, 33,21) | 19.81 (17.22, 21.91) | 17.01 (13.84, 19.43) |
| Cholesteryl Esters to Total Lipids in Medium VLDL % | Relative lipoprotein lipid concentration | 18,41 (15,42, 21,19) | 15,66 (12,63, 18,65) | 14.45 (13.4, 15.26) | 13.59 (12.78, 14.51) |
| Free Cholesterol to Total Lipids in Medium VLDL % | Relative lipoprotein lipid concentration | 14,41 (13,4, 15,33) | 13,61 (12,56, 14,58) | 42.87 (38.86, 47.45) | 47.04 (42.79, 51.75) |
| Triglycerides to Total Lipids in Medium VLDL % | Relative lipoprotein lipid concentration | 44,21 (39,73, 49,08) | 48,5 (43,68, 53,49) | 25.46 (24.26, 26.53) | 24.67 (23.59, 25.87) |
| Phospholipids to Total Lipids in Small VLDL % | Relative lipoprotein lipid concentration | 24,31 (23,18, 25,54) | 23,55 (22,41, 24,69) | 40.32 (37.31, 42.68) | 39.08 (36.14, 41.44) |
| Cholesterol to Total Lipids in Small VLDL % | Relative lipoprotein lipid concentration | 39,45 (36,66, 42,09) | 38,19 (35,28, 40,93) | 24.09 (22.22, 25.46) | 23.55 (21.84, 25.21) |
| Cholesteryl Esters to Total Lipids in Small VLDL % | Relative lipoprotein lipid concentration | 24,32 (22,59, 25,92) | 23,93 (22,2, 25,58) | 16.15 (14.88, 17.31) | 15.33 (14.23, 16.48) |
| Free Cholesterol to Total Lipids in Small VLDL % | Relative lipoprotein lipid concentration | 15,06 (13,86, 16,32) | 14,26 (12,99, 15,44) | 34.21 (30.74, 38.46) | 36.17 (32.6, 40.14) |
| Triglycerides to Total Lipids in Small VLDL % | Relative lipoprotein lipid concentration | 36,22 (32,45, 40,06) | 38,25 (34,43, 42,27) | 28.74 (28.11, 29.34) | 28.9 (28.23, 29.42) |
| Phospholipids to Total Lipids in Very Small VLDL % | Relative lipoprotein lipid concentration | 28,99 (28,51, 29,51) | 29,28 (28,76, 29,83) | 54.19 (52.13, 56) | 52.7 (50.61, 54.94) |
| Cholesterol to Total Lipids in Very Small VLDL % | Relative lipoprotein lipid concentration | 53,3 (50,93, 55,4) | 51,44 (48,47, 53,86) | 38.26 (36.44, 39.84) | 36.74 (34.67, 38.79) |
| Cholesteryl Esters to Total Lipids in Very Small VLDL % | Relative lipoprotein lipid concentration | 37,09 (34,94, 39,01) | 35,24 (32,59, 37,44) | 15.92 (15.68, 16.17) | 16.01 (15.74, 16.28) |

| | | | | | |
|---|---|---|---|---|---|
| Free Cholesterol to Total Lipids in Very Small VLDL % | Relative lipoprotein lipid concentration | 16,22 (15,89, 16,5) | 16,21 (15,79, 16,51) | 17.18 (15.54, 18.94) | 18.49 (16.57, 20.3) |
| Triglycerides to Total Lipids in Very Small VLDL % | Relative lipoprotein lipid concentration | 17,73 (15,89, 19,78) | 19,29 (17,19, 21,89) | 23.53 (22.95, 24.05) | 23.65 (23.11, 24.08) |
| Phospholipids to Total Lipids in IDL % | Relative lipoprotein lipid concentration | 23,8 (23,19, 24,38) | 23,8 (23,24, 24,35) | 69.82 (68.69, 70.73) | 69.34 (68.17, 70.43) |
| Cholesterol to Total Lipids in IDL % | Relative lipoprotein lipid concentration | 68,88 (67,23, 70,24) | 68,3 (66,4, 69,73) | 51.99 (50.78, 52.99) | 51.94 (50.77, 52.99) |
| Cholesteryl Esters to Total Lipids in IDL % | Relative lipoprotein lipid concentration | 50,83 (49,47, 52,03) | 50,33 (48,84, 51,55) | 17.76 (17.23, 18.27) | 17.38 (16.87, 17.82) |
| Free Cholesterol to Total Lipids in IDL % | Relative lipoprotein lipid concentration | 17,92 (17,3, 18,52) | 17,85 (17,07, 18,53) | 6.84 (6.15, 7.72) | 7.09 (6.27, 8.06) |
| Triglycerides to Total Lipids in IDL % | Relative lipoprotein lipid concentration | 7,39 (6,45, 8,54) | 7,95 (6,88, 9,42) | 22.01 (21.67, 22.37) | 22.07 (21.72, 22.49) |
| Phospholipids to Total Lipids in Large LDL % | Relative lipoprotein lipid concentration | 22,44 (21,93, 22,99) | 22,56 (22,12, 23,06) | 72.65 (71.87, 73.34) | 72.72 (71.77, 73.47) |
| Cholesterol to Total Lipids in Large LDL % | Relative lipoprotein lipid concentration | 71,76 (70,91, 72,53) | 71,33 (70,38, 72,2) | 53.31 (52.48, 54.09) | 53.87 (53.06, 54.59) |
| Cholesteryl Esters to Total Lipids in Large LDL % | Relative lipoprotein lipid concentration | 52,8 (52, 53,53) | 52,86 (52,09, 53,54) | 19.41 (18.95, 19.87) | 18.81 (18.32, 19.29) |
| Free Cholesterol to Total Lipids in Large LDL % | Relative lipoprotein lipid concentration | 19,04 (18,34, 19,67) | 18,57 (17,79, 19,21) | 5.34 (4.73, 5.97) | 5.2 (4.62, 5.92) |
| Triglycerides to Total Lipids in Large LDL % | Relative lipoprotein lipid concentration | 5,68 (5,03, 6,47) | 5,92 (5,15, 6,93) | 26.75 (26.37, 27.13) | 26.35 (25.81, 26.82) |
| Phospholipids to Total Lipids in Medium LDL % | Relative lipoprotein lipid concentration | 26,15 (25,58, 26,7) | 26,13 (25,63, 26,6) | 68.72 (67.89, 69.33) | 69.3 (68.6, 69.85) |
| Cholesterol to Total Lipids in Medium LDL % | Relative lipoprotein lipid concentration | 68,98 (68,16, 69,64) | 68,88 (68,03, 69,51) | 47.82 (46.7, 48.9) | 49.41 (48.23, 50.65) |
| Cholesteryl Esters to Total Lipids in Medium LDL % | Relative lipoprotein lipid concentration | 49,12 (47,86, 50,28) | 49,7 (48,65, 50,72) | 20.85 (20.03, 21.49) | 19.84 (18.77, 20.66) |
| Free Cholesterol to Total Lipids in Medium LDL % | Relative lipoprotein lipid concentration | 19,86 (18,84, 20,8) | 19,13 (17,94, 20,16) | 4.6 (4.09, 5.25) | 4.38 (3.94, 4.93) |
| Triglycerides to Total Lipids in Medium LDL % | Relative lipoprotein lipid concentration | 4,87 (4,3, 5,63) | 4,98 (4,35, 5,89) | 31.94 (30.97, 32.77) | 31.03 (30.2, 31.77) |
| Phospholipids to Total Lipids in Small LDL % | Relative lipoprotein lipid concentration | 31,32 (30,24, 32,46) | 30,8 (29,86, 31,78) | 64.04 (62.96, 64.85) | 64.62 (63.79, 65.3) |
| Cholesterol to Total Lipids in Small LDL % | Relative lipoprotein lipid concentration | 63,75 (62,59, 64,74) | 63,8 (62,66, 64,75) | 44.99 (44.06, 46) | 46 (45.12, 46.96) |
| Cholesteryl Esters to Total Lipids in Small LDL % | Relative lipoprotein lipid concentration | 45,84 (44,52, 47,14) | 46,4 (45,22, 47,54) | 18.94 (18.34, 19.43) | 18.67 (17.66, 19.28) |
| Free Cholesterol to Total Lipids in Small LDL % | Relative lipoprotein lipid concentration | 18,04 (16,96, 18,87) | 17,57 (16,24, 18,58) | 4.18 (3.67, 4.73) | 4.35 (3.81, 5.12) |
| Triglycerides to Total Lipids in Small LDL % | Relative lipoprotein lipid concentration | 4,79 (4,12, 5,68) | 5,22 (4,39, 6,39) | 48.67 (46.28, 50.3) | 43.48 (39.6, 46.53) |
| Phospholipids to Total Lipids in Very Large HDL % | Relative lipoprotein lipid concentration | 47,8 (44,8, 50,01) | 43,43 (39,58, 46,56) | 48.31 (47, 50.42) | 52.34 (49.72, 55.42) |
| Cholesterol to Total Lipids in Very Large HDL % | Relative lipoprotein lipid concentration | 48,32 (46,62, 50,53) | 51,01 (48,44, 54,18) | 36.29 (35.42, 37.67) | 37.79 (36.16, 39.65) |
| Cholesteryl Esters to Total Lipids in Very Large HDL % | Relative lipoprotein lipid concentration | 35,15 (33,92, 36,45) | 35,21 (33,33, 37,12) | 11.99 (11.2, 13.2) | 14.64 (13.13, 16.45) |
| Free Cholesterol to Total Lipids in Very Large HDL % | Relative lipoprotein lipid concentration | 13,26 (12,02, 14,94) | 16,17 (14,3, 18,17) | 2.75 (2.22, 3.52) | 4.02 (3, 5.58) |
| Triglycerides to Total Lipids in Very Large HDL % | Relative lipoprotein lipid concentration | 3,66 (2,72, 5,13) | 5,28 (3,72, 7,32) | 48.46 (47.81, 49.4) | 49.41 (48.38, 50.68) |
| Phospholipids to Total Lipids in Large HDL % | Relative lipoprotein lipid concentration | 49,25 (48,12, 50,71) | 50,27 (48,75, 52,16) | 48.66 (47.1, 49.64) | 46.87 (44.71, 48.56) |
| Cholesterol to Total Lipids in Large HDL % | Relative lipoprotein lipid concentration | 46,55 (43,94, 48,51) | 44,04 (40,21, 46,82) | 37.85 (36.63, 38.76) | 36.62 (34.51, 38.15) |
| Cholesteryl Esters to Total Lipids in Large HDL % | Relative lipoprotein lipid concentration | 36,11 (33,76, 37,81) | 34,22 (30,65, 36,7) | 10.78 (10.45, 11.07) | 10.35 (9.88, 10.71) |
| Free Cholesterol to Total Lipids in Large HDL % | Relative lipoprotein lipid concentration | 10,52 (10,09, 10,88) | 9,98 (9,33, 10,47) | 2.93 (2.28, 3.7) | 3.59 (2.75, 4.9) |
| Triglycerides to Total Lipids in Large HDL % | Relative lipoprotein lipid concentration | 4,11 (3,09, 5,6) | 5,69 (4, 8,06) | 45.86 (45.33, 46.36) | 46.38 (45.87, 47.07) |
| Phospholipids to Total Lipids in Medium HDL % | Relative lipoprotein lipid concentration | 46,58 (46, 47,33) | 47,26 (46,49, 48,2) | 50.63 (49.48, 51.94) | 49.27 (47.75, 50.82) |
| Cholesterol to Total Lipids in Medium HDL % | Relative lipoprotein lipid concentration | 48,68 (46,81, 50,18) | 47,06 (44,92, 48,92) | 41.61 (40.45, 42.64) | 40.76 (39.33, 42.02) |
| Cholesteryl Esters to Total Lipids in Medium HDL % | Relative lipoprotein lipid concentration | 40,06 (38,44, 41,34) | 39,05 (37,07, 40,68) | 9.13 (8.79, 9.45) | 8.65 (8.33, 8.93) |
| Free Cholesterol to Total Lipids in Medium HDL % | Relative lipoprotein lipid concentration | 8,63 (8,25, 9,01) | 8,07 (7,7, 8,44) | 3.46 (2.67, 4.33) | 4.25 (3.34, 5.26) |
| Triglycerides to Total Lipids in Medium HDL % | Relative lipoprotein lipid concentration | 4,75 (3,74, 5,92) | 5,67 (4,53, 6,95) | 56.81 (56.21, 57.49) | 56.05 (55.42, 56.75) |
| Phospholipids to Total Lipids in Small HDL % | Relative lipoprotein lipid concentration | 57,26 (56,51, 58,04) | 56,61 (55,84, 57,39) | 39.92 (39.1, 40.86) | 40.06 (38.82, 41.02) |
| Cholesterol to Total Lipids in Small HDL % | Relative lipoprotein lipid concentration | 38,67 (37,47, 39,72) | 38,63 (37,33, 39,78) | 29.54 (28.64, 30.41) | 29.96 (28.76, 30.88) |

| | | | | | |
|---|---|---|---|---|---|
| Cholesteryl Esters to Total Lipids in Small HDL % | Relative lipoprotein lipid concentration | 28,62 (27,41, 29,65) | 28,83 (27,54, 29,96) | 10.41 (10.13, 10.79) | 10.1 (9.86, 10.35) |
| Free Cholesterol to Total Lipids in Small HDL % | Relative lipoprotein lipid concentration | 10,03 (9,74, 10,37) | 9,79 (9,54, 10,07) | 3.15 (2.6, 3.8) | 3.89 (3.29, 4.6) |
| Triglycerides to Total Lipids in Small HDL % | Relative lipoprotein lipid concentration | 4,09 (3,37, 4,9) | 4,79 (4,04, 5,66) | 5.19 (4.58, 5.82) | 5.19 (4.53, 5.83) |

**Supplementary Table S8.** General information and assessment of adiposity indices in UK Biobank and Epirus Health Study. Abbreviations: ABSI, a body shape index; BMI, body mass index; EHS, Epirus Health Study; HC, hip circumference; HI, hip index; UKB, UK Biobank; WC, waist circumference; WHR, waist to hip ratio; WHI, waist to HI index.

| **Adiposity** | **Measurement** | | **UKB Data-Field** |
|---|---|---|---|
| | **UKB** | **EHS** | |
| **Height** | Standing height was measured using a Seca 202 device | Standing height was measured using a Seca 202 device | **50.0.0** |
| **Weight** | Weight was measured by various means during the initial Assessment Centre visit. This field amalgamates these values into a single item. | Weight was measured using Seca equipment. | **21002.0.0** |
| **Body Fat %** | Body composition estimation by impedance measurement. | Body composition estimation by impedance measurement. | **23099.0.0** |
| **BMI** | **Weight / Height$^2$** | **Weight / Height$^2$** | |
| **WC** | Measuring tape | Measuring tape | **48.0.0** |
| **HC** | Measuring tape | Measuring tape | **49.0.0** |
| **WHR** | WC / HC | WC / HC | |
| **ABSI** | WC(mm)*Weight(kg)$^{-2/3}$*Height(m)$^{5/6}$ | WC(mm)*Weight(kg)$^{-2/3}$*Height(m)$^{5/6}$ | |
| **HI** | **Female**: HC(cm)*Weight(kg)$^{-0.482}$*Height(cm)$^{0.31}$ **Male**: HC(cm)*Weight(kg)$^{-2/5}$*Height(cm)$^{1/5}$ | **Female**: HC(cm)*Weight(kg)$^{-0.482}$*Height(cm)$^{0.31}$ **Male**: HC(cm)*Weight(kg)$^{-2/5}$*Height(cm)$^{1/5}$ | |
| **WHI** | WHR*Weight(kg)$^{-2/5}$*Height(cm)$^{1/5}$ | WHR*Weight(kg)$^{-2/5}$*Height(cm)$^{1/5}$ | |

### Supplementary Figures

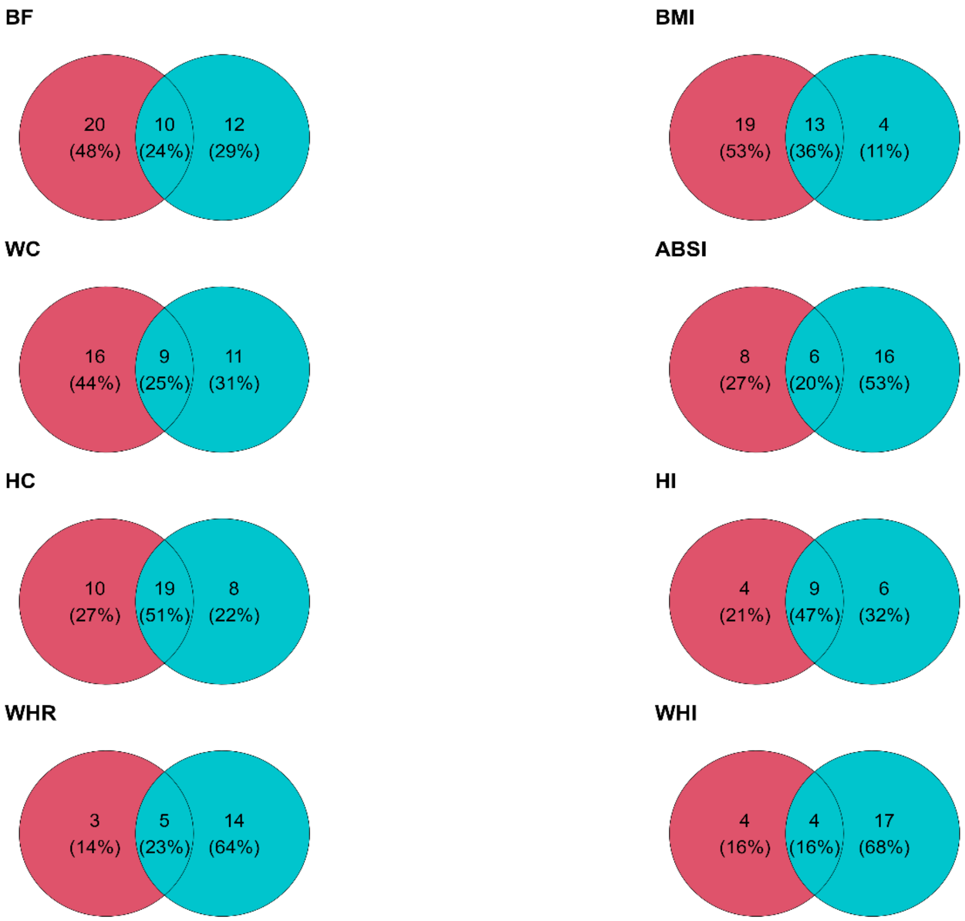


**Supplementary Figure S1.** Overlaps of the metabolic signatures between females (red) and males (light blue). Abbreviations: ABSI, a body shape index; BF; body fat %, BMI, body mass index; Hip, hip circumference; HI, hip index; WC, waist circumference; WHI, waist to HI index; WHR, waist to hip ratio.

**(Female)**

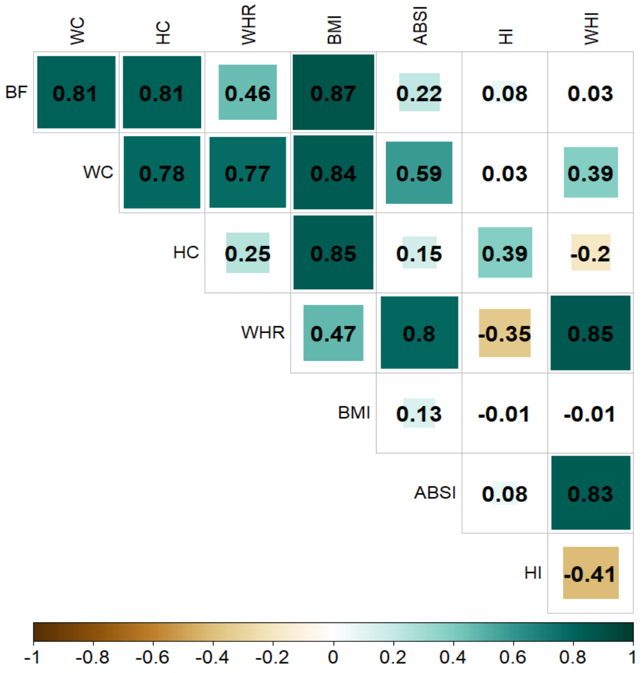


**(Male)**

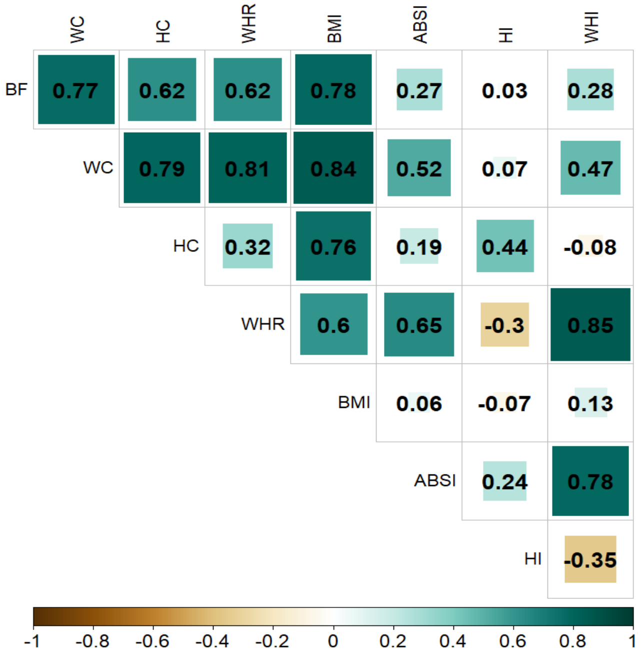


**Supplementary Figure S2.** Pearson correlation coefficients between eight body adiposity indices by sex. Green and brown colors present higher or lower correlations, respectively. Color intensity presents the magnitude of the association. Abbreviations: ABSI, A Body Shape Index; BMI, body mass index (kg/m$^2$); HI, Hip Index.

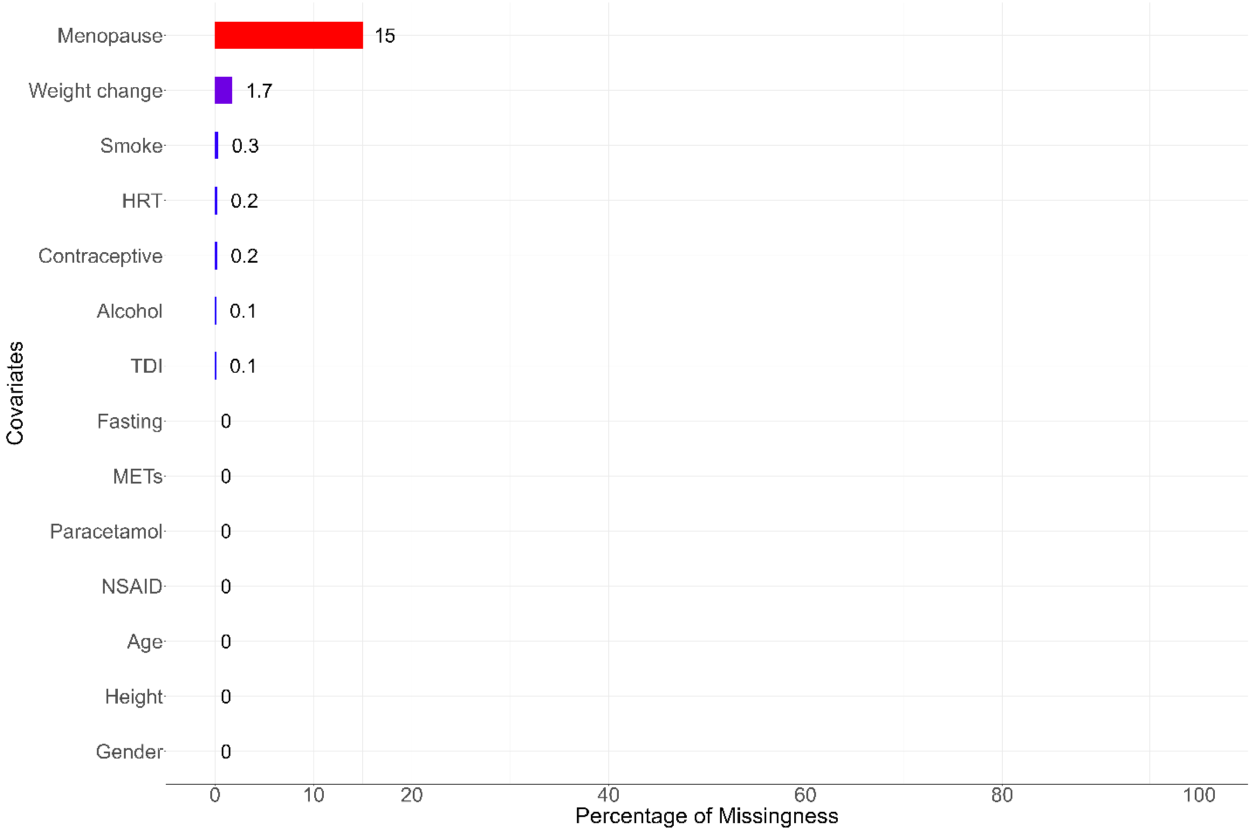


**Supplementary Figure S3.** Missing values percentages of the covariates used in analyses. Abbreviations: HRT, hormone replacement therapy; METs, metabolic equivalents of task; NSAID, non-steroidal anti-inflammatory drugs; TDI, town deprivation index.

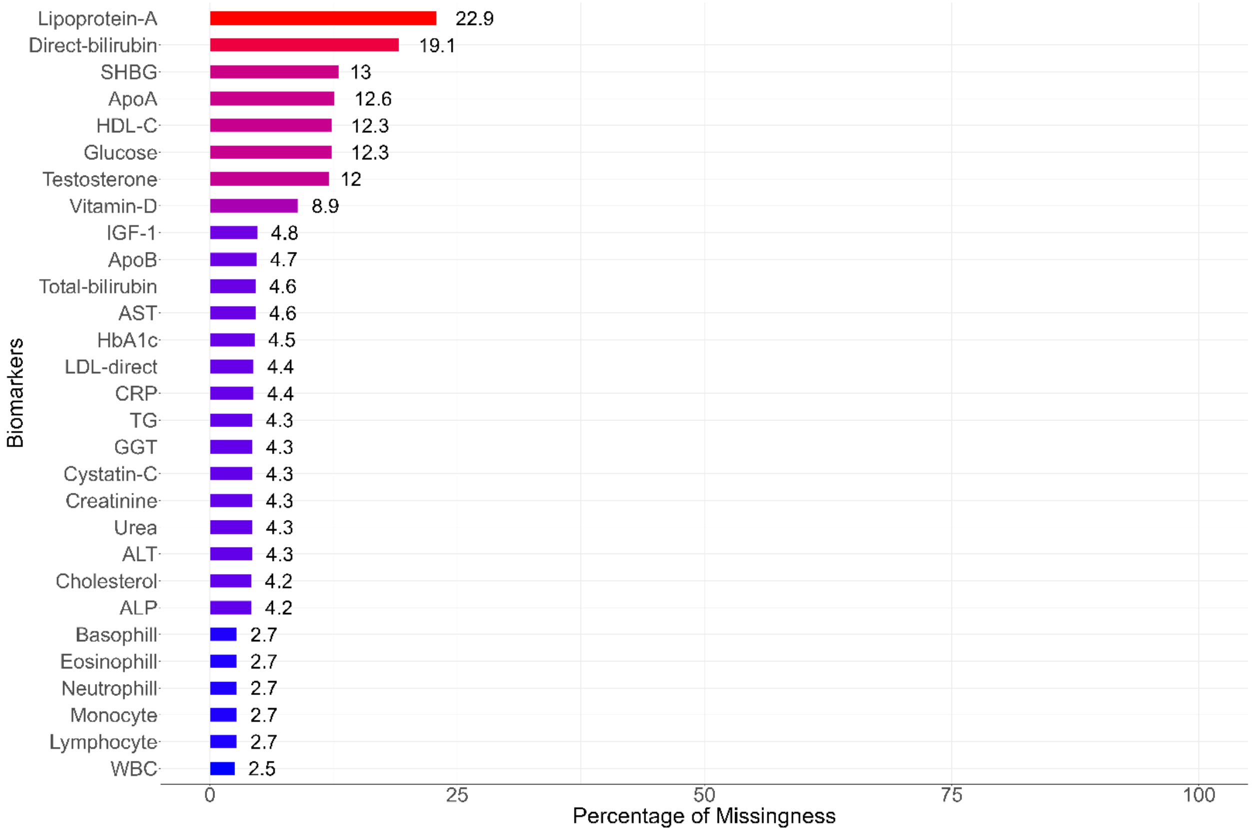


**Supplementary Figure S4.** Missing values percentages of the UK Biobank clinical biomarkers. Abbreviations: ALP, alkaline phosphatase; ALT, alanine aminotransferase; Apo, apolipoprotein; AST, aspartate aminotransferase; CRP, c-reactive protein; GGT, gamma glutamyltransferase; HbA1c, glycated hemoglobin; TRG, triglycerides; WBC, white blood cells.

**(Female)**

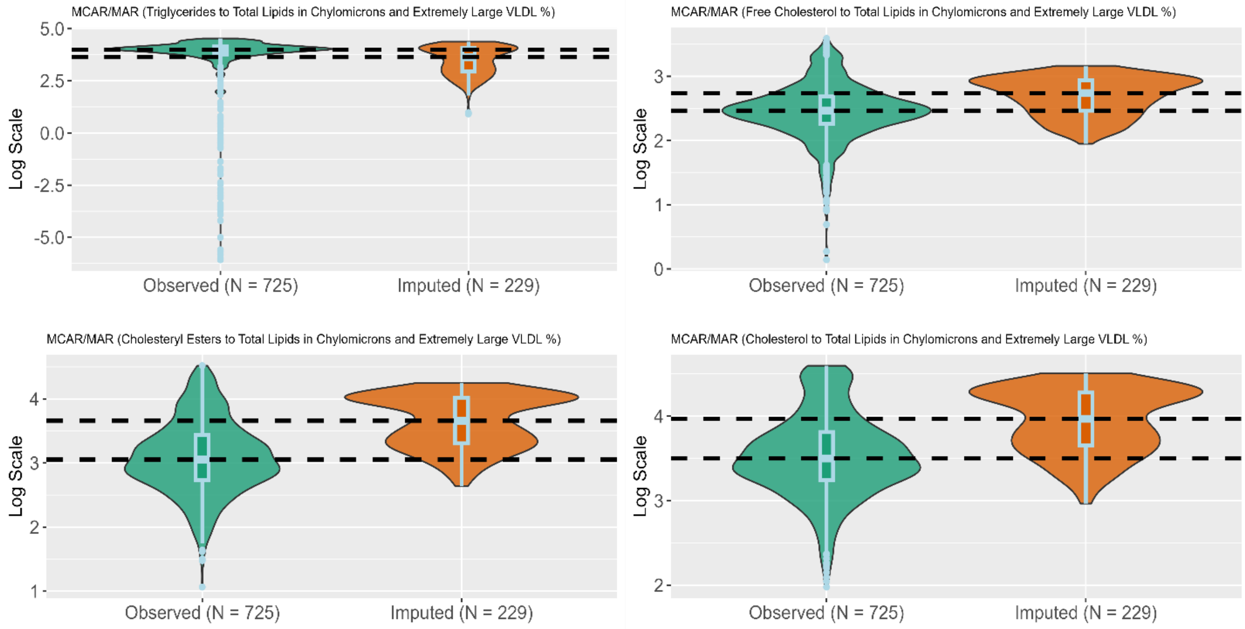


**(Male)**

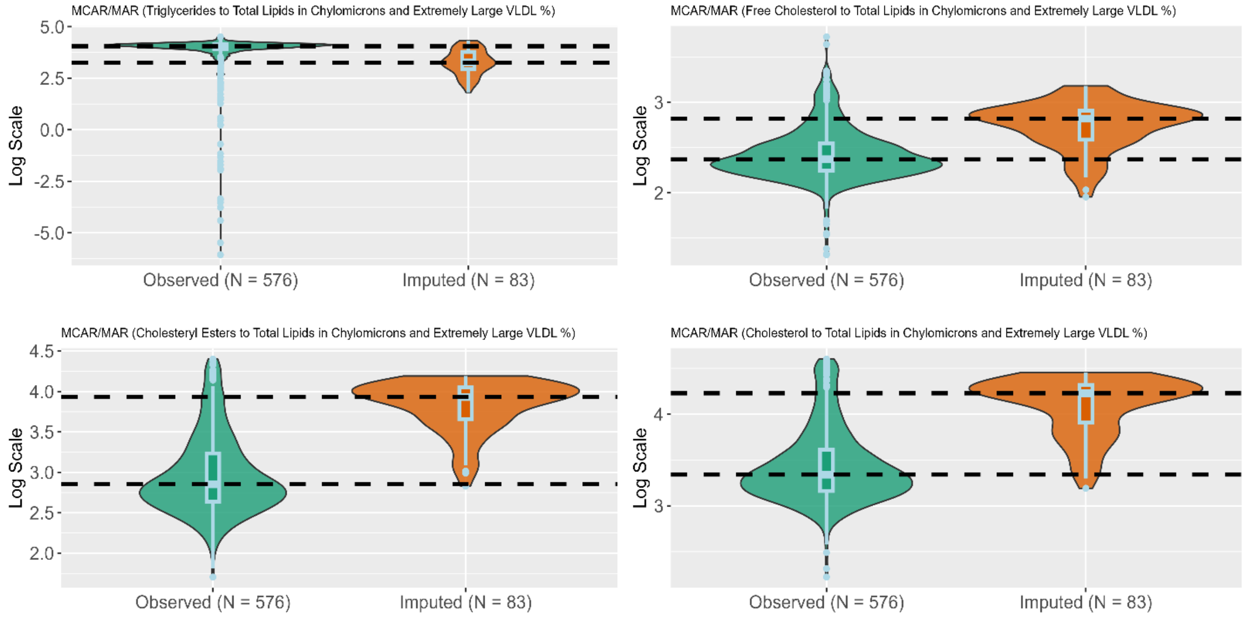


**Supplementary Figure S5.** This figure shows sex-specific distributions of observed and imputed nuclear magnetic resonance metabolites in the Epirus Health Study. Metabolites that are presented had the highest percentage of missingness compared to others. Abbreviations: MAR; Missing at Random; MCAR; Missing Complete at Random.

**(Female)**

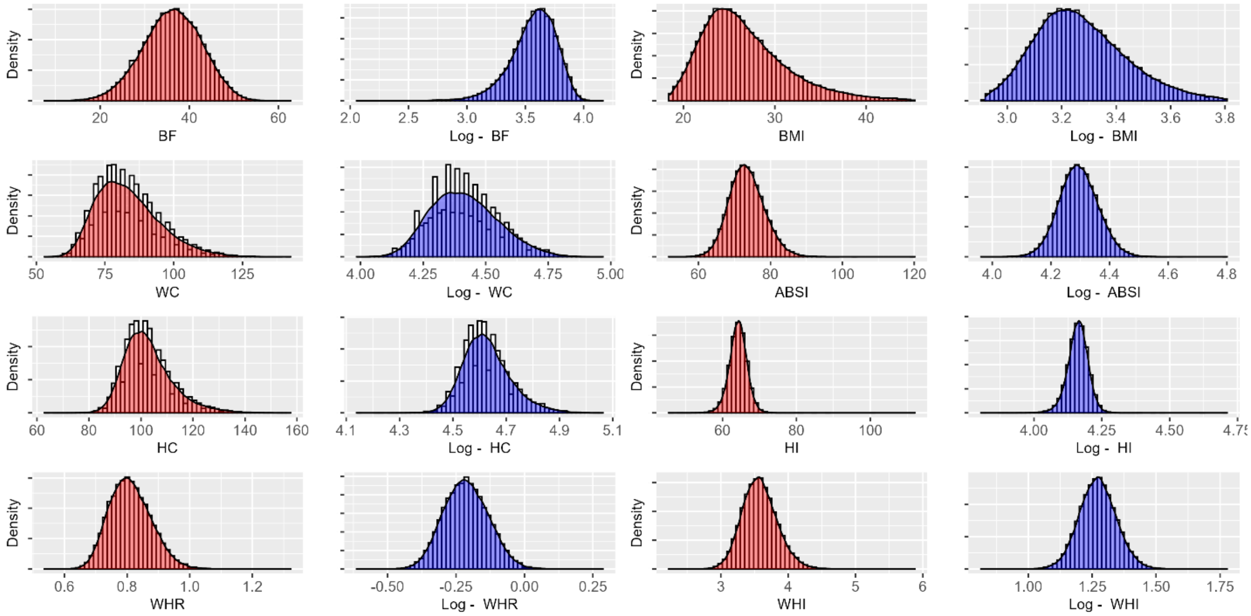


**(Male)**

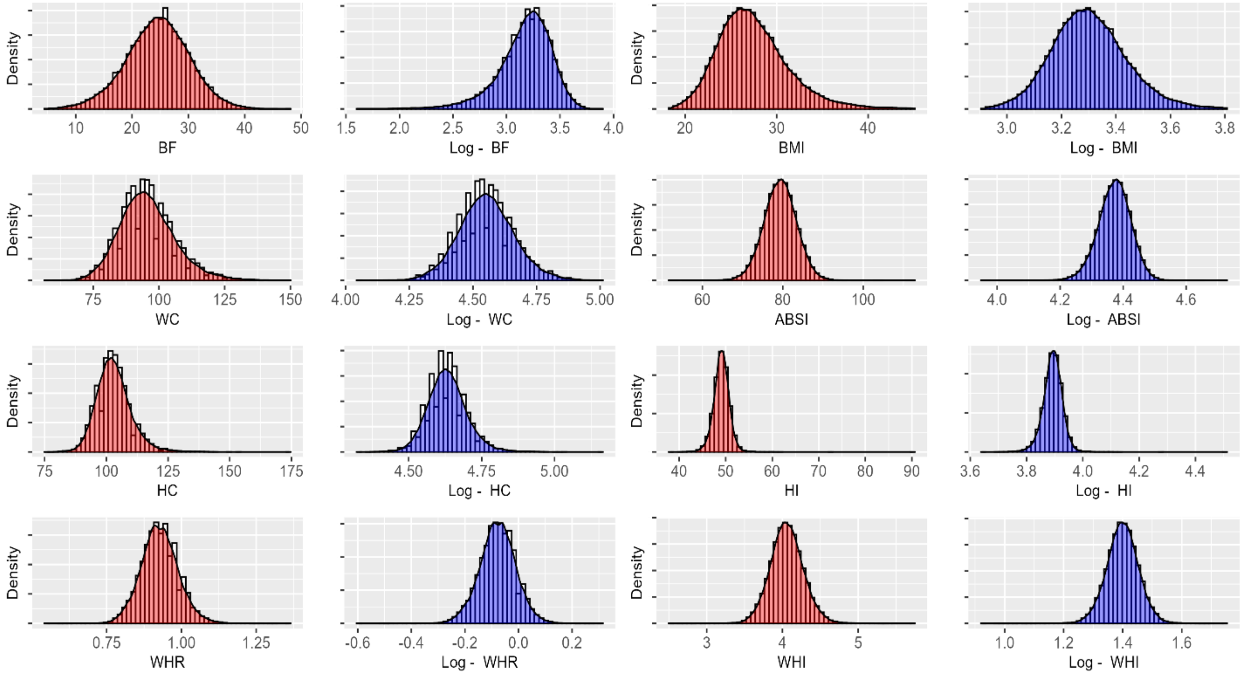


**Supplementary Figure S6.** Normal and log-transformed distributions for each body adiposity index by sex in UK Biobank. Red and blue colors present the normal and log-transformed distributions, respectively. <u>Abbreviations</u>: ABSI, A Body Shape Index; BMI, body mass index; HI, Hip Index.

**(Female)**

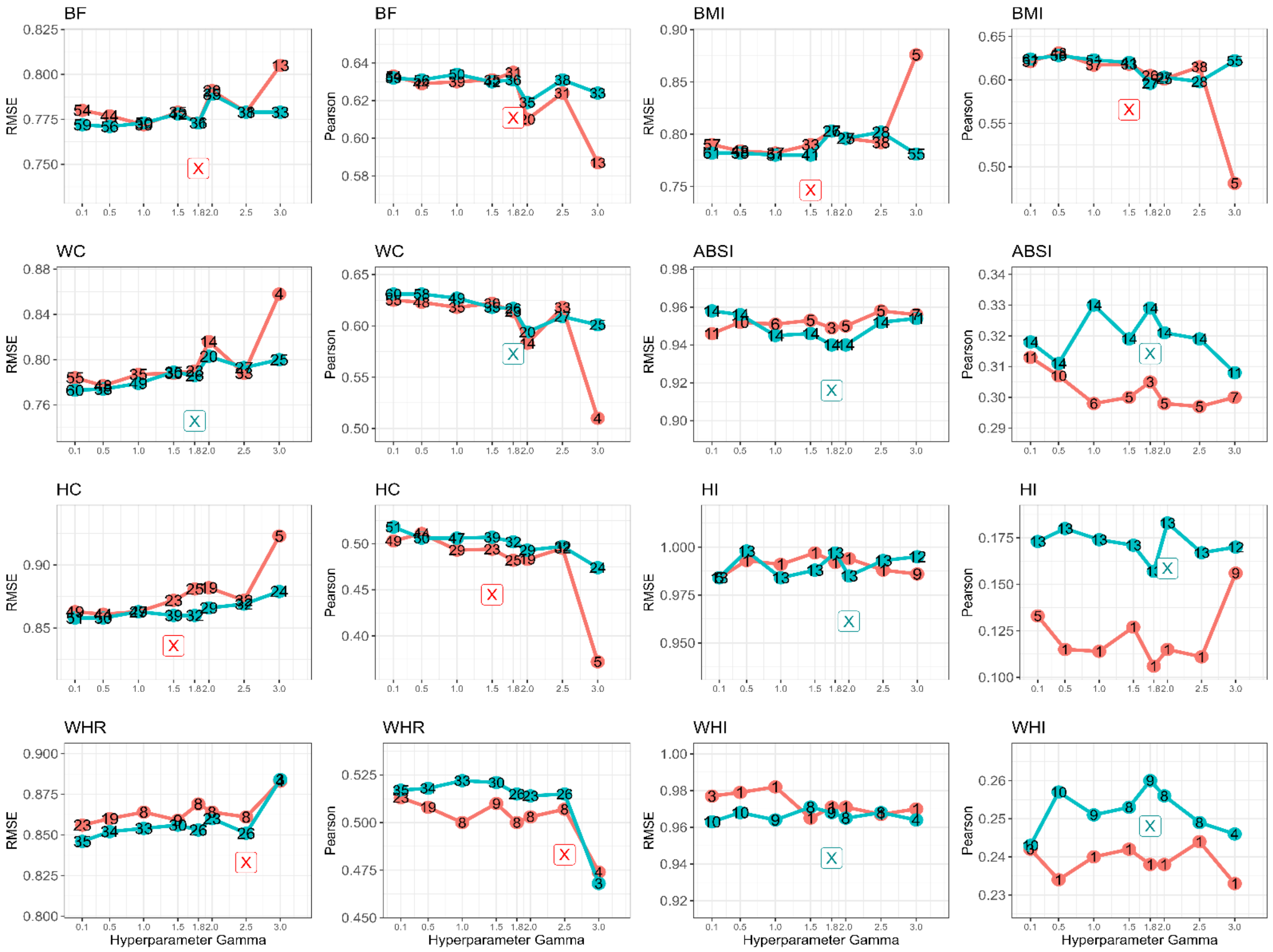
BF
BF
BMI
BMI
WC
WC
ABSI
ABSI
HC
HC
HI
HI
WHR
WHR
WHI
WHI
RMSE
Pearson
Hyperparameter Gamma

**(Male)**

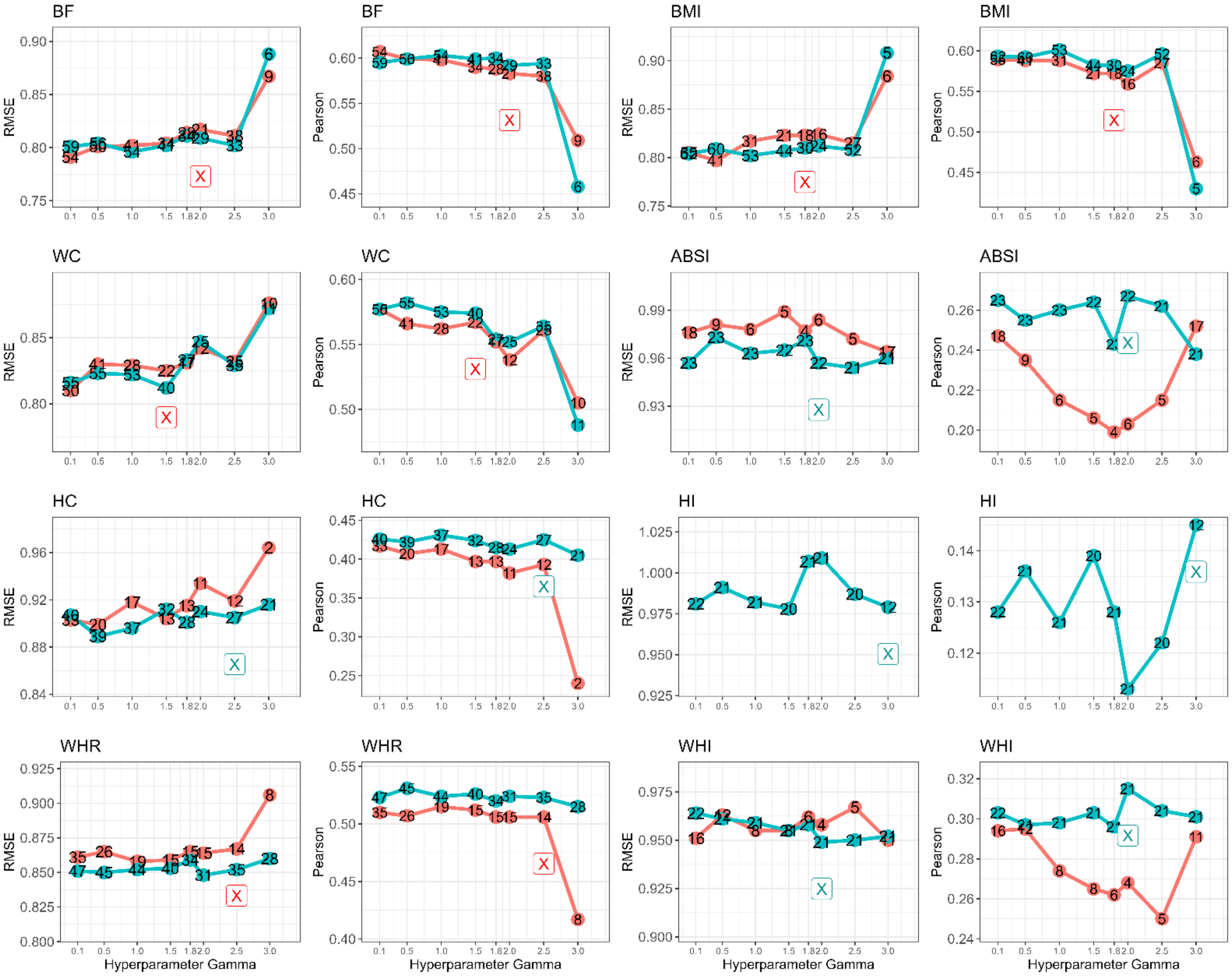


**Supplementary Figure S7**. Hyperparameter tuning and selection of the sex-stratified MSA-net for all adiposity indices. Red and blue colors present lambda min and lambda 1SE criterion, respectively. Values in circles present the number of identified metabolites after selecting the best combination of L1 and L2 norms. Colored boxes present the respective parameter selection of gamma, and lambda criterion. Abbreviations: ABSI, a body shape index; BMI, body mass index; HI, hip index, SE; standard error.
